# Two years of COVID-19 Pandemic: Framework of Health Interventions in a Brazilian City

**DOI:** 10.1101/2022.08.05.22278481

**Authors:** Vanessa dos Santos Faiões, Helvécio Cardoso Corrêa Póvoa, Bruna Alves Thurler, Gabriela Ceccon Chianca, Andréa Videira Assaf, Natalia Lopes Pontes Póvoa Iorio

## Abstract

The COVID-19 pandemic and its effects on public health have urgently demanded effective health policies to avoid the spread of COVID-19. Thus, public administrators have implemented non-pharmacological and pharmacological interventions to mitigate the pandemic’s impacts and strengthen health services. The aim of this ecological study is to describe the scenario of COVID-19 pandemic in a Brazilian city, during two years. This ecological study was carried out in Nova Friburgo, a Brazilian city, for 105 weeks (two years), from March 29, 2020 (week 1) to April 02, 2022 (week 105). Data on COVID-19 cases and COVID-19 deaths, occupation of COVID-19 exclusive beds in hospitals, community mobility, vaccination, government regulation on the opening of city establishments and city risk assessment were collected from public datasets. Four waves of COVID-19 cases and deaths were observed during this period. The first case occurred in week 1 and first death in week 3 of this study. The highest peaks of cases and deaths were observed during the third wave with 1,131 cases (week 54) and 47 deaths (week 55) and where the highest occupation of COVID-19 exclusive beds in local hospitals occurred. Interventions from more restrictive to more flexible, were implemented throughout this study, including lockdown and gradual return in economic and social strata levels. Vaccination began on week 43 and at the end of this study 89.91% of the total population was vaccinated with at least one dose, being 83.22% fully vaccinated. A deep description of several interventions used to avoid COVID-19 spread in a Brazilian city during two years of this pandemic can help promote better decision-making in the future while it exposes the challenges of conducting public health policies in a pandemic scenario.

## Introduction

The current pandemic of a coronavirus-associated acute respiratory disease called coronavirus disease 19 (COVID-19) is caused by severe acute respiratory syndrome coronavirus 2 (SARS-CoV-2) - previously known as 2019 novel coronavirus [1]. The distribution throughout the host body cells of SARS-CoV-2 receptors, angiotensin-converting enzyme 2 (ACE2) and transmembrane protease serine 2 (TMPRSS2), immune-evasive and immuno-regulative viral properties are responsible for the organ-specific manifestations of COVID-19, proliferation and progression of the disease. The outcomes of COVID-19 vary according to the disease spectrum, with a significant proportion of morbidity and mortality in patients that present severe to critical illness. It is also observed that a combination of immunocompromised hosts with overwhelming viral load may result in poor outcomes [2].

To date (Aug 05, 2022) the COVID-19 pandemic has spread to 230 countries, areas or territories with a total of 587,379,861 and 6,433,118 confirmed cases and deaths, respectively [3]. In Brazil, the first case of COVID-19 was reported in São Paulo, the largest Brazilian city, on February 26, 2020, by a man with travel history from Italy [4]. COVID-19 was responsible for 33,994,470 cases and 679,802 deaths in Brazil. Considering deaths per population, Brazil presents the sixteenth highest rate with 3,151 deaths/million people through August 05, 2022 [3].

Several public health interventions such as lockdown, curfew, closure and restrictions on establishment openings, personal hygiene, environmental sanitation, adequate and appropriate use of masks, and vaccination, were effective in mitigating the burden of COVID-19 [5-15]. Although the lack of mobilization by certain leaders combined with harsh social conditions suggest a scenario of political disarticulation that impacts the dynamics of the pandemic, such as the critical situation observed in the Brazilian state of Amazonas, the description of these perspectives allows for a panoramic evaluation of health crisis management [16].

The aim of this study is to describe the interventions adopted by a Brazilian city to avoid the spread of COVID-19 and its indicators such as: cases and deaths; cases by age group, sex and residential area; occupation of COVID-19 exclusive beds in hospitals; and community mobility.

## Methods

### Study design and city profile

This is an ecological study about the landscape of non-pharmacological and pharmacological interventions, as well as social and health indicators, adopted during the COVID-19 pandemic in the city of Nova Friburgo, Rio de Janeiro, Brazil. This study includes data collected between March 29^th^, 2020, (week of the first notified COVID-19 case in Nova Friburgo) [17] and April 02^nd^, 2022, from open websites.

Nova Friburgo (S 22° 16’ 55”, W 42° 31’ 52”) [18] is located in the mountainous region of Rio de Janeiro state, with total territory of 935,429 Km^2^, and an estimated population of 191,664 [19]. This city has eight districts divided into rural (Riograndina, Campo do Coelho, Amparo, Lumiar, São Pedro da Serra and Mury) and urban zones (Nova Friburgo and Conselheiro Paulino) [20], with approximately 87.76% of the population residing in the latter [21].

The main economic activities in Nova Friburgo include tourism, textile and metallurgic industries, cut flowers, olericulture and goat culture [18], resulting in gross domestic product per capita R$ 28,107.56 (approximately 5,986.70 US dollars) and human development index 0.745 [19]. The local health service has four tertiary care hospitals, three private and one public. The public hospital has 249 beds, and 49 of them are in the Intensive Care Unit (ICU), while private hospitals have 251 beds, being 36 ICU beds [22].

### Data collection

All data were collected from open websites: COVID-19 monitoring panel of Rio de Janeiro State Health Department [17], city’s official transparency portal [23-26], city’s official social media [27], city’s official COVID-19 monitoring panel [28], Nova Friburgo’s official website [29], city’s official news portal [30] and Google COVID-19 Community Mobility Reports [31]. The researchers received previous training, by the coordinator of this study, in searching through databases and clustering data; which were then grouped per week from Sunday to Saturday, totalling 105 weeks (Figure 1). The data was initially collected by one researcher and checked by another. The conflicting data was checked again by one of the researchers and confirmed by the other.

**Figure 1.**
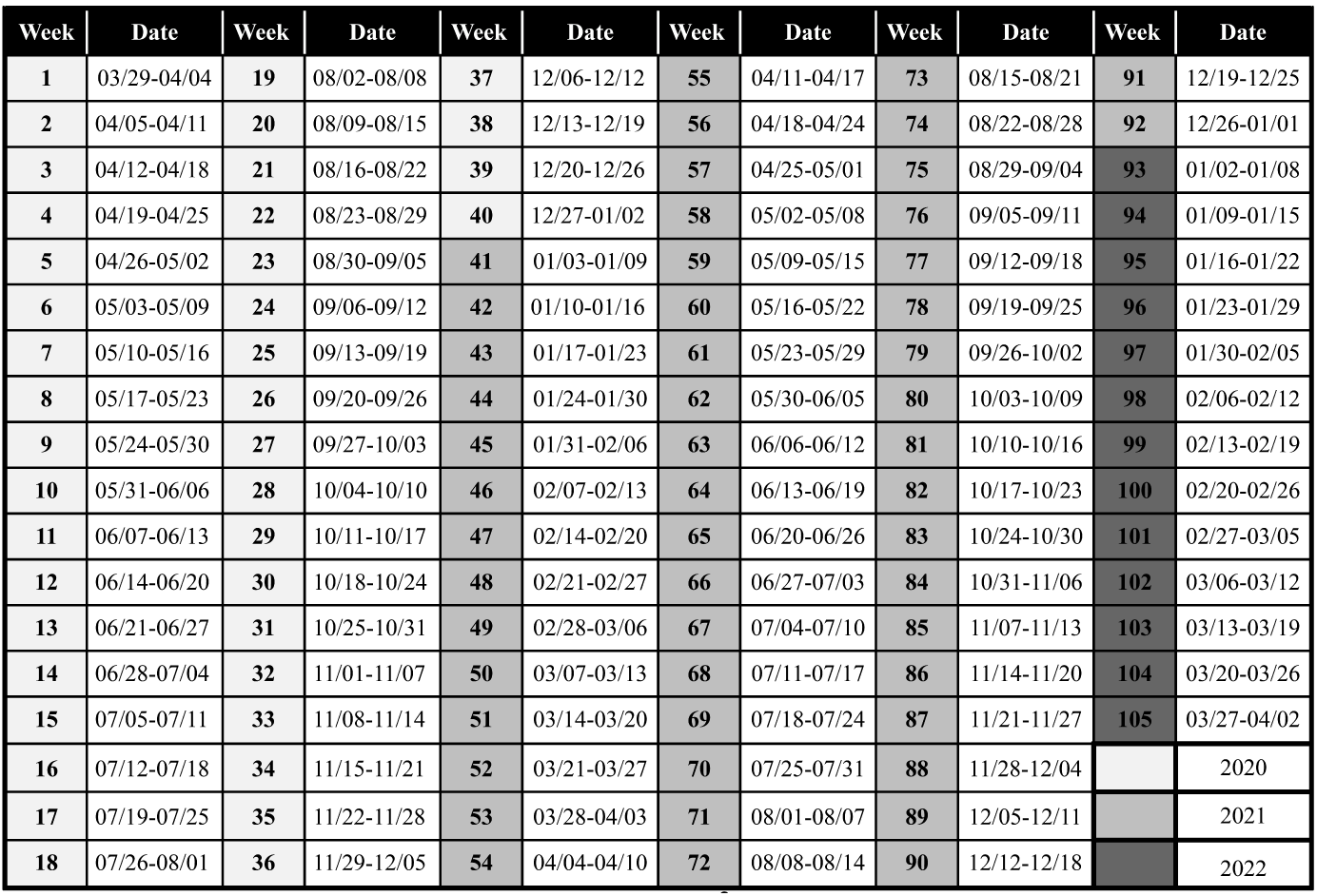
One hundred and five weeks that compose the timeframe of this study.

#### City establishment regulamentation and health events

Data regarding regulation on the functioning of city establishments during the pandemic was directly extracted from municipal decrees [24] and each business or social segment (eg: industries, stores, restaurants, tourism) was organized in no more than five categories (encoded by a combination of letters and signals) in order to clarify the degree of flexibility and restriction in each segment. Regulation for functioning of establishments was set according to the decree valid for the most number of days in that week. Information regarding the city’s main health events related to the COVID-19 pandemic were obtained from three different sources [27, 29, 30].

#### Color flags

Nova Friburgo’s colored flag coding systems involved risk assessment (very low, low, moderate, high and very high), according to the city of Nova Friburgo [27, 29] and according to the state of Rio de Janeiro [17], following indicators presented in Figure 2. The other 15 surrounding cities that compose the mountainous region of Rio de Janeiro and this entire region also received their own colored flag coding systems according to the state of Rio de Janeiro [17]. The estimated population of these 16 cities were also obtained [32].

**Figure 2.**
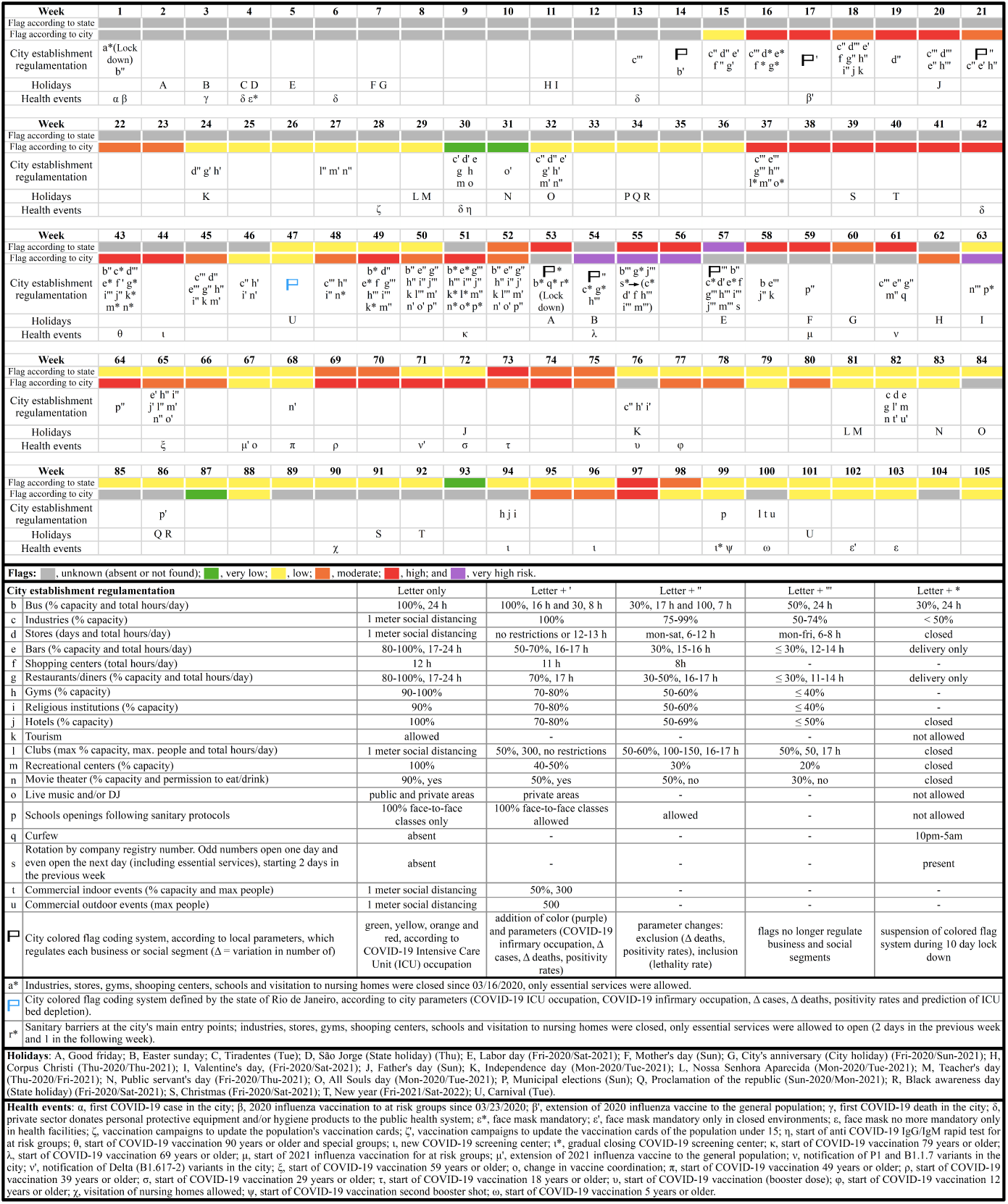
Timeline highlighting the major municipal decrees, local health events, and municipal, state and national holidays in Nova Friburgo. Each lower case letter not represented in the following week indicates the permanency of the regulative measures from the previous one.

#### COVID-19 cases and COVID-19 deaths

The number of new COVID-19 cases/week was obtained by subtracting the total cases of the current week from the total of cases in the previous one, considering the last data published in each week [27, 25]. The same methodology was used to calculate the number of COVID-19 deaths/week.

#### COVID-19 cases/groups

The percent of accumulated COVID-19 cases per age group (≤ 9; 10-19; 20-29; 30-39; 40-49; 50-59 and ≥ 60 years old), sex and residential area (urban/rural) was observed weekly based on data from the last available day of each week [28]. The age groups <1, 1-4, 5-9 years old were added up to maintain the 10 year difference between age groups.

#### COVID-19 beds

The number of COVID-19 infirmary and Intensive Care Unit (ICU) beds in the city’s hospitals was calculated by average occupancy and availability per week, according to the data published in the city’s official social media [27]. Only weeks with at least three days of available data were included.

This study also assessed the presence or absence of COVID-19 exclusive infirmary and ICU beds, in public hospitals from all cities in Rio de Janeiro’s mountainous region [17].

#### Community mobility

Community mobility in Nova Friburgo was verified by Google LLC “Google COVID-19 Community Mobility Reports” [31]. Average movement per week for each category (n=6) was calculated only when at least three days/week of information were available.

#### Vaccination progress

Data from the last available day of each week [26, 27] were grouped in fully and partially vaccinated for COVID-19. Individuals are considered fully vaccinated 14 days after receiving all recommended doses in the primary series [33]. For each week the percentage of fully and partially vaccinated individuals was calculated by F = (A/B) x 100 and P = (C/B) x 100, respectively. Where, F = percentage of fully vaccinated individuals; A = number of fully vaccinated individuals; B = city’s total population [21]; P = percentage of partially vaccinated individuals; C = number of partially vaccinated individuals. Additionally, data from individuals who received a booster dose after all recommended doses in their primary series were also included in this study. Data were replicated from the previous week for weeks without available data.

## Results

The effect of municipal decrees and local health events in Nova Friburgo; and municipal, state and national holidays were condensed in a timeline (Figure 2). This timeline also shows Nova Friburgo’s colored flag coding systems according to both the city of Nova Friburgo and the state of Rio de Janeiro. According to the city of Nova Friburgo, the flag system began on week 15 and classified COVID-19 risk by colored flags: four weeks with very high risk (purple), 21 with high risk (red), 18 with moderate (orange), 27 with low (yellow), and three with very low risk (green). The state of Rio de Janeiro started attributing flags for Nova Friburgo on week 47 and totalized 56 flags, being one purple, eight red, seven orange, 39 yellow and one green. After the colored flag systems were instated, 18 and three colored flags according to the city and the state of Rio de Janeiro, respectively, were not found for Nova Friburgo. When the city’s colored flag coding system regulated business and social segments (weeks 15-56), lockdown (week 53) was the only time Nova Friburgo did not receive a colored flag. During the 105 weeks of study, business segments were allowed to open between 84 and 91 weeks, according to their specific features, while social segments between 48 and 86 weeks.

Figure 3 shows the state’s colored flag system for each of the 16 cities of the mountainous region of Rio de Janeiro and for this entire region starting on week 47, totalizing 59 flags for the entire mountainous region and 56 for each individual city within this region. This region has an estimated population of 981,159, with an average of 61,322 per city, ranging from 5,646 to 307,144.

**Figure 3.**
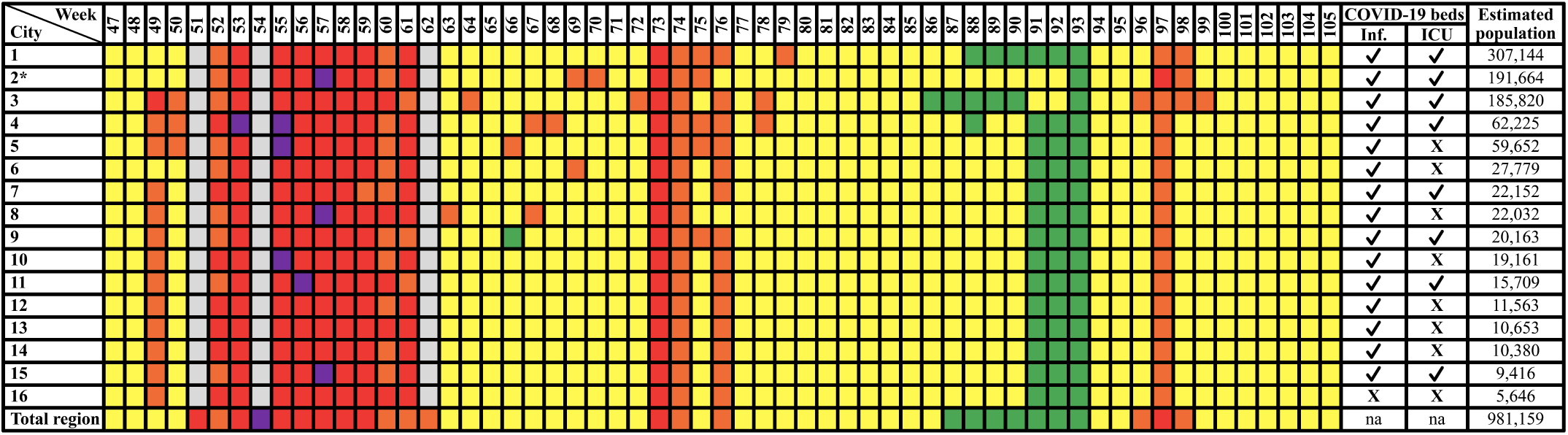
State color flag system for all 16 cities of the mountainous region and for this entire region of the state of Rio de Janeiro. Cities: 1, Petrópolis; 2, Nova Friburgo; 3, Teresópolis; 4, Guapimirim; 5, Cachoeiras de Macacu; 6, Bom Jardim; 7, Cordeiro; 8, São José do Vale do Rio Preto; 9, Cantagalo; 10, Carmo; 11, Sumidouro; 12, Duas Barras; 13, Trajano de Moraes; 14, Santa Maria Madalena; 15, São Sebastião do Alto; 16, Macuco; * City where this study was conducted. Flag colors: gray, unknown (absent or not found); green, very low; yellow, low; orange, moderate; red, high; purple, very high risk. ✔, present; **X**, absent; na, not applicable.

The first case of COVID-19 occurred during week 1 and the first death during week 3. It is possible to observe four waves in both COVID-19 cases and deaths/week, with peaks of 310 (week 19), 701 (week 41), 1,131 (week 54) and 709 (week 96); and 11 (week 21), 17 (week 40 and 42), 47 (week 55) and 12 (week 98), respectively. The peaks of new COVID-19 cases occur almost simultaneously to agglutination periods of flags orange and/or red and/or purple (Figure 4).

**Figure 4.**
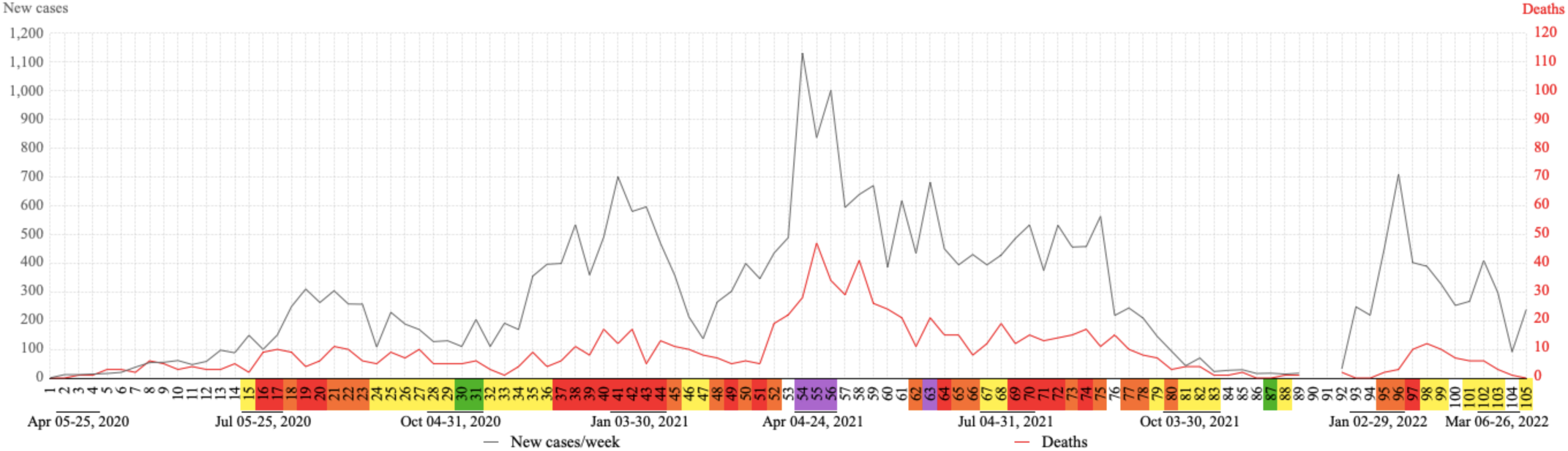
Evolution of weekly COVID-19 cases and deaths in Nova Friburgo. Flag colors: green, very low; yellow, low; orange, moderate; red, high; purple, very high risk.

The ≥ 60 age group had the highest concentration of accumulated COVID-19 cases in 69 of the 80 weeks with available data, ranging from 31.5% (week 13) to 19.8% (week 67) (Figure 5). In the last 29 weeks with available data, the highest concentration of accumulated COVID-19 cases was interchanged between the 30-39 (19.8-20.2%), 40-49 (19.9-20.5%) and ≥ 60 (19.8-20.5%) age groups. During all analyzed periods the ≤ 9 and 10-19 age groups remained below 6% of accumulated COVID-19 cases, although the latter presented a continuous increase. The 20-29 age group presented the highest percentage increase in COVID-19 cases during this study, more than 8%, when comparing week 14 (8.4%) to week 96 (16.9%). On the other hand, the ≥ 60 age group presented the highest percentage decrease, almost 12%, when comparing week 13 (31.5%) to week 67 (19.8%) (Figure 5). Considering the distribution of accumulated COVID-19 cases according to sex and residential area, in week 96, 54.5% are women and 88.5% of total positive cases live in urban areas. Data prior to week 13 and after 96 regarding age group, sex and residential area were not available, as well as data for weeks 80, 90, 91 and 95.

**Figure 5:**
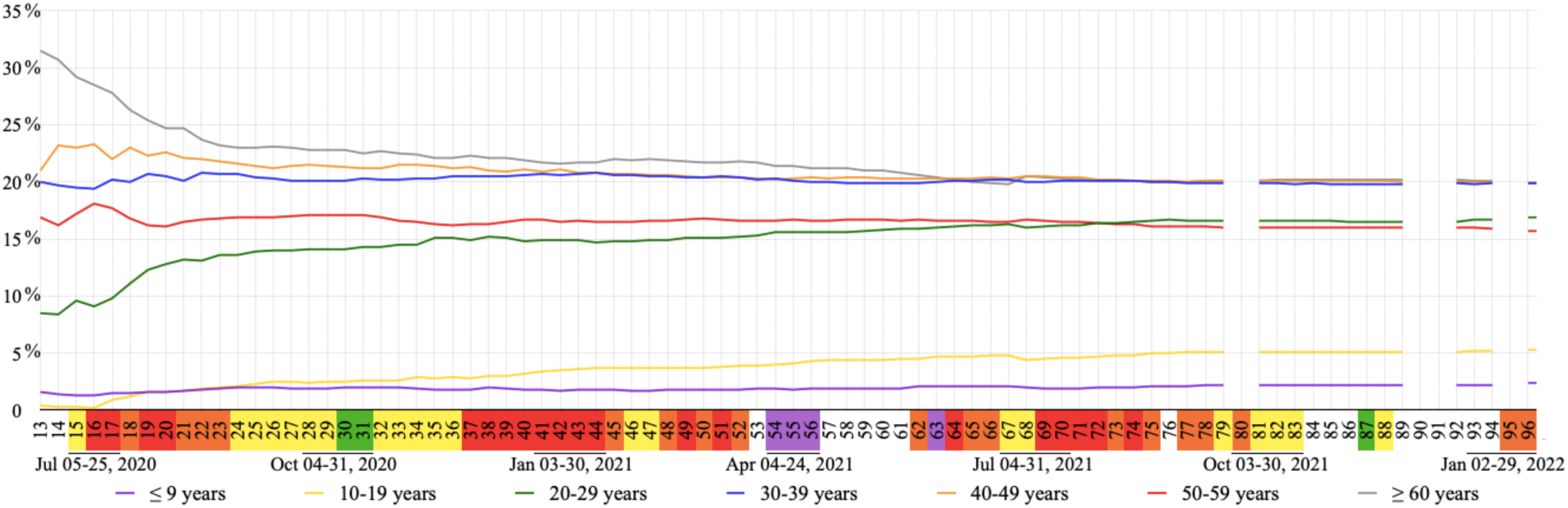
Percentage of accumulated COVID-19 cases per week in each age group. Flag colors: green, very low; yellow, low; orange, moderate; red, high; purple, very high risk.

One of the 16 cities of Rio de Janeiro’s mountainous region did not present public infirmary and ICU beds for COVID-19; other seven cities did not present ICU beds for COVID-19 in their public hospitals (Figure 3). Nova Friburgo’s public hospital offered 27.94 (±18.16) COVID-19 exclusive infirmary and 18.19 (±3.88) COVID-19 exclusive ICU beds weekly. Private hospitals in this city offered 44.57 (±14.11) and 20.7 (±6.65), respectively (Figure 6).

**Figure 6.**
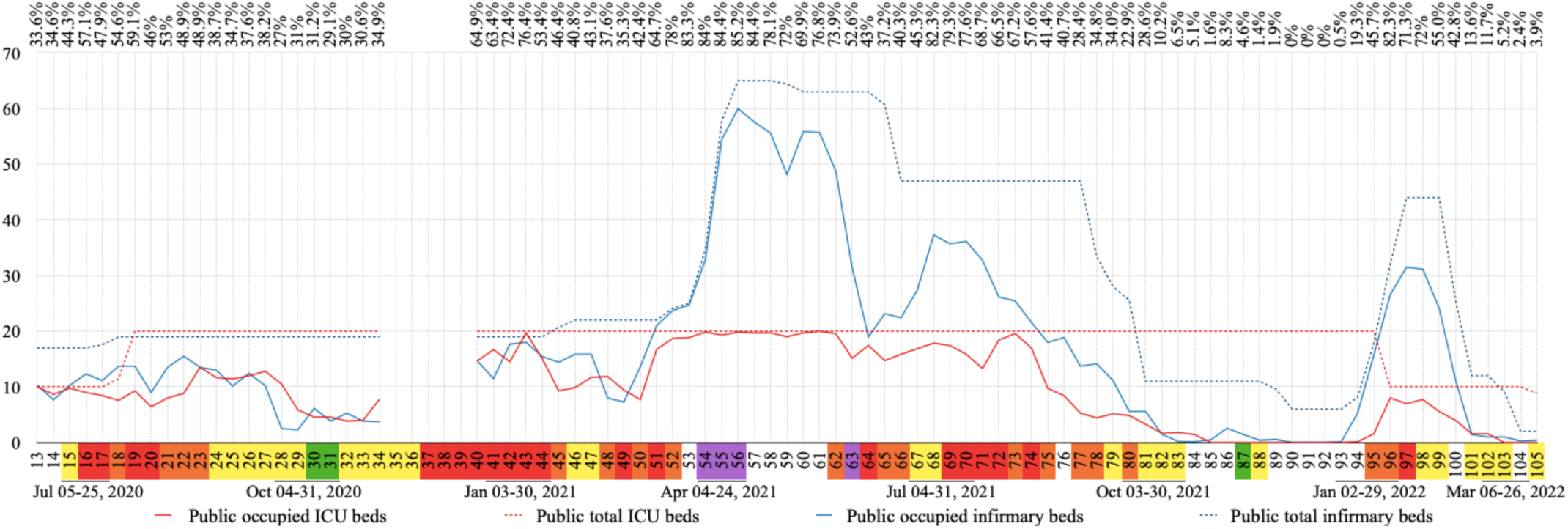
COVID-19 exclusive public bed occupation per week in absolute numbers. In the line above the graph, is the total occupation percentage for COVID-19 exclusive beds (infirmary and ICU) in public hospitals. Flag colors: green, very low; yellow, low; orange, moderate; red, high; purple, very high risk. ICU, intensive care unit.

The occupation of COVID-19 exclusive beds in public and private hospitals are shown in Figures 6 and 7, respectively. Out of 105 weeks analyzed in this study, no data was found for 17 of them. Data for at least three days in each week were found for the other 88 weeks included in this study. During this period the number of COVID-19 exclusive beds, in all four hospitals, ranged from 52 to 185, where 23 to 50 of them were in the ICU.

**Figure 7.**
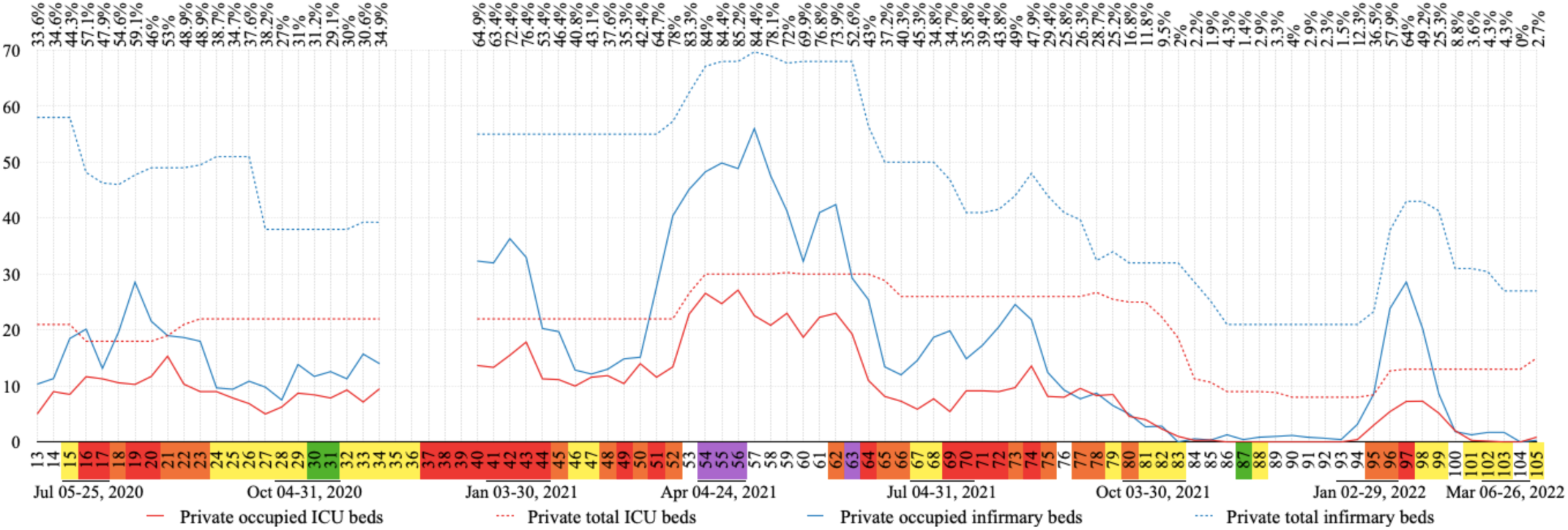
COVID-19 exclusive private bed occupation per week in absolute numbers. In the line above the graph, is the total occupation percentage for COVID-19 exclusive beds (infirmary and ICU) in private hospitals. Flag colors: green, very low; yellow, low; orange, moderate; red, high; purple, very high risk. ICU, intensive care unit.

Weekly occupancy of COVID-19 exclusive beds in public hospitals ranged from 0 to 100% for ICU and 0 to 98.9% for infirmary, while in private hospitals these rates ranged from 0 to 90.5% and 0 to 80.3%, respectively. The highest ICU bed occupation period in public hospitals was between week 52 and 62 (93.6-100%), and high occupations were also observed during weeks 13 (100%), 15 (97.5%), 43 (98.6%) and 73 (97.9%). In private hospitals this period occurred between week 53 and 56 (82.4-90.5%); where high rates were also observed during weeks 21 (80.7%) and 43 (81.2%). Infirmary beds presented the highest occupation during weeks 51-56 (92.3-98.9%) and weeks 42-43 (93-94.7%) in public hospitals, and the same was observed in private hospitals during week 57 (80.3%). Vacant COVID-19 beds in public hospitals were observed from week 85 through 93 and 103 through 105 for ICU, while for infirmary beds this was observed from week 90 through 92. In private hospitals, vacant ICU COVID-19 beds were observed from week 86 through 93 and 103 through 104 (0%). Absence of occupancy was repeated for private infirmary beds during week 83 and 104 (Figures 6-7).

Figure 8 highlights 20 different moments: nine municipal decrees and 11 holidays in Nova Friburgo. These moments were evaluated in regard to community mobility [31], according to six categories: retail and recreation, grocery and pharmacy, parks, transit stations, workplaces, and residential. Gaps in information were observed in two categories (parks and transit stations) for four weeks (21-24) with none, one or only two days of data per week. All other weeks analyzed had data for all days of the week, except the grocery category, which had data for six days during 3 weeks (21-23).

**Figure 8.**
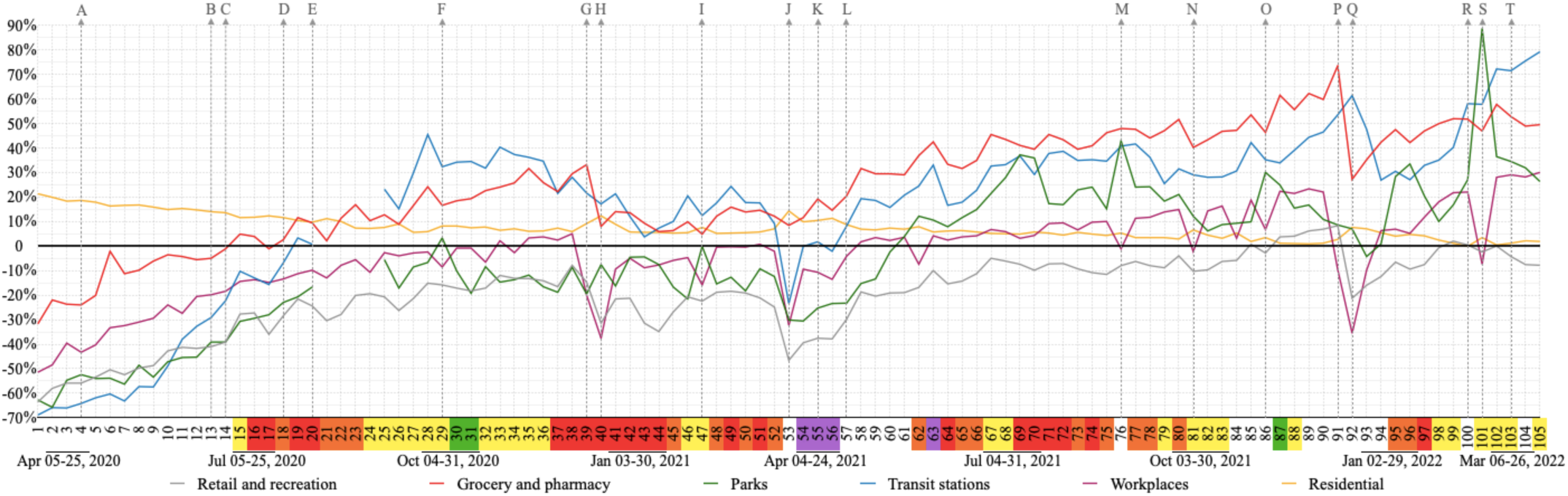
Impact of decrees and holidays on community mobility per week. Decrees: A, face mask mandatory; B, reopening industries; C, bus 100% capacity and total hours 16/day; D, reopening several business and social segments; J, lockdown; K, rotation by company registry number; L, flags no longer regulate business and social segments; R, decree commercial events one meter social distancing; T, decree for a more flexible mask use. Holidays: E, Father’s day 2020 (Sun); F, Nossa Senhora Aparecida 2020 (Mon); G, Christmas 2020 (Fri); H, New year 2021 (Fri); I, Carnival 2021 (Tue); M, Independence day 2021 (Tue); N, Nossa Senhora Aparecida 2021 (Tue); O, Proclamation of the republic and black awareness day 2021 (Mon and Sat); P, Christmas 2021 (Sat); Q, New year 2022 (Sat); S, Carnival 2022 (Tue). Flag colors: green, very low; yellow, low; orange, moderate; red, high; purple, very high risk.

After the mandatory mask use decree (week 4), all categories but one (residential) showed an increase in mobility. This increase continued after decrees for reopening industries (week 13) and expansion of bus schedules/fleet (week 14). Immediately after these decrees, grocery and pharmacy mobility crossed the baseline (week 15), and all categories, except one (residential), continued to show an increase in mobility.

Between week 18 (several business/social segments were reopened) and week 20 (Father’s day), transit stations’ mobility crossed the baseline. The unique week where park mobility was above the baseline in 2020 was on the national holiday of Nossa Senhora Aparecida (week 29).

The highest peak of grocery and pharmacy mobility in 2020 was during Christmas (week 39). During the last week of 2020 (week 40), all categories showed a decrease in their mobility with exception of residential and parks. Week 47 (carnival 2021) was the first time in 2021 that the category parks touched the baseline.

During the 10 day lockdown (mainly during week 53) all mobility categories, except one (residential) decreased. This was also the week where the highest peak of residential mobility and smallest peaks of retail and recreation and transit stations in 2021 were observed (Figura 8).

After the decree of rotation by company registry number (week 55), when odd numbers opened one day and even opened the next day (including essential services), it was possible to observe a gentle decrease of retail and recreation, grocery and pharmacy, transit stations, and workplaces mobilities. After the decree of flags no longer regulated business and social segments (week 57) the mobility categories of retail and recreation, grocery and pharmacy, parks, and transit stations increased (Figura 8).

Although weeks with holidays showed a decrease in movement for workplaces, a highlight in this decrease was demonstrated on New Years 2022. Christmas in 2021 repeated the previous year’s peak for grocery and pharmacy, representing the highest peak of this study. Later on, when the decree regulating commercial events (week 100) established one meter social distancing for such events, an increased movement in parks was observed, reaching the highest peak in this study during carnival of 2022 (week 101). In the following week, the decree for more flexible mask use occurred (week 103) and the highest peak of transit station movement was observed in the last week of this study (week 105) (Figure 8).

Figure 9 shows an increase in new cases that coincides with a decrease in transit station mobility and this category was under the baseline from week 1 through week 18 (reopening several business and social segments) and then during week 53 (lockdown) and week 56 (during the rotation by company registry number).

**Figure 9.**
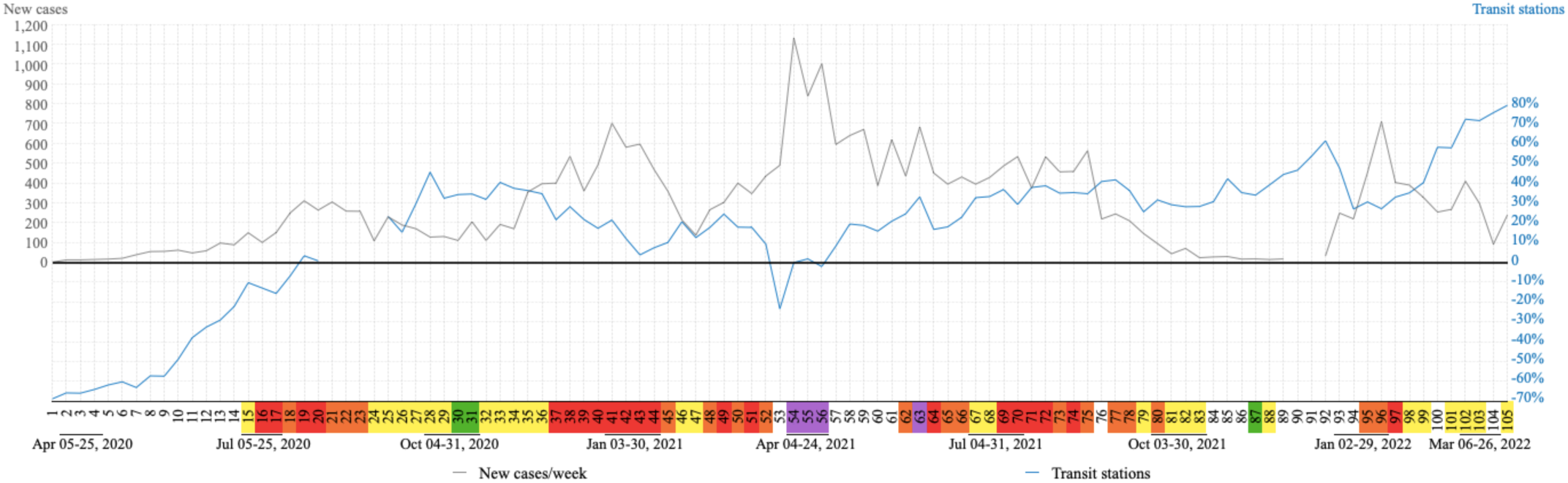
Relationship between transit station and COVID-19 cases. Flag colors: green, very low; yellow, low; orange, moderate; red, high; purple, very high risk.

The vaccination process started during week 43. During week 68, almost half of the population was vaccinated (48.03%), being 12.76% of them fully vaccinated. In week 77, when 80.45% of the population was vaccinated with at least one dose and 36.06% were fully vaccinated, the booster doses of the COVID-19 vaccine started being given to at-risk groups. The under 12 age group started receiving vaccination during week 97, when 81.76% of the population was fully vaccinated and 30.36% had received their booster dose. In the last possible analyzed week (104) of this study 89.91% of individuals had received at least one dose of the COVID-19 vaccine, being 83.21% fully vaccinated. At this time, 40.03% of the population had received their booster dose (Figure 10). Data from weeks 92, 97, 103, 104 and 105 were not available.

**Figure 10.**
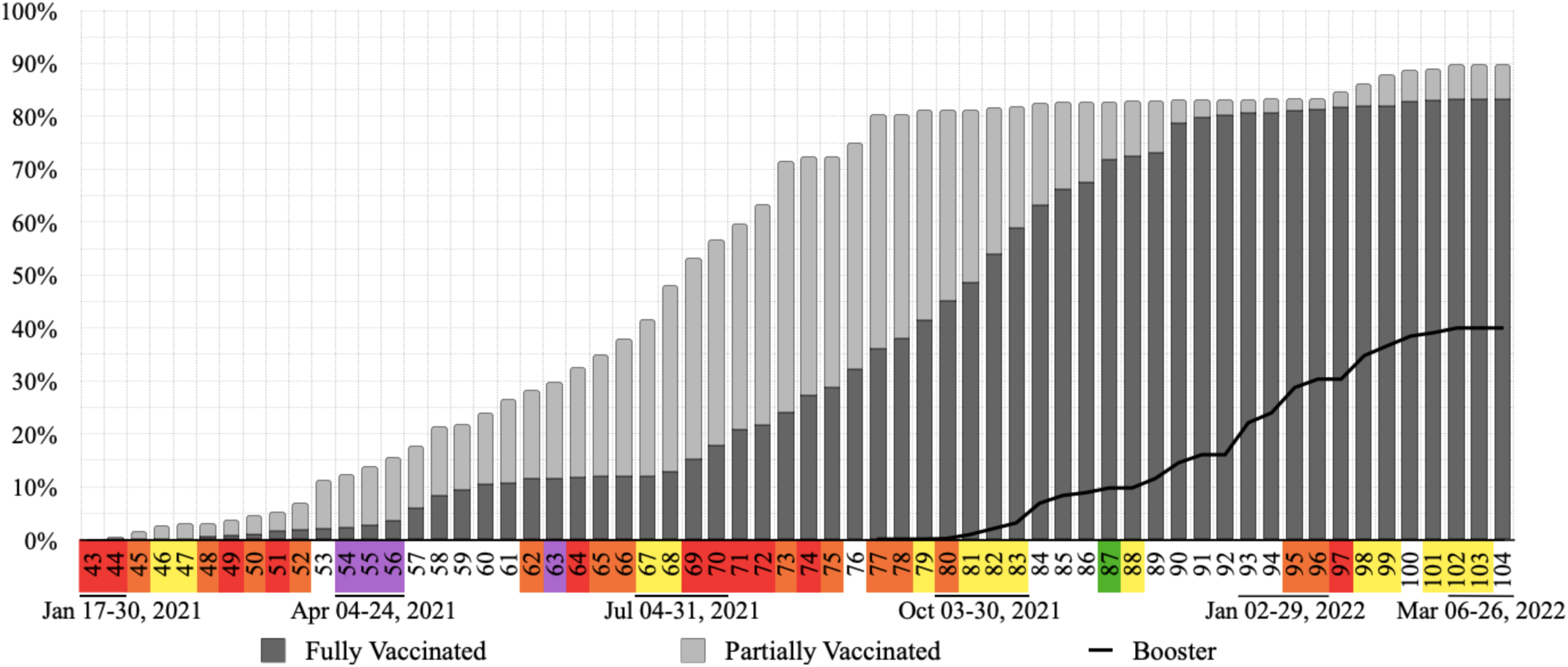
Accumulated vaccination rates per week in Nova Friburgo. Flag colors: yellow, low; orange, moderate; red, high; purple, very high risk.

## Discussion

A suitable combination of non-pharmaceutical interventions is necessary to halt the spread of COVID-19, especially in the absence of efficient antiviral medication and before the COVID-19 vaccine [6]. Thus, the initial strategy of adopting and updating the color flag systems to alert on COVID-19 risk levels could have demonstrated a concern with the collapse of the health system and helped guide recommendations and non-pharmaceutical interventions in the city of Nova Friburgo.

The city of Nova Friburgo and state of Rio de janeiro classified COVID-19 risk levels for Nova Friburgo according to different parameters (Figure 2) [17, 24]. This explains why the same week is classified with two different flag colors, one from the city and another from the state government. Additionally, during this study it was possible to observe weeks classified with the same flag color and different permissions and restrictions in business and social segments, due to municipal decrees frequently updating local restriction regulations for each flag color [24]. From week 57 on, the colored flag system according to the city of Nova Friburgo no longer regulated business and social segments, so 24 out of 25 flag colors that were not found for Nova Friburgo occurred after this decree.

In Brazil, some small cities do not have municipal hospitals. Following the principle of regionalization of Brazil’s unified health system, hospital care in small Brazilian cities is usually dependent on a consortium with larger cities [34]. According to Gomes and coworkers, the spread of COVID-19 to these small cities can affect the health system of the region’s reference larger city [35]. Half of the cities of the mountainous region of Rio de Janeiro State do not have ICU beds exclusively for COVID-19 patients in their public hospitals. This fact could have increased patient mobility between these cities, and promoted weekly clusters of flag colors within the cities of this region as observed in Figure 3.

In this study common patterns were observed between the waves of COVID-19 cases and deaths, where the cases waves start prior to the deaths waves. This becomes clear in the third wave, which begins on week 48 and 52 for COVID-19 cases and deaths, respectively. This gap could be justified by the mean time from disease onset to death, approximately 17-18 days [36, 37].

The high number of COVID-19 cases reflects on more restrictive flags (orange, red and purple) and consequently less transit station mobility. This can justify the mirror effect reflection on the “x axis”, observed in Figure 9, between the increase in new cases of COVID-19 and decrease in transit station mobility. All four waves of cases and deaths were preceded by moments of flexibility.

The first wave happened after reopening industries and increasing bus circulation; during this wave, several business and social segments were also reopened. Retail and recreation, grocery and pharmacy, parks, transit stations and workplaces continued their increase in mobility according to Google LLC “Google COVID-19 Community Mobility Reports”[31], during this period.

The decree for mandatory mask use (week 4) might have been a contributing factor for the mobility profile observed during this first wave due to Nova Friburgo’s strong textile segment. During week 19, although the city had a red flag, an exception was made to allow stores to open on that particular Saturday (day before Father’s day) which may have fostered that Saturday as the day of greatest mobility of transit stations amongst weeks 1-19 (data not shown).

Increases in accumulated COVID-19 cases in the 10-19 and 20-29 age groups were observed after the start of economic recovery during week 15. The percentage of industry capacity was expanded, and stores, bars, shopping centers and restaurants/diners were reopened during this week, which might suggest that these age groups may be the largest portion of the production and consumer lines of such establishments. In contrast, the age group ≥ 60 showed the largest percentage decrease during this period as municipal decrees recommended at-risk groups be put on paid leave and follow the health recommendations for this age group seeking their protection from COVID-19 [24]. This group had an overall high concentration of accumulated COVID-19 cases during this study, especially during the first weeks. It is important to highlight that the elderly and health professionals composed the priority testing group for COVID-19 [30], especially in the beginning of the pandemic, when the tests were limited.

The second wave came after the first two (of a total of three) green flags attributed by the city of Nova Friburgo and consequently increased flexibility. Although restrictive decrees were initially instated at the beginning of this wave (week 37), only after more restrictive decrees (week 43) was it possible to observe the start of its decline. Throughout this second wave a decrease in mobility was observed in four categories: retail and recreation, grocery and pharmacy, transit stations and workplaces. However, the increase in mobility in the grocery and pharmacy category associated with Christmas (week 39) could have contributed to an enlarged wave as well as its highest peaks during this second wave (weeks 41-43).

Curfews, lockdown, store and restaurant closures, cancellation of gatherings, mandatory work from home and the closure of educational institutions are the most effective non-pharmacological interventions [6]. During the second wave, educational institutions remained closed, but clubs were closed only four weeks before the peak, and restaurants and recreational centers only closed two weeks after the peak of this wave. Stores and shopping centers remained open throughout the second wave; curfew and lockdown were not instituted at this time.

The pharmacological intervention for COVID-19 in Nova Friburgo city started during week 43. The city immunized, during the analyzed period, its population against COVID-19 with vaccines from AstraZeneca (Oxford), CoronaVac (Sinovac), Pfizer (BioNTech) and Janssen (Johnson & Johnson). In week 45, during the decrease of the second wave, only 1.67 % of the population was vaccinated and nobody was fully vaccinated.

The third wave reached the highest values of COVID-19 cases and deaths and purple flags were only observed during this wave. The peak in number of deaths by COVID-19 for this study was observed during week 55, when only 14.85% of the population were vaccinated, being 3.65% fully vaccinated. During this wave more restrictive actions were observed, such as: curfew, rotation by company registration number and the return of lockdown. The highest offer, in number, of COVID-19 exclusive beds in hospitals was observed during this wave. Although bed occupancy progressed, no field hospitals were opened in the city (data not shown).

An increase of almost 300% in the number of COVID-19 exclusive public infirmary beds was observed during the third wave. In contrast, the number of COVID-19 exclusive public ICU beds which had doubled during the first wave (20 ICU beds) remained the same during the third wave. It is important to highlight that the third wave showed a greater number of weeks with total occupation of public ICU beds. In Rio de Janeiro state, there were patients waiting for public ICU and infirmary beds throughout this study [17], so the total public bed occupation observed in Nova Friburgo probably also occurred in many public hospitals in the state of Rio de Janeiro.

Lockdown can reduce COVID-19 rate transmission by 81% [5]. This intervention returned for 10 days during the third wave, when only essential services were allowed to open and all businesses and social segments were closed. According to Sharma and coworkers [10] closing businesses is an effective measure to control infection. Although during this decree mobility had decreased for all categories, except one (residential), the peak of COVID-19 cases was observed immediately after the end of lockdown. This could be justified by the delay between transmission and the manifestation of symptoms [7], combined with a possible delay in notification of COVID-19 cases. On the other hand, flexibility of non-pharmacological interventions at the end of the second wave could have had an impact on the number of COVID-19 cases observed in the third wave.

Nighttime curfew decree occurred only during the third wave and lasted eight weeks, which were flanked by a conglomerate of mostly red flags in Nova Friburgo and all cities in the mountainous region. The curfew weeks were also flanked by high rates of COVID-19 bed occupation, including total occupation of public ICUs. Sharma and coworkers [10] observed that nighttime curfews reduced COVID-19 transmission by around 13% in 114 regions of seven European countries; this non-pharmaceutical intervention and mandatory mask use can help curb transmission associated with face-to-face businesses.

The rotation by company registration number included all business, social segments and essential services. This decree can be responsible for a break, after lockdown, in the upward trend of increasing the mobility of five categories (retail and recreation, grocery and pharmacy, parks, workplaces and transit stations).

In week 67, during the third wave, the coordinator for the municipal vaccination strategy changed and vaccination began for all people aged ≥ 50. According to Moore and coworkers [8], at the end of the first phase of the vaccination programme, when it is offered to all people older than 50, strong non-pharmacological interventions still are required to avoid surges of infection.

Six weeks after the change in coordinator (week 73), at least one dose was offered to all people older than 18 and the percentage of fully vaccinated individuals doubled. Changes in this coordination and expansion of vaccination days could have been responsible for advances in vaccination rates and the beginning of booster vaccination, during week 77.

The offer of vaccination to all individuals aged 18-29 (week 72-73) could be related to a break in the upward trend of COVID-19 accumulated cases within these age groups. Stability of COVID-19 accumulated cases for all analyzed age groups observed during week 82 (end of the third wave), could be associated with the percentage of fully vaccinated individuals which exceeded 50%.

In 2021, a study suggested that the effectiveness of any isolated non-pharmaceutical intervention can be limited, while a combination of multiple interventions appeared more effective in reducing the spread of COVID-19 [9]. When comparing the first three COVID-19 waves in Nova Friburgo, a possible trend was observed where each wave was approximately two times bigger than the previous one. Suggesting that if other measures (or other combinations of interventions) to mitigate the dissemination of the virus had been adopted, these possible trends might have been different. In contrast, this trend was not observed for the fourth wave, which was similar to the second one.

The period between the end of the third and beginning of the fourth wave was characterized by the first weeks with no deaths in this study (weeks 86, 87, 93 and 94), absence of occupation of ICU beds by COVID-19 patients in all four hospitals in the city (weeks 86-93), more than 80% of the population fully vaccinated (week 92), peak in mobility of grocery and pharmacy, and an increase in transit station mobility at the end of 2021 (weeks 91-92).

The fourth wave was marked by the return of hospital occupation, although this occupation did not last as long as it did during the other waves and no total occupation of ICU and infirmary beds was observed. During this wave we observed a break in the increase of the transit station mobility; however this break was not similar to the previous one, which was responsible for values below the baseline.

During the fourth wave, mobility for most categories was above the baseline which may be related to increased flexibility, traditionally celebrated holiday in the state (Carnival) and repressed tourist demands. Although Carnival was not officially decreed, mobility in workplaces decreased, as typical during the holidays. Additionally, the largest peak of mobility in parks was observed during this timeframe which could be associated with rural tourism in the city. This was previously observed in Portugal, where the growth of tourism in rural areas may be associated with better conditions for social distancing and safety [38].

During the third wave of COVID-19 cases in Nova Friburgo city, the variants gamma and delta were predominant in the state of Rio de Janeiro and in Brazil, while omicron was predominant during the fourth wave [17, 39, 40]. So, COVID-19 cases observed during each wave of this study could be related with different predominance of virus variants and lineages.

In week 104, the last one updated for vaccination, 89.91% of the population received at least one dose of the vaccine, being 83.22% fully vaccinated. At this moment 40% were also vaccinated with booster doses. It is important to highlight that children under 5 years old were not part of the vaccination schedule.

Studies have demonstrated different levels of neutralizing capacities of antibodies by COVID-19 vaccines for different SARS-CoV-2 variants [41-43]. Booster vaccines have demonstrated an increased protection for mild COVID-19 disease, although this protection reduces over time [44]. This dose might also offer greater protection against severe and fatal disease and is effective in preventing severe COVID-19-related outcomes [44, 45]. Booster doses also increase production of neutralizing antibodies that are not susceptible to escape by variants [46]. Additionally, vaccinated people who were also exposed to SARS-CoV-2, called hybrid immunity, present numerous SARS-CoV-2-specific memory B cells that are not susceptible to escape by variants and higher neutralizing antibody titers [46, 47].

The high rates of fully vaccinated individuals, booster doses and hybrid immunity could explain the different profile observed during the fourth wave even in a moment with more flexibility.

COVID-19 effects seriously impact health systems and the economy [48]. To balance measures that protect the health of the population, but negatively impact the economy is a challenge [7]. Viana and coworkers [15] suggest that the rush to restart socioeconomic activities could be associated with new COVID-19 waves, while Milne and coworkers [7] report the negative impacts of strict social distancing on the economy. It’s important to highlight that non-pharmacological interventions should not be completely removed once the vaccination programme is complete [8], reinforcing possible social and economic impacts.

Identification of the Omicron variant in the state of Rio de Janeiro [17, 39] and its circulation in Nova Friburgo [17], after a period with no COVID-19 hospitalizations at local hospitals as well as greater social and business flexibility, could demonstrate that the dynamics of COVID-19 control is a fragile equilibrium. So, we strongly recommend that measures such as: epidemiological surveillance, genomic monitoring, scientific researches and development of new vaccines to old and new pathogens are strengthened, in order to understand the natural history of diseases, prevent the spread of pathogens and new pandemics, reducing negative impacts on public health and the economy.

This ecological study has inherent limitations to its design. We detected five main limitations in this study. (i) The dataset relies on information available in secondary data from public sources. It is important that the data are quickly and readily available; (ii) Possible underreporting of COVID-19 cases, as the number of detected cases is not necessarily the same as the number of true cases. Low testing rates and asymptomatic cases may explain this difference; (iii) We observed absence of information during some weeks and/or days of this study, limiting the completeness of the evaluated data. Additionally information such as offer and occupation of COVID-19 hospital beds, COVID-19 cases according to age group, sex and residential area were not available from the first to twelfth week of this study; (iv) The use of Google COVID-19 Community Mobility Reports. These reports are from anonymized data only from users who have turned on their location history setting, which is off by default. The data are also dependent on connectivity and privacy thresholds, in order to guarantee anonymity. So, this data represents the mobility behavior of a sample of Google users of the Nova Friburgo population; (v) As any ecological study, associations must be performed carefully, because low sensitivity correlations through unstudied confounding factors may be present. As well as the ecological fallacy, intrinsic to the study design.

## Conclusion

To the best of our knowledge, no previous study performed a deep description of the several interventions used to avoid COVID-19 spread and their effects in a Brazilian city, during a long period of the COVID-19 pandemic. Although the authors recognize the limitations of this study, we strongly believe that this does not detract from the validity of the study, since the description of these interventions and their effects are important to understand COVID-19 spread and pandemic response. In conclusion, in this framework of 105 analyzed weeks in a Brazilian city during the COVID-19 pandemic, we describe the main interventions adopted and their suggestive correlation with COVID-19 spread and possible implications for local public health. This provides elements that facilitate the interpretation of data, through associations of interventions that can potentially help or hinder the impacts of the COVID-19 pandemic on the epidemiological scenario.

## Data Availability

All data used in this study are from open websites and, therefore, publicly available. The raw data extracted from these sites are included in a supplementary file.

## Acknowledgements

Fundação Carlos Chagas Filho de Amparo à Pesquisa do Estado do Rio de Janeiro (FAPERJ) and Coordenação de Aperfeiçoamento de Pessoal de Nível Superior (CAPES).

## Author Contributions

Conception, design, data collection, analysis, interpretation, writing, review and edition were performed by Vanessa Dos Santos Faiões. Helvécio Cardoso Corrêa Póvoa and Gabriela Ceccon Chianca contributed to the design, data analysis, interpretation and revision. Bruna Thurler Alves contributed with data collection, interpretation and revision. Andréa Videira Assaf contributed to the conception, design, data analysis, interpretation and revision. Conception, design, data collection, analysis, interpretation, writing, revision, edition and supervision were performed by the coordinator of this study, Natalia Lopes Pontes Póvoa Iorio. All authors contributed to the critical reviews and approved this manuscript.

## Disclosures

### Compliance with Ethics Guidelines

This article is based on open data that did not apply to ethical approval.

