## Supplementary file for "Two years of COVID-19 Pandemic: Framework of Health Interventions in a Brazilian City"

Helvécio Cardoso Corrêa Póvoa[b]

Bruna Alves Thurler[b]

Gabriela Ceccon Chianca[c]

Andréa Videira Assaf[a,d]

Natalia Lopes Pontes Póvoa Iorio[a,b]

[a]- Postgraduate Program in Dentistry, Universidade Federal Fluminense (UFF), Instituto de Saúde de Nova Friburgo, Nova Friburgo, Rio de Janeiro, Brazil

[b]- Department of Basic Science, Universidade Federal Fluminense (UFF), Instituto de Saúde de Nova Friburgo, Nova Friburgo, Rio de Janeiro, Brazil

[c]- School of Pharmacy, Universidade Estácio de Sá (UNESA), Nova Friburgo, Rio de Janeiro, Brazil

[d]- Department of Specific Formation, Instituto de Saúde de Nova Friburgo, Universidade Federal Fluminense (UFF), Nova Friburgo, Rio de Janeiro, Brazil

Declarations of interest: none

Correspondence to Dr. Natalia Lopes Pontes Póvoa Iorio, Fluminense Federal University, Nova Friburgo, RJ, Brazil

Rua Doutor Sílvio Henrique Braune, 22 – Centro, Nova Friburgo, Rio de Janeiro Brazil, CEP- 28625-650

#### Supplementary appendix

1 Database table of cases and deaths in Nova Friburgo 2

2 Database table of accumulated cases per groups 12

3 Database table of COVID-19 exclusive beds 15

4 Database table of vaccination progress in Nova Friburgo 29

5 Database table of community mobility in Nova Friburgo 33

6 Description of the database sources in this manuscript 51

7 Description of the color flag system indicators 53

### 1 Database table of cases and deaths in Nova Friburgo

| Date | Deaths | Cases |
|---|---|---|
| 2020-04-03 | | 1 |
| 2020-04-06 | | 4 |
| 2020-04-07 | | 7 |
| 2020-04-08 | | 7 |
| 2020-04-10 | | 13 |
| 2020-04-13 | | 16 |
| 2020-04-15 | 1 | 22 |
| 2020-04-17 | 1 | 25 |
| 2020-04-20 | 1 | 33 |
| 2020-04-22 | 2 | 33 |
| 2020-04-24 | 2 | 39 |
| 2020-04-27 | 3 | 43 |
| 2020-04-29 | 4 | 47 |
| 2020-05-01 | 5 | 55 |
| 2020-05-04 | 5 | 65 |
| 2020-05-06 | 7 | 69 |
| 2020-05-08 | 8 | 75 |
| 2020-05-11 | 8 | 89 |
| 2020-05-13 | 9 | 97 |
| 2020-05-15 | 10 | 113 |
| 2020-05-18 | 12 | 121 |
| 2020-05-20 | 13 | 137 |
| 2020-05-22 | 16 | 167 |
| 2020-05-25 | 18 | 190 |
| 2020-05-29 | 21 | 222 |
| 2020-06-01 | 23 | 230 |
| 2020-06-03 | 24 | 270 |
| 2020-06-05 | 24 | 283 |
| 2020-06-08 | 28 | 311 |
| 2020-06-10 | 28 | 325 |
| 2020-06-12 | 28 | 330 |
| 2020-06-15 | 29 | 342 |
| 2020-06-17 | 30 | 365 |
| 2020-06-19 | 31 | 388 |

| | | |
|---|---|---|
| 2020-06-22 | 32 | 407 |
| 2020-06-24 | 32 | 434 |
| 2020-06-25 | 32 | 445 |
| 2020-06-26 | 34 | 485 |
| 2020-06-29 | 34 | 504 |
| 2020-06-30 | 35 | 516 |
| 2020-07-01 | 37 | 545 |
| 2020-07-03 | 39 | 573 |
| 2020-07-06 | 39 | 603 |
| 2020-07-08 | 40 | 661 |
| 2020-07-09 | 40 | 705 |
| 2020-07-10 | 41 | 722 |
| 2020-07-13 | 45 | 750 |
| 2020-07-14 | 48 | 771 |
| 2020-07-15 | 49 | 790 |
| 2020-07-16 | 49 | 802 |
| 2020-07-17 | 50 | 822 |
| 2020-07-20 | 53 | 853 |
| 2020-07-21 | 54 | 870 |
| 2020-07-22 | 55 | 906 |
| 2020-07-23 | 58 | 940 |
| 2020-07-24 | 60 | 972 |
| 2020-07-27 | 62 | 1033 |
| 2020-07-28 | 63 | 1077 |
| 2020-07-29 | 66 | 1118 |
| 2020-07-30 | 68 | 1169 |
| 2020-07-31 | 69 | 1221 |
| 2020-08-03 | 69 | 1298 |
| 2020-08-04 | 73 | 1354 |
| 2020-08-05 | 73 | 1399 |
| 2020-08-06 | 73 | 1463 |
| 2020-08-07 | 73 | 1531 |
| 2020-08-10 | 77 | 1589 |
| 2020-08-11 | 78 | 1621 |
| 2020-08-12 | 78 | 1709 |
| 2020-08-13 | 78 | 1742 |
| 2020-08-14 | 79 | 1794 |
| 2020-08-17 | 82 | 1841 |
| 2020-08-18 | 83 | 1896 |
| 2020-08-19 | 88 | 1978 |
| 2020-08-20 | 88 | 2055 |
| 2020-08-21 | 90 | 2098 |

| | | |
|---|---|---|
| 2020-08-24 | 93 | 2154 |
| 2020-08-25 | 94 | 2211 |
| 2020-08-26 | 96 | 2275 |
| 2020-08-27 | 100 | 2298 |
| 2020-08-28 | 100 | 2356 |
| 2020-08-31 | 102 | 2418 |
| 2020-09-01 | 103 | 2449 |
| 2020-09-02 | 104 | 2498 |
| 2020-09-04 | 106 | 2613 |
| 2020-09-08 | 106 | 2656 |
| 2020-09-09 | 109 | 2677 |
| 2020-09-10 | 110 | 2704 |
| 2020-09-11 | 111 | 2721 |
| 2020-09-14 | 114 | 2730 |
| 2020-09-15 | 114 | 2801 |
| 2020-09-16 | 116 | 2844 |
| 2020-09-17 | 120 | 2901 |
| 2020-09-18 | 120 | 2950 |
| 2020-09-21 | 122 | 3000 |
| 2020-09-22 | 123 | 3036 |
| 2020-09-23 | 123 | 3070 |
| 2020-09-24 | 126 | 3093 |
| 2020-09-25 | 127 | 3138 |
| 2020-09-28 | 133 | 3157 |
| 2020-09-29 | 133 | 3193 |
| 2020-09-30 | 134 | 3241 |
| 2020-10-02 | 137 | 3307 |
| 2020-10-05 | 137 | 3340 |
| 2020-10-06 | 138 | 3361 |
| 2020-10-07 | 138 | 3395 |
| 2020-10-09 | 142 | 3434 |
| 2020-10-13 | 144 | 3453 |
| 2020-10-14 | 146 | 3473 |
| 2020-10-15 | 146 | 3529 |
| 2020-10-16 | 147 | 3564 |
| 2020-10-19 | 149 | 3582 |
| 2020-10-20 | 149 | 3600 |
| 2020-10-21 | 151 | 3629 |
| 2020-10-22 | 151 | 3644 |
| 2020-10-23 | 152 | 3674 |
| 2020-10-26 | 153 | 3708 |
| 2020-10-27 | 153 | 3736 |

| | | |
|---|---|---|
| 2020-10-28 | 156 | 3817 |
| 2020-10-29 | 158 | 3858 |
| 2020-10-30 | 158 | 3878 |
| 2020-11-03 | 159 | 3898 |
| 2020-11-04 | 160 | 3916 |
| 2020-11-05 | 161 | 3939 |
| 2020-11-06 | 161 | 3988 |
| 2020-11-09 | 162 | 4005 |
| 2020-11-10 | 162 | 4049 |
| 2020-11-11 | 162 | 4070 |
| 2020-11-12 | 162 | 4142 |
| 2020-11-13 | 162 | 4179 |
| 2020-11-16 | 162 | 4208 |
| 2020-11-17 | 162 | 4227 |
| 2020-11-18 | 164 | 4303 |
| 2020-11-19 | 166 | 4348 |
| 2020-11-23 | 169 | 4391 |
| 2020-11-25 | 173 | 4543 |
| 2020-11-27 | 175 | 4703 |
| 2020-11-30 | 177 | 4820 |
| 2020-12-02 | 177 | 4962 |
| 2020-12-04 | 179 | 5099 |
| 2020-12-07 | 179 | 5193 |
| 2020-12-09 | 181 | 5358 |
| 2020-12-11 | 185 | 5498 |
| 2020-12-14 | 189 | 5630 |
| 2020-12-16 | 192 | 5868 |
| 2020-12-18 | 196 | 6032 |
| 2020-12-21 | 200 | 6174 |
| 2020-12-23 | 204 | 6391 |
| 2020-12-28 | 214 | 6532 |
| 2020-12-30 | 221 | 6882 |
| 2021-01-04 | 231 | 7058 |
| 2021-01-05 | 232 | 7238 |
| 2021-01-06 | 233 | 7350 |
| 2021-01-07 | 233 | 7519 |
| 2021-01-08 | 233 | 7583 |
| 2021-01-11 | 240 | 7633 |
| 2021-01-12 | 244 | 7833 |
| 2021-01-13 | 248 | 7949 |
| 2021-01-14 | 249 | 8055 |
| 2021-01-15 | 250 | 8163 |

| | | |
|---|---|---|
| 2021-01-18 | 250 | 8283 |
| 2021-01-19 | 252 | 8500 |
| 2021-01-20 | 254 | 8579 |
| 2021-01-21 | 255 | 8644 |
| 2021-01-22 | 255 | 8759 |
| 2021-01-25 | 261 | 8849 |
| 2021-01-26 | 262 | 8941 |
| 2021-01-27 | 263 | 9049 |
| 2021-01-28 | 267 | 9144 |
| 2021-01-29 | 268 | 9228 |
| 2021-02-01 | 275 | 9342 |
| 2021-02-02 | 277 | 9442 |
| 2021-02-03 | 278 | 9488 |
| 2021-02-04 | 279 | 9522 |
| 2021-02-05 | 279 | 9586 |
| 2021-02-06 | 280 | 9649 |
| 2021-02-08 | 280 | 9649 |
| 2021-02-09 | 282 | 9697 |
| 2021-02-10 | 284 | 9729 |
| 2021-02-11 | 287 | 9753 |
| 2021-02-12 | 289 | 9799 |
| 2021-02-15 | 291 | 9832 |
| 2021-02-17 | 293 | 9870 |
| 2021-02-18 | 296 | 9901 |
| 2021-02-19 | 297 | 9936 |
| 2021-02-22 | 297 | 9997 |
| 2021-02-23 | 299 | 10081 |
| 2021-02-24 | 301 | 10135 |
| 2021-02-25 | 303 | 10169 |
| 2021-02-26 | 304 | 10201 |
| 2021-03-01 | 306 | 10243 |
| 2021-03-02 | 306 | 10329 |
| 2021-03-03 | 307 | 10367 |
| 2021-03-04 | 308 | 10431 |
| 2021-03-05 | 309 | 10503 |
| 2021-03-08 | 312 | 10568 |
| 2021-03-09 | 312 | 10662 |
| 2021-03-10 | 314 | 10751 |
| 2021-03-11 | 315 | 10857 |
| 2021-03-12 | 315 | 10902 |
| 2021-03-15 | 318 | 10983 |
| 2021-03-16 | 318 | 11083 |

| | | |
|---|---|---|
| 2021-03-17 | 318 | 11131 |
| 2021-03-18 | 318 | 11189 |
| 2021-03-19 | 320 | 11248 |
| 2021-03-22 | 324 | 11283 |
| 2021-03-23 | 327 | 11384 |
| 2021-03-24 | 330 | 11493 |
| 2021-03-25 | 334 | 11613 |
| 2021-03-26 | 339 | 11683 |
| 2021-03-29 | 348 | 11821 |
| 2021-03-30 | 354 | 11945 |
| 2021-03-31 | 358 | 12050 |
| 2021-04-01 | 361 | 12172 |
| 2021-04-05 | 376 | 12400 |
| 2021-04-06 | 381 | 12616 |
| 2021-04-07 | 383 | 12872 |
| 2021-04-08 | 387 | 13137 |
| 2021-04-09 | 389 | 13303 |
| 2021-04-12 | 404 | 13545 |
| 2021-04-13 | 413 | 13694 |
| 2021-04-14 | 422 | 13841 |
| 2021-04-15 | 428 | 13981 |
| 2021-04-16 | 436 | 14140 |
| 2021-04-19 | 451 | 14300 |
| 2021-04-20 | 454 | 14598 |
| 2021-04-21 | 459 | 14792 |
| 2021-04-22 | 466 | 14983 |
| 2021-04-23 | 470 | 15141 |
| 2021-04-26 | 477 | 15299 |
| 2021-04-27 | 485 | 15437 |
| 2021-04-28 | 491 | 15533 |
| 2021-04-29 | 494 | 15630 |
| 2021-04-30 | 499 | 15735 |
| 2021-05-03 | 514 | 15873 |
| 2021-05-04 | 520 | 16056 |
| 2021-05-05 | 525 | 16167 |
| 2021-05-06 | 534 | 16280 |
| 2021-05-07 | 540 | 16374 |
| 2021-05-10 | 548 | 16479 |
| 2021-05-11 | 552 | 16669 |
| 2021-05-12 | 558 | 16877 |
| 2021-05-13 | 563 | 16964 |
| 2021-05-14 | 566 | 17044 |

| | | |
|---|---|---|
| 2021-05-17 | 575 | 17100 |
| 2021-05-18 | 578 | 17170 |
| 2021-05-19 | 587 | 17256 |
| 2021-05-20 | 588 | 17355 |
| 2021-05-21 | 590 | 17430 |
| 2021-05-24 | 597 | 17524 |
| 2021-05-25 | 602 | 17598 |
| 2021-05-26 | 605 | 17756 |
| 2021-05-27 | 608 | 17947 |
| 2021-05-28 | 611 | 18048 |
| 2021-05-31 | 619 | 18191 |
| 2021-06-01 | 622 | 18317 |
| 2021-06-02 | 622 | 18482 |
| 2021-06-07 | 630 | 18584 |
| 2021-06-08 | 631 | 18744 |
| 2021-06-09 | 632 | 18959 |
| 2021-06-10 | 642 | 19021 |
| 2021-06-11 | 643 | 19164 |
| 2021-06-14 | 648 | 19343 |
| 2021-06-15 | 652 | 19417 |
| 2021-06-16 | 654 | 19505 |
| 2021-06-17 | 656 | 19549 |
| 2021-06-18 | 658 | 19614 |
| 2021-06-21 | 663 | 19763 |
| 2021-06-22 | 665 | 19854 |
| 2021-06-23 | 669 | 19919 |
| 2021-06-24 | 671 | 19967 |
| 2021-06-25 | 673 | 20008 |
| 2021-06-28 | 675 | 20154 |
| 2021-06-29 | 680 | 20199 |
| 2021-06-30 | 680 | 20286 |
| 2021-07-01 | 680 | 20378 |
| 2021-07-02 | 681 | 20438 |
| 2021-07-05 | 687 | 20551 |
| 2021-07-06 | 690 | 20618 |
| 2021-07-07 | 690 | 20680 |
| 2021-07-08 | 691 | 20749 |
| 2021-07-09 | 693 | 20832 |
| 2021-07-12 | 700 | 20840 |
| 2021-07-13 | 702 | 20905 |
| 2021-07-14 | 705 | 21084 |
| 2021-07-15 | 708 | 21188 |

| | | |
|---|---|---|
| 2021-07-16 | 712 | 21260 |
| 2021-07-19 | 715 | 21317 |
| 2021-07-20 | 719 | 21438 |
| 2021-07-21 | 720 | 21553 |
| 2021-07-22 | 720 | 21637 |
| 2021-07-23 | 724 | 21746 |
| 2021-07-26 | 732 | 21947 |
| 2021-07-27 | 732 | 22046 |
| 2021-07-28 | 734 | 22137 |
| 2021-07-29 | 736 | 22229 |
| 2021-07-30 | 739 | 22279 |
| 2021-08-02 | 743 | 22396 |
| 2021-08-03 | 746 | 22495 |
| 2021-08-04 | 750 | 22521 |
| 2021-08-05 | 750 | 22571 |
| 2021-08-06 | 752 | 22654 |
| 2021-08-09 | 755 | 22832 |
| 2021-08-10 | 758 | 22982 |
| 2021-08-11 | 763 | 23060 |
| 2021-08-12 | 765 | 23141 |
| 2021-08-13 | 766 | 23186 |
| 2021-08-16 | 771 | 23339 |
| 2021-08-17 | 776 | 23464 |
| 2021-08-18 | 777 | 23527 |
| 2021-08-19 | 778 | 23600 |
| 2021-08-20 | 781 | 23642 |
| 2021-08-23 | 788 | 23712 |
| 2021-08-24 | 790 | 23812 |
| 2021-08-25 | 793 | 23927 |
| 2021-08-26 | 794 | 24059 |
| 2021-08-27 | 798 | 24100 |
| 2021-08-30 | 806 | 24341 |
| 2021-08-31 | 807 | 24450 |
| 2021-09-01 | 808 | 24559 |
| 2021-09-02 | 808 | 24619 |
| 2021-09-03 | 809 | 24663 |
| 2021-09-08 | 819 | 24733 |
| 2021-09-09 | 821 | 24826 |
| 2021-09-10 | 824 | 24881 |
| 2021-09-13 | 831 | 24961 |
| 2021-09-14 | 832 | 25039 |
| 2021-09-15 | 832 | 25070 |

| | | |
|---|---|---|
| 2021-09-16 | 833 | 25093 |
| 2021-09-17 | 834 | 25125 |
| 2021-09-20 | 838 | 25185 |
| 2021-09-21 | 842 | 25215 |
| 2021-09-22 | 842 | 25235 |
| 2021-09-23 | 842 | 25308 |
| 2021-09-24 | 842 | 25334 |
| 2021-09-27 | 843 | 25357 |
| 2021-09-28 | 844 | 25426 |
| 2021-09-29 | 849 | 25451 |
| 2021-10-01 | 849 | 25479 |
| 2021-10-04 | 849 | 25497 |
| 2021-10-05 | 850 | 25526 |
| 2021-10-07 | 850 | 25556 |
| 2021-10-08 | 852 | 25573 |
| 2021-10-13 | 855 | 25593 |
| 2021-10-14 | 855 | 25604 |
| 2021-10-15 | 856 | 25616 |
| 2021-10-18 | 857 | 25629 |
| 2021-10-19 | 857 | 25633 |
| 2021-10-20 | 860 | 25642 |
| 2021-10-21 | 860 | 25673 |
| 2021-10-22 | 860 | 25686 |
| 2021-10-26 | 860 | 25698 |
| 2021-10-27 | 860 | 25703 |
| 2021-10-28 | 860 | 25706 |
| 2021-10-29 | 861 | 25709 |
| 2021-11-03 | 862 | 25723 |
| 2021-11-04 | 862 | 25730 |
| 2021-11-05 | 862 | 25736 |
| 2021-11-08 | 863 | 25747 |
| 2021-11-09 | 863 | 25751 |
| 2021-11-10 | 864 | 25753 |
| 2021-11-11 | 864 | 25759 |
| 2021-11-12 | 864 | 25765 |
| 2021-11-16 | 864 | 25772 |
| 2021-11-17 | 864 | 25777 |
| 2021-11-18 | 864 | 25779 |
| 2021-11-19 | 864 | 25781 |
| 2021-11-22 | 864 | 25784 |
| 2021-11-23 | 864 | 25784 |
| 2021-11-24 | 864 | 25794 |

| | | |
|---|---|---|
| 2021-11-25 | 864 | 25796 |
| 2021-11-26 | 864 | 25798 |
| 2021-11-29 | 865 | 25804 |
| 2021-11-30 | 865 | 25807 |
| 2021-12-01 | 865 | 25807 |
| 2021-12-02 | 865 | 25809 |
| 2021-12-03 | 865 | 25812 |
| 2021-12-06 | 865 | 25819 |
| 2021-12-07 | 865 | 25826 |
| 2021-12-08 | 865 | 25827 |
| 2021-12-09 | 865 | 25827 |
| 2021-12-10 | 866 | 25829 |
| 2021-12-28 | 868 | 25847 |
| 2021-12-29 | 868 | 25854 |
| 2021-12-30 | 868 | 25861 |
| 2022-01-03 | 868 | 25884 |
| 2022-01-04 | 868 | 25941 |
| 2022-01-05 | 868 | 25994 |
| 2022-01-06 | 868 | 26055 |
| 2022-01-07 | 868 | 26109 |
| 2022-01-11 | 868 | 26130 |
| 2022-01-12 | 868 | 26170 |
| 2022-01-13 | 868 | 26261 |
| 2022-01-14 | 868 | 26328 |
| 2022-01-18 | 870 | 26450 |
| 2022-01-20 | 870 | 26632 |
| 2022-01-21 | 870 | 26780 |
| 2022-01-24 | 870 | 26930 |
| 2022-01-25 | 871 | 27224 |
| 2022-01-26 | 873 | 27330 |
| 2022-01-27 | 873 | 27489 |
| 2022-01-31 | 876 | 27612 |
| 2022-02-01 | 876 | 27679 |
| 2022-02-02 | 878 | 27784 |
| 2022-02-04 | 883 | 27891 |
| 2022-02-08 | 888 | 27944 |
| 2022-02-09 | 891 | 28031 |
| 2022-02-10 | 894 | 28132 |
| 2022-02-11 | 895 | 28280 |
| 2022-02-14 | 896 | 28336 |
| 2022-02-15 | 903 | 28451 |
| 2022-02-16 | 904 | 28473 |

| | | |
|---|---|---|
| 2022-02-17 | 904 | 28542 |
| 2022-02-18 | 905 | 28607 |
| 2022-02-22 | 909 | 28722 |
| 2022-02-23 | 911 | 28777 |
| 2022-02-24 | 912 | 28820 |
| 2022-02-25 | 912 | 28860 |
| 2022-03-02 | 917 | 28939 |
| 2022-03-03 | 918 | 29026 |
| 2022-03-04 | 918 | 29127 |
| 2022-03-07 | 920 | 29197 |
| 2022-03-09 | 923 | 29423 |
| 2022-03-10 | 923 | 29497 |
| 2022-03-11 | 924 | 29536 |
| 2022-03-14 | 925 | 29594 |
| 2022-03-15 | 925 | 29665 |
| 2022-03-16 | 925 | 29718 |
| 2022-03-17 | 927 | 29827 |
| 2022-03-18 | 927 | 29833 |
| 2022-03-21 | 928 | 29865 |
| 2022-03-22 | 928 | 29886 |
| 2022-03-23 | 928 | 29901 |
| 2022-03-24 | 928 | 29901 |
| 2022-03-25 | 928 | 29924 |
| 2022-03-29 | 928 | 30028 |
| 2022-03-30 | 928 | 30080 |
| 2022-04-01 | 928 | 30163 |

### 2 Database table of accumulated cases per groups

| | **Sex %** | | **Rural area %** | | | | | | | | **Age group %** | | | | | | | | |
|---|---|---|---|---|---|---|---|---|---|---|---|---|---|---|---|---|---|---|---|
| **Date** | Male | Female | Amparo | Campo do coelho | Riograndina | São Pedro | Mury | Lumiar | Total rural | <1 | 1-4 | 5-9 | 10-14 | 15-19 | 20-29 | 30-39 | 40-49 | 50-59 | ≥60 |
| 2020-06-26 | 49.9 | 50.1 | 1.2 | 4.3 | 1 | 0.4 | 2.7 | 0.6 | 10.2 | | 0.8 | 0.8 | | 0.4 | 8.5 | 20 | 21 | 16.9 | 31.5 |
| 2020-07-03 | 49.9 | 50.1 | 1.2 | 4.2 | 1.7 | 0.3 | 2.4 | 0.7 | 10.5 | | 0.7 | 0.7 | | 0.3 | 8.4 | 19.7 | 23.2 | 16.2 | 30.7 |
| 2020-07-10 | 50.1 | 49.9 | 1.4 | 4.3 | 1.9 | 0.4 | 2.2 | 0.7 | 10.9 | | 0.7 | 0.6 | | 0.3 | 9.6 | 19.5 | 23 | 17.2 | 29.2 |
| 2020-07-17 | 49.4 | 50.6 | 1.3 | 4 | 2.6 | 0.5 | 2.1 | 0.6 | 11.1 | | 0.6 | 0.7 | | 0.2 | 9.1 | 19.4 | 23.3 | 18.1 | 28.5 |

| | | | | | | | | | | | | | | | | | | | |
|---|---|---|---|---|---|---|---|---|---|---|---|---|---|---|---|---|---|---|---|
| 2020-07-24 | 48.8 | 51.2 | 1.9 | 3.7 | 2.3 | 0.5 | 2 | 0.6 | 11 | | 0.7 | 0.8 | 0.3 | 0.6 | 9.8 | 20.2 | 22 | 17.7 | 27.8 |
| 2020-07-31 | 48.4 | 51.6 | 2.1 | 3.4 | 2.2 | 0.6 | 2.2 | 0.7 | 11.2 | | 0.8 | 0.7 | 0.3 | 0.9 | 11.1 | 20 | 23 | 16.8 | 26..3 |
| 2020-08-07 | 47.9 | 52.1 | 1.8 | 3 | 2.1 | 0.6 | 2.1 | 0.8 | 10.4 | | 0.9 | 0.7 | 0.4 | 1.2 | 12.3 | 20.7 | 22.3 | 16.2 | 25.4 |
| 2020-08-14 | 47.4 | 52.6 | 2 | 2.7 | 2.2 | 0.6 | 2.2 | 0.7 | 10.4 | | 0.9 | 0.7 | 0.4 | 1.2 | 12.8 | 20.5 | 22.6 | 16.1 | 24.7 |
| 2020-08-21 | .47.1 | 52.9 | 1.9 | 2.7 | 2.2 | 0.6 | 2.4 | 0.7 | 10.5 | | 0.9 | 0.8 | 0.4 | 1.3 | 13.2 | 20.1 | 22.1 | 165 | 24.7 |
| 2020-08-28 | 47 | 53 | 2 | 2.7 | 2.1 | 0.5 | 2.7 | 0.7 | 10.7 | | 0.9 | 0.9 | 0.6 | 1.3 | 13.1 | 20.8 | 22 | 16.7 | 23.7 |
| 2020-09-04 | 46.8 | 53.2 | 1.9 | 2.5 | 2.1 | 0.5 | 2.7 | 0.6 | 10.3 | | 1 | 0.9 | 0.6 | 1.4 | 13.6 | 20.7 | 21.8 | 16.8 | 23.2 |
| 2020-09-11 | 46.6 | 53.4 | 1.8 | 2.5 | 2.1 | 0.5 | 2.6 | 0.6 | 10.1 | | 1 | 1 | 0.7 | 1.4 | 13.6 | 20.7 | 21.6 | 16.9 | 23 |
| 2020-09-18 | 46.5 | 53.5 | 2 | 2.4 | 2.2 | 0.5 | 2.6 | 0.5 | 10.2 | | 1 | 1 | 0.7 | 1.6 | 13.9 | 20.4 | 21.4 | 16.9 | 23 |
| 2020-09-25 | 46.2 | 53.8 | 2.1 | 2.4 | 2.4 | 0.4 | 2.6 | 0.6 | 10.5 | | 1 | 1 | 0.7 | 1.8 | 14 | 20.3 | 21.2 | 16.9 | 23.1 |
| 2020-10-02 | 45.9 | 54.1 | 2.1 | 2.5 | 2.4 | 0.5 | 2.6 | 0.6 | 10.7 | | 1 | 0.9 | 0.8 | 1.7 | 14 | 20.1 | 21.4 | 17 | 23 |
| 2020-10-09 | 45.8 | 54.2 | 2.2 | 2.5 | 2.3 | 0.6 | 2.4 | 0.6 | 10.6 | | 1 | 0.9 | 0.8 | 1.6 | 14.1 | 20.1 | 21.5 | 17.1 | 22.8 |
| 2020-10-16 | 46 | 54 | 2.2 | 2.4 | 2.3 | 0.6 | 2.5 | 0.6 | 10.6 | | 1 | 0.9 | 0.8 | 1.7 | 14.1 | 20.1 | 21.4 | 17.1 | 22.8 |
| 2020-10-23 | 45.9 | 54.1 | 2.3 | 2.4 | 2.3 | 0.6 | 2.4 | 0.7 | 10.7 | | 1 | 1 | 0.8 | 1.7 | 14.1 | 20.1 | 21.3 | 17.1 | 22.8 |
| 2020-10-30 | 46 | 54 | 2.2 | 2.3 | 2.3 | 0.6 | 2.4 | 0.7 | 10.5 | | 1 | 1 | 0.8 | 1.8 | 14.3 | 20.3 | 21.2 | 17.1 | 22.5 |
| 2020-11-06 | 45.9 | 54.1 | 2.3 | 2.3 | 2.3 | 0.6 | 2.4 | 0.8 | 10.7 | | 1 | 1 | 0.8 | 1.8 | 14.3 | 20.2 | 21.2 | 16.9 | 22.7 |
| 2020-11-13 | 45.8 | 54.2 | 2.3 | 2.2 | 2.3 | 0.6 | 2.4 | 0.8 | 10.6 | | 1 | 1 | 0.8 | 1.8 | 14.5 | 20.2 | 21.5 | 16.6 | 22.5 |
| 2020-11-18 | 45.9 | 54.1 | 2.3 | 2.2 | 2.3 | 0.6 | 2.5 | 0.8 | 10.7 | | 1 | 0.9 | 1 | 1.9 | 14.5 | 20.3 | 21.5 | 16.5 | 22.4 |
| 2020-11-27 | 45.8 | 54.2 | 2.2 | 2.2 | 2.3 | 0.6 | 2.4 | 0.9 | 10.6 | | 0.9 | 0.9 | 0.9 | 1.9 | 15.1 | 20.3 | 21.4 | 16.3 | 22.1 |
| 2020-12-04 | 45.7 | 54.3 | 2.2 | 2.3 | 2.2 | 0.6 | 2.4 | 0.9 | 10.6 | | 0.9 | 0.9 | 0.9 | 2 | 15.1 | 20.5 | 21.2 | 16.2 | 22.1 |
| 2020-12-11 | 45.5 | 54.5 | 2.1 | 2.2 | 2.3 | 0.6 | 2.4 | 0.9 | 10.5 | | 0.9 | 0.9 | 0.9 | 1.9 | 14.9 | 20.5 | 21.3 | 16.3 | 22.3 |
| 2020-12-18 | 45.5 | 54.5 | 2.1 | 2.3 | 2.2 | 0.6 | 2.4 | 1 | 10.6 | 0.2 | 0.9 | 0.9 | 0.9 | 2.1 | 15.2 | 20.5 | 21 | 16.3 | 22.1 |
| 2020-12-23 | 45.3 | 54.7 | 2 | 2.2 | 2.2 | 0.6 | 2.4 | 1.1 | 10.5 | 0.2 | 0.9 | 0.8 | 0.9 | 2.1 | 15.1 | 20.5 | 20.9 | 16.5 | 22.1 |
| 2020-12-30 | 45.5 | 54.5 | 2.1 | 2.2 | 2.2 | 0.6 | 2.4 | 1.1 | 10.6 | 0.2 | 0.8 | 0.8 | 0.9 | 2.3 | 14.8 | 20.6 | 21.1 | 16.7 | 21.9 |
| 2021-01-08 | 45.1 | 54.9 | 2.1 | 2.1 | 2.1 | 0.6 | 2.3 | 1.1 | 10.3 | 0.2 | 0.8 | 0.8 | 1 | 2.4 | 14.9 | 20.7 | 20.9 | 16.7 | 21.7 |
| 2021-01-15 | 45.1 | 54.9 | 2 | 2.1 | 2 | 0.6 | 2.3 | 1.2 | 10.2 | 0.2 | 0.7 | 0.8 | 1 | 2.5 | 14.9 | 20.6 | 21.1 | 16.5 | 21.6 |
| 2021-01-22 | 45.1 | 54.9 | 2.1 | 2.1 | 2.1 | 0.6 | 2.3 | 1.2 | 10.4 | 0.2 | 0.8 | 0.8 | 1 | 2.6 | 14.9 | 20.7 | 20.8 | 16.6 | 21.7 |
| 2021-01-29 | 45 | 55 | 2.1 | 2.2 | 2.1 | 0.6 | 2.2 | 1.3 | 10.5 | 0.2 | 0.8 | 0.8 | 1 | 2.7 | 14.7 | 20.8 | 20.8 | 16.5 | 21.7 |
| 2021-02-05 | 45 | 55 | 2.1 | 2.1 | 2.1 | 0.5 | 2.2 | 1.3 | 10.3 | 0.2 | 0.8 | 0.8 | 1 | 2.7 | 14.8 | 20.6 | 20.7 | 16.5 | 22 |
| 2021-02-12 | 44.9 | 55.1 | 2.1 | 2.1 | 2.1 | 0.5 | 2.2 | 1.3 | 10.3 | 0.2 | 0.7 | 0.8 | 1 | 2.7 | 14.8 | 20.6 | 20.7 | 16.5 | 21.9 |
| 2021-02-19 | 45 | 55 | 2.1 | 2.1 | 2.1 | 0.5 | 2.2 | 1.3 | 10.3 | 0.2 | 0.7 | 0.8 | 1 | 2.7 | 14.9 | 20.5 | 20.6 | 16.6 | 22 |
| 2021-02-26 | 44.8 | 55.2 | 2.1 | 2.1 | 2.1 | 0.5 | 2.1 | 1.3 | 10.2 | 0.2 | 0.8 | 0.8 | 1 | 2.7 | 14.9 | 20.5 | 20.6 | 16.6 | 21.9 |
| 2021-03-05 | 44.9 | 55.1 | 2.1 | 2.1 | 2.2 | 0.5 | 2.2 | 1.3 | 10.4 | 0.2 | 0.8 | 0.8 | 1 | 2.7 | 15.1 | 20.4 | 20.5 | 16.7 | 21.8 |
| 2021-03-12 | 45 | 55 | 2.1 | 2.1 | 2.2 | 0.5 | 2.2 | 1.3 | 10.4 | 0.2 | 0.8 | 0.8 | 1 | 2.7 | 15.1 | 20.4 | 20.4 | 16.8 | 21.7 |
| 2021-03-19 | 45.1 | 54.9 | 2.1 | 2.1 | 2.3 | 0.5 | 2.1 | 1.3 | 10.4 | 0.2 | 0.8 | 0.8 | 1 | 2.8 | 15.1 | 20.5 | 20.4 | 16.7 | 21.7 |
| 2021-03-26 | 45.1 | 54.9 | 2.1 | 2.1 | 2.3 | 0.5 | 2.1 | 1.3 | 10.4 | 0.2 | 0.8 | 0.8 | 1.1 | 2.8 | 15.2 | 20.4 | 20.4 | 16.6 | 21.8 |
| 2021-04-01 | 45.1 | 54.9 | 2.2 | 2.1 | 2.3 | 0.5 | 2.1 | 1.3 | 10.5 | 0.3 | 0.8 | 0.8 | 1.1 | 2.8 | 15.3 | 20.2 | 20.3 | 16.6 | 21.7 |
| 2021-04-08 | 45.1 | 54.9 | 2.3 | 2.1 | 2.3 | 0.5 | 2 | 1.4 | 10.6 | 0.3 | 0.8 | 0.8 | 1.1 | 2.9 | 15.6 | 20.3 | 20.2 | 16.6 | 21.4 |
| 2021-04-15 | 45.4 | 54.6 | 2.4 | 2.1 | 2.3 | 0.5 | 2.1 | 1.4 | 10.8 | 0.3 | 0.7 | 0.8 | 1.1 | 3 | 15.6 | 20.1 | 20.3 | 16.7 | 21.4 |
| 2021-04-22 | 45.4 | 54.6 | 2.5 | 2.1 | 2.4 | 0.5 | 2 | 1.5 | 11 | 0.4 | 0.7 | 0.8 | 1.2 | 3.1 | 15.6 | 20 | 20.4 | 16.6 | 21.2 |
| 2021-04-29 | 45.3 | 54.7 | 2.5 | 2.2 | 2.4 | 0.5 | 2 | 1.5 | 11.1 | 0.4 | 0.7 | 0.8 | 1.2 | 3.2 | 15.6 | 20 | 20.3 | 16.6 | 21.2 |
| 2021-05-06 | 45.4 | 54.6 | 2.5 | 2.2 | 2.4 | 0.4 | 2 | 1.4 | 10.9 | 0.4 | 0.7 | 0.8 | 1.2 | 3.2 | 15.6 | 19.9 | 20.4 | 16.7 | 21.2 |

| | | | | | | | | | | | | | | | | | | | |
|---|---|---|---|---|---|---|---|---|---|---|---|---|---|---|---|---|---|---|---|
| 2021-05-13 | 45.5 | 54.4 | 2.5 | 2.2 | 2.4 | 0.5 | 1.9 | 1.5 | 11 | 0.4 | 0.7 | 0.8 | 1.2 | 3.2 | 15.7 | 19.9 | 20.4 | 16.7 | 21 |
| 2021-05-20 | 45.6 | 54.4 | 2.5 | 2.2 | 2.4 | 0.5 | 1.9 | 1.6 | 11.1 | 0.4 | 0.7 | 0.8 | 1.2 | 3.2 | 15.8 | 19.9 | 20.3 | 16.7 | 21 |
| 2021-05-27 | 45.5 | 54.5 | 2.5 | 2.3 | 2.4 | 0.5 | 1.9 | 1.6 | 11.2 | 0.4 | 0.7 | 0.8 | 1.2 | 3.3 | 15.9 | 19.9 | 20.3 | 16.6 | 20.8 |
| 2021-06-02 | 45.5 | 54.5 | 2.5 | 2.3 | 2.4 | 0.5 | 1.9 | 1.7 | 11.3 | 0.5 | 0.7 | 0.9 | 1.2 | 3.3 | 15.9 | 19.9 | 20.3 | 16.7 | 20.6 |
| 2021-06-10 | 45.4 | 54.6 | 2.4 | 2.3 | 2.4 | 0.5 | 1.9 | 1.6 | 11.1 | 0.5 | 0.7 | 0.9 | 1.3 | 3.4 | 16 | 20 | 20.3 | 16.6 | 20.4 |
| 2021-06-17 | 45.5 | 54.5 | 2.4 | 2.4 | 2.5 | 0.5 | 1.9 | 1.7 | 11.4 | 0.5 | 0.7 | 0.9 | 1.3 | 3.4 | 16.1 | 20.1 | 20.3 | 16.6 | 20.2 |
| 2021-06-24 | 45.5 | 54.5 | 2.4 | 2.3 | 2.6 | 0.5 | 1.9 | 1.7 | 11.4 | 0.5 | 0.7 | 0.9 | 1.3 | 3.4 | 16.2 | 20.1 | 20.3 | 16.6 | 20 |
| 2021-07-01 | 45.5 | 54.4 | 2.3 | 2.3 | 2.6 | 0.5 | 1.8 | 1.8 | 11.3 | 0.5 | 0.7 | 0.9 | 1.3 | 3.5 | 16.2 | 20.2 | 20.4 | 16.5 | 19.9 |
| 2021-07-08 | 45.5 | 54.5 | 2.3 | 2.3 | 2.6 | 0.5 | 1.8 | 1.8 | 11.3 | 0.5 | 0.7 | 0.9 | 1.3 | 3.5 | 16.3 | 20.2 | 20.3 | 16.5 | 19.8 |
| 2021-07-15 | 45.5 | 54.5 | 2.3 | 2.3 | 2.6 | 0.6 | 1.8 | 1.8 | 11.4 | 0.4 | 0.8 | 0.8 | 1.3 | 3.1 | 16 | 20 | 20.5 | 16.7 | 20.5 |
| 2021-07-22 | 45.7 | 54.3 | 2.3 | 2.3 | 2.6 | 0.6 | 1.9 | 1.8 | 11.5 | 0.4 | 0.7 | 0.8 | 1.3 | 3.2 | 16.1 | 20 | 20.5 | 16.6 | 20.4 |
| 2021-07-29 | 45.7 | 54.3 | 2.2 | 2.2 | 2.6 | 0.7 | 1.9 | 1.9 | 11.5 | 0.4 | 0.7 | 0.8 | 1.4 | 3.2 | 16.2 | 20.1 | 20.4 | 16.5 | 20.3 |
| 2021-08-05 | 45.7 | 54.3 | 2.2 | 2.2 | 2.6 | 0.7 | 1.9 | 2 | 11.6 | 0.4 | 0.7 | 0.8 | 1.4 | 3.2 | 16.2 | 20.1 | 20.4 | 16.5 | 20.2 |
| 2021-08-12 | 45.7 | 54.3 | 2.2 | 2.2 | 2.6 | 0.7 | 1.9 | 1.9 | 11.5 | 0.4 | 0.7 | 0.9 | 1.5 | 3.2 | 16.4 | 20.1 | 20.2 | 16.4 | 20.1 |
| 2021-08-19 | 45.6 | 54.3 | 2.2 | 2.2 | 2.6 | 0.7 | 1.9 | 1.9 | 11.5 | 0.4 | 0.7 | 0.9 | 1.5 | 3.3 | 16.4 | 20.1 | 20.2 | 16.3 | 20.1 |
| 2021-08-26 | 45.7 | 54.3 | 2.3 | 2.2 | 2.6 | 0.7 | 1.9 | 1.9 | 11.6 | 0.4 | 0.7 | 0.9 | 1.5 | 3.3 | 16.5 | 20.1 | 20.1 | 16.3 | 20.1 |
| 2021-09-02 | 45.7 | 54.2 | 2.3 | 2.2 | 2.6 | 0.7 | 2 | 1.9 | 11.7 | 0.4 | 0.8 | 0.9 | 1.6 | 3.4 | 16.6 | 20 | 20.1 | 16.1 | 20 |
| 2021-09-09 | 45.7 | 54.3 | 2.3 | 2.2 | 2.5 | 0.7 | 2 | 1.9 | 11.6 | 0.4 | 0.8 | 0.9 | 1.6 | 3.4 | 16.7 | 20 | 20.1 | 16.1 | 20 |
| 2021-09-16 | 45.7 | 54.3 | 2.3 | 2.2 | 2.5 | 0.6 | 2 | 1.9 | 11.5 | 0.4 | 0.8 | 0.9 | 1.6 | 3.5 | 16.6 | 19.9 | 20 | 16.1 | 20 |
| 2021-09-23 | 45.8 | 54.2 | 2.3 | 2.2 | 2.5 | 0.6 | 2 | 1.9 | 11.5 | 0.4 | 0.8 | 1 | 1.6 | 3.5 | 16.6 | 19.9 | 20.1 | 16.1 | 20.1 |
| 2021-09-30 | 45.8 | 54.2 | 2.3 | 2.2 | 2.5 | 0.6 | 2 | 1.9 | 11.5 | 0.4 | 0.8 | 1 | 1.6 | 3.5 | 16.6 | 19.9 | 20.1 | 16 | 20.1 |
| 2021-10-14 | 45.7 | 54.2 | 2.3 | 2.2 | 2.5 | 0.6 | 2 | 1.9 | 11.5 | 0.4 | 0.8 | 1 | 1.6 | 3.5 | 16.6 | 19.9 | 20.1 | 16 | 20.1 |
| 2021-10-21 | 45.7 | 54.3 | 2.3 | 2.2 | 2.5 | 0.6 | 2 | 1.9 | 11.5 | 0.4 | 0.8 | 1 | 1.6 | 3.5 | 16.6 | 19.9 | 20.1 | 16 | 20.2 |
| 2021-10-28 | 45.7 | 54.3 | 2.3 | 2.2 | 2.5 | 0.6 | 2 | 1.9 | 11.5 | 0.4 | 0.8 | 1 | 1.6 | 3.5 | 16.6 | 19.8 | 20.1 | 16 | 20.2 |
| 2021-11-04 | 45.7 | 54.3 | 2.3 | 2.2 | 2.5 | 0.6 | 2 | 1.9 | 11.5 | 0.4 | 0.8 | 1 | 1.6 | 3.5 | 16.6 | 19.9 | 20.1 | 16 | 20.2 |
| 2021-11-11 | 45.7 | 54.3 | 2.3 | 2.2 | 2.5 | 0.6 | 2 | 1.9 | 11.5 | 0.4 | 0.8 | 1 | 1.6 | 3.5 | 16.6 | 19.8 | 20.1 | 16 | 20.2 |
| 2021-11-18 | 45.7 | 54.3 | 2.3 | 2.2 | 2.5 | 0.6 | 2 | 1.9 | 11.5 | 0.4 | 0.8 | 1 | 1.6 | 3.5 | 16.5 | 19.8 | 20.1 | 16 | 20.2 |
| 2021-11-25 | 45.7 | 54.3 | 2.3 | 2.2 | 2.5 | 0.6 | 2 | 1.9 | 11.5 | 0.4 | 0.8 | 1 | 1.6 | 3.5 | 16.5 | 19.8 | 20.1 | 16 | 20.2 |
| 2021-12-02 | 45.7 | 54.3 | 2.3 | 2.2 | 2.5 | 0.6 | 2 | 1.9 | 11.5 | 0.4 | 0.8 | 1 | 1.6 | 3.5 | 16.5 | 19.8 | 20.1 | 16 | 20.2 |
| 2021-12-09 | 45.7 | 54.3 | 2.3 | 2.2 | 2.5 | 0.6 | 2 | 1.9 | 11.5 | 0.4 | 0.8 | 1 | 1.6 | 3.5 | 16.5 | 19.8 | 20 | 16 | 20.2 |
| 2021-12-30 | 45.7 | 54.3 | 2.3 | 2.2 | 2.5 | 0.6 | 2 | 1.9 | 11.5 | 0.4 | 0.8 | 1 | 1.6 | 3.5 | 16.5 | 19.9 | 20 | 16 | 20.2 |
| 2022-01-06 | 45.7 | 54.3 | 2.3 | 2.1 | 2.5 | 0.7 | 2 | 1.9 | 11.5 | 0.4 | 0.8 | 1 | 1.6 | 3.6 | 16.7 | 19.8 | 20 | 16 | 20.1 |
| 2022-01-13 | 45.6 | 54.4 | 2.3 | 2.1 | 2.5 | 0.7 | 2 | 1.9 | 11.5 | 0.4 | 0.8 | 1 | 1.6 | 3.6 | 16.7 | 19.9 | 20 | 15.9 | 20.1 |
| 2022-01-27 | 45.5 | 54.5 | 2.3 | 2.1 | 2.5 | 0.7 | 2 | 1.9 | 11.5 | 0.4 | 0.9 | 1.1 | 1.7 | 3.6 | 16.9 | 19.9 | 19.9 | 15.7 | 19.9 |

### 3 Database table of COVID-19 exclusive beds

| | ICU TOTAL | | INF. TOTAL | | HMRS - ICU | | HMRS - INF | | Unimed - ICU | | Unimed - INF | | São Lucas - ICU | | São Lucas - INF | | Serrano - ICU | | Serrano - INF | |
|---|---|---|---|---|---|---|---|---|---|---|---|---|---|---|---|---|---|---|---|---|
| Date | Occ. | Bed | Occ. | Bed | Occ. | Bed | Occ. | Bed | Occ. | Bed | Occ. | Bed | Occ. | Bed | Occ. | Bed | Occ. | Bed | Occ. | Bed |
| 2020-06-24 | 14 | 31 | 21 | 75 | 10 | 10 | 9 | 17 | 1 | 10 | 5 | 28 | 1 | 6 | 5 | 20 | 2 | 5 | 2 | 10 |
| 2020-06-25 | 15 | 31 | 20 | 75 | 10 | 10 | 9 | 17 | 1 | 10 | 5 | 28 | 2 | 6 | 5 | 20 | 2 | 5 | 1 | 10 |
| 2020-06-26 | 16 | 31 | 21 | 75 | 10 | 10 | 13 | 17 | 2 | 10 | 4 | 28 | 2 | 6 | 3 | 20 | 2 | 5 | 1 | 10 |
| 2020-06-29 | 18 | 31 | 19 | 75 | 10 | 10 | 7 | 17 | 3 | 10 | 6 | 28 | 3 | 6 | 3 | 20 | 2 | 5 | 3 | 10 |
| 2020-06-30 | 17 | 31 | 22 | 75 | 9 | 10 | 8 | 17 | 3 | 10 | 7 | 28 | 3 | 6 | 4 | 20 | 2 | 5 | 3 | 10 |
| 2020-07-01 | 18 | 31 | 16 | 75 | 7 | 10 | 8 | 17 | 5 | 10 | 3 | 28 | 4 | 6 | 2 | 20 | 2 | 5 | 3 | 10 |
| 2020-07-06 | 17 | 31 | 29 | 75 | 10 | 10 | 16 | 17 | 3 | 10 | 8 | 28 | 3 | 6 | 4 | 20 | 1 | 5 | 1 | 10 |
| 2020-07-08 | 16 | 31 | 31 | 75 | 9 | 10 | 9 | 17 | 3 | 10 | 11 | 28 | 3 | 6 | 8 | 20 | 1 | 5 | 3 | 10 |
| 2020-07-09 | 19 | 31 | 28 | 75 | 10 | 10 | 7 | 17 | 5 | 10 | 10 | 28 | 3 | 6 | 8 | 20 | 1 | 5 | 3 | 10 |
| 2020-07-10 | 21 | 31 | 27 | 75 | 10 | 10 | 9 | 17 | 6 | 10 | 7 | 28 | 4 | 6 | 6 | 20 | 1 | 5 | 5 | 10 |
| 2020-07-13 | 23 | 28 | 37 | 73 | 10 | 10 | 15 | 17 | 6 | 10 | 10 | 28 | 5 | 6 | 5 | 20 | 2 | 2 | 7 | 8 |
| 2020-07-14 | 22 | 28 | 35 | 64 | 9 | 10 | 13 | 17 | 7 | 10 | 7 | 21 | 4 | 6 | 8 | 18 | 2 | 2 | 7 | 8 |
| 2020-07-15 | 19 | 28 | 34 | 63 | 7 | 10 | 14 | 17 | 6 | 10 | 7 | 21 | 4 | 6 | 6 | 18 | 2 | 2 | 7 | 7 |
| 2020-07-16 | 20 | 28 | 32 | 63 | 9 | 10 | 8 | 17 | 5 | 10 | 10 | 21 | 4 | 6 | 7 | 18 | 2 | 2 | 7 | 7 |
| 2020-07-17 | 21 | 28 | 31 | 63 | 10 | 10 | 12 | 17 | 5 | 10 | 7 | 21 | 4 | 6 | 5 | 18 | 2 | 2 | 7 | 7 |
| 2020-07-18 | 19 | 28 | 26 | 65 | 9 | 10 | 12 | 17 | 4 | 10 | 5 | 21 | 4 | 6 | 5 | 20 | 2 | 2 | 4 | 7 |
| 2020-07-19 | 20 | 28 | 24 | 65 | 9 | 10 | 10 | 17 | 4 | 10 | 5 | 21 | 5 | 6 | 2 | 20 | 2 | 2 | 7 | 7 |
| 2020-07-20 | 23 | 28 | 21 | 63 | 10 | 10 | 12 | 17 | 6 | 10 | 5 | 21 | 5 | 6 | 2 | 18 | 2 | 2 | 2 | 7 |
| 2020-07-21 | 21 | 28 | 18 | 63 | 9 | 10 | 12 | 17 | 6 | 10 | 2 | 21 | 5 | 6 | 2 | 18 | 1 | 2 | 2 | 7 |
| 2020-07-22 | 15 | 28 | 30 | 63 | 7 | 10 | 17 | 17 | 3 | 10 | 5 | 21 | 4 | 6 | 5 | 18 | 1 | 2 | 3 | 7 |
| 2020-07-23 | 20 | 28 | 29 | 63 | 8 | 10 | 14 | 17 | 6 | 10 | 7 | 21 | 5 | 6 | 5 | 18 | 1 | 2 | 3 | 7 |
| 2020-07-24 | 20 | 28 | 25 | 65 | 8 | 10 | 8 | 19 | 6 | 10 | 8 | 21 | 5 | 6 | 6 | 18 | 1 | 2 | 3 | 7 |
| 2020-07-25 | 19 | 28 | 23 | 65 | 8 | 10 | 5 | 19 | 4 | 10 | 8 | 21 | 6 | 6 | 7 | 18 | 1 | 2 | 3 | 7 |
| 2020-07-26 | 21 | 28 | 27 | 65 | 7 | 10 | 9 | 19 | 7 | 10 | 9 | 21 | 6 | 6 | 6 | 18 | 1 | 2 | 3 | 7 |
| 2020-07-27 | 21 | 28 | 22 | 65 | 7 | 10 | 10 | 19 | 7 | 10 | 7 | 21 | 6 | 6 | 4 | 18 | 1 | 2 | 1 | 7 |
| 2020-07-28 | 21 | 28 | 35 | 65 | 7 | 10 | 18 | 19 | 8 | 10 | 8 | 21 | 5 | 6 | 7 | 18 | 1 | 2 | 2 | 7 |
| 2020-07-29 | 14 | 28 | 36 | 65 | 7 | 10 | 16 | 19 | 2 | 10 | 9 | 21 | 5 | 6 | 9 | 18 | 0 | 2 | 2 | 7 |

| | | | | | | | | | | | | | | | | | | | | |
|---|---|---|---|---|---|---|---|---|---|---|---|---|---|---|---|---|---|---|---|---|
| 2020-07-30 | 18 | 28 | 37 | 65 | 9 | 10 | 15 | 19 | 2 | 10 | 11 | 21 | 6 | 6 | 7 | 18 | 1 | 2 | 4 | 7 |
| 2020-07-31 | 18 | 28 | 35 | 65 | 9 | 10 | 12 | 19 | 2 | 10 | 12 | 21 | 6 | 6 | 7 | 18 | 1 | 2 | 4 | 7 |
| 2020-08-01 | 14 | 38 | 42 | 65 | 7 | 20 | 16 | 19 | 1 | 10 | 13 | 21 | 5 | 6 | 10 | 18 | 1 | 2 | 3 | 7 |
| 2020-08-02 | 18 | 38 | 42 | 65 | 9 | 20 | 14 | 19 | 2 | 10 | 16 | 21 | 6 | 6 | 9 | 18 | 1 | 2 | 3 | 7 |
| 2020-08-03 | 21 | 38 | 41 | 65 | 10 | 20 | 11 | 19 | 4 | 10 | 18 | 21 | 6 | 6 | 8 | 18 | 1 | 2 | 4 | 7 |
| 2020-08-04 | 21 | 38 | 44 | 65 | 10 | 20 | 12 | 19 | 5 | 10 | 18 | 21 | 5 | 6 | 10 | 18 | 1 | 2 | 4 | 7 |
| 2020-08-05 | 20 | 38 | 43 | 68 | 10 | 20 | 16 | 19 | 3 | 10 | 15 | 21 | 6 | 6 | 7 | 18 | 1 | 2 | 5 | 10 |
| 2020-08-06 | 21 | 38 | 45 | 68 | 10 | 20 | 17 | 19 | 5 | 10 | 15 | 21 | 5 | 6 | 8 | 18 | 1 | 2 | 5 | 10 |
| 2020-08-07 | 20 | 38 | 41 | 68 | 9 | 20 | 14 | 19 | 5 | 10 | 13 | 21 | 5 | 6 | 9 | 18 | 1 | 2 | 5 | 10 |
| 2020-08-08 | 16 | 38 | 40 | 68 | 7 | 20 | 12 | 19 | 3 | 10 | 14 | 21 | 5 | 6 | 7 | 18 | 1 | 2 | 7 | 10 |
| 2020-08-09 | 14 | 38 | 41 | 68 | 7 | 20 | 12 | 19 | 2 | 10 | 14 | 21 | 4 | 6 | 8 | 18 | 1 | 2 | 7 | 10 |
| 2020-08-10 | 14 | 38 | 38 | 68 | 6 | 20 | 11 | 19 | 2 | 10 | 13 | 21 | 5 | 6 | 7 | 18 | 1 | 2 | 7 | 10 |
| 2020-08-11 | 15 | 38 | 29 | 68 | 6 | 20 | 7 | 19 | 2 | 10 | 9 | 21 | 6 | 6 | 5 | 18 | 1 | 2 | 8 | 10 |
| 2020-08-12 | 19 | 38 | 28 | 68 | 6 | 20 | 8 | 19 | 6 | 10 | 7 | 21 | 6 | 6 | 5 | 18 | 1 | 2 | 8 | 10 |
| 2020-08-13 | 22 | 38 | 25 | 68 | 7 | 20 | 7 | 19 | 8 | 10 | 8 | 21 | 6 | 6 | 6 | 18 | 1 | 2 | 5 | 10 |
| 2020-08-14 | 21 | 38 | 24 | 68 | 7 | 20 | 9 | 19 | 8 | 10 | 5 | 21 | 5 | 6 | 5 | 18 | 1 | 2 | 5 | 10 |
| 2020-08-15 | 22 | 38 | 28 | 68 | 6 | 20 | 9 | 19 | 8 | 10 | 6 | 21 | 6 | 6 | 5 | 18 | 2 | 2 | 8 | 10 |
| 2020-08-16 | 25 | 38 | 29 | 68 | 7 | 20 | 11 | 19 | 10 | 10 | 6 | 21 | 6 | 6 | 4 | 18 | 2 | 2 | 8 | 10 |
| 2020-08-17 | 23 | 38 | 28 | 68 | 8 | 20 | 11 | 19 | 8 | 10 | 8 | 21 | 5 | 6 | 2 | 18 | 2 | 2 | 7 | 10 |
| 2020-08-18 | 25 | 38 | 31 | 68 | 9 | 20 | 14 | 19 | 8 | 10 | 7 | 21 | 6 | 6 | 3 | 18 | 2 | 2 | 7 | 10 |
| 2020-08-20 | 22 | 40 | 33 | 68 | 6 | 20 | 16 | 19 | 7 | 10 | 9 | 21 | 7 | 8 | 2 | 18 | 2 | 2 | 6 | 10 |
| 2020-08-21 | 24 | 40 | 36 | 68 | 9 | 20 | 14 | 19 | 7 | 10 | 10 | 21 | 6 | 8 | 5 | 18 | 2 | 2 | 7 | 10 |
| 2020-08-22 | 21 | 40 | 38 | 68 | 9 | 20 | 15 | 19 | 5 | 10 | 10 | 21 | 5 | 8 | 6 | 18 | 2 | 2 | 7 | 10 |
| 2020-08-23 | 18 | 40 | 38 | 68 | 7 | 20 | 17 | 19 | 5 | 10 | 8 | 21 | 4 | 8 | 6 | 18 | 2 | 2 | 7 | 10 |
| 2020-08-24 | 19 | 40 | 35 | 68 | 8 | 20 | 18 | 19 | 5 | 10 | 6 | 21 | 4 | 8 | 6 | 18 | 2 | 2 | 5 | 10 |
| 2020-08-25 | 18 | 40 | 32 | 68 | 8 | 20 | 14 | 19 | 5 | 10 | 5 | 21 | 4 | 8 | 8 | 18 | 1 | 2 | 5 | 10 |
| 2020-08-26 | 18 | 42 | 33 | 68 | 9 | 20 | 17 | 19 | 5 | 10 | 6 | 21 | 3 | 10 | 6 | 18 | 1 | 2 | 4 | 10 |
| 2020-08-27 | 21 | 42 | 38 | 68 | 10 | 20 | 14 | 19 | 3 | 10 | 10 | 21 | 7 | 10 | 5 | 18 | 1 | 2 | 4 | 10 |
| 2020-08-29 | 21 | 42 | 34 | 68 | 11 | 20 | 13 | 19 | 6 | 10 | 12 | 21 | 3 | 10 | 6 | 18 | 1 | 2 | 3 | 10 |
| 2020-08-30 | 22 | 42 | 35 | 68 | 14 | 20 | 13 | 19 | 4 | 10 | 13 | 21 | 3 | 10 | 6 | 18 | 1 | 2 | 3 | 10 |
| 2020-08-31 | 22 | 42 | 35 | 68 | 13 | 20 | 17 | 19 | 5 | 10 | 13 | 21 | 3 | 10 | 3 | 18 | 1 | 2 | 5 | 10 |
| 2020-09-01 | 23 | 42 | 30 | 68 | 14 | 20 | 14 | 19 | 5 | 10 | 10 | 21 | 3 | 10 | 4 | 18 | 1 | 2 | 3 | 10 |
| 2020-09-04 | 23 | 42 | 22 | 70 | 13 | 20 | 10 | 19 | 6 | 10 | 7 | 23 | 3 | 10 | 2 | 18 | 1 | 2 | 3 | 10 |
| 2020-09-10 | 21 | 42 | 23 | 70 | 13 | 20 | 14 | 19 | 6 | 10 | 4 | 23 | 0 | 10 | 1 | 18 | 2 | 2 | 4 | 10 |
| 2020-09-11 | 20 | 42 | 23 | 70 | 11 | 20 | 13 | 19 | 6 | 10 | 6 | 23 | 1 | 10 | 1 | 18 | 2 | 2 | 3 | 10 |
| 2020-09-12 | 21 | 42 | 22 | 70 | 11 | 20 | 12 | 19 | 6 | 10 | 7 | 23 | 2 | 10 | 0 | 18 | 2 | 2 | 3 | 10 |
| 2020-09-13 | 21 | 42 | 23 | 70 | 12 | 20 | 12 | 19 | 5 | 10 | 8 | 23 | 2 | 10 | 0 | 18 | 2 | 2 | 3 | 10 |
| 2020-09-14 | 18 | 42 | 23 | 70 | 10 | 20 | 11 | 19 | 5 | 10 | 8 | 23 | 1 | 10 | 1 | 18 | 2 | 2 | 3 | 10 |
| 2020-09-15 | 18 | 42 | 16 | 70 | 10 | 20 | 12 | 19 | 5 | 10 | 2 | 23 | 1 | 10 | 1 | 18 | 2 | 2 | 1 | 10 |
| 2020-09-16 | 19 | 42 | 19 | 70 | 12 | 20 | 11 | 19 | 4 | 10 | 5 | 23 | 1 | 10 | 2 | 18 | 2 | 2 | 1 | 10 |
| 2020-09-17 | 20 | 42 | 18 | 70 | 13 | 20 | 6 | 19 | 4 | 10 | 7 | 23 | 1 | 10 | 2 | 18 | 2 | 2 | 1 | 10 |
| 2020-09-18 | 20 | 42 | 19 | 70 | 12 | 20 | 8 | 19 | 5 | 10 | 6 | 23 | 1 | 10 | 4 | 18 | 2 | 2 | 1 | 10 |

| | | | | | | | | | | | | | | | | | | | | |
|---|---|---|---|---|---|---|---|---|---|---|---|---|---|---|---|---|---|---|---|---|
| 2020-09-19 | 19 | 42 | 21 | 70 | 11 | 20 | 11 | 19 | 5 | 10 | 6 | 23 | 1 | 10 | 4 | 18 | 2 | 2 | 0 | 10 |
| 2020-09-20 | 21 | 42 | 21 | 70 | 12 | 20 | 13 | 19 | 6 | 10 | 5 | 23 | 1 | 10 | 3 | 18 | 2 | 2 | 0 | 10 |
| 2020-09-21 | 20 | 42 | 14 | 70 | 11 | 20 | 8 | 19 | 6 | 10 | 4 | 23 | 1 | 10 | 2 | 18 | 2 | 2 | 0 | 10 |
| 2020-09-22 | 20 | 42 | 25 | 70 | 13 | 20 | 14 | 19 | 4 | 10 | 7 | 23 | 1 | 10 | 3 | 18 | 2 | 2 | 1 | 10 |
| 2020-09-23 | 19 | 42 | 24 | 70 | 13 | 20 | 13 | 19 | 3 | 10 | 9 | 23 | 1 | 10 | 1 | 18 | 2 | 2 | 1 | 10 |
| 2020-09-24 | 17 | 42 | 26 | 70 | 12 | 20 | 13 | 19 | 3 | 10 | 9 | 23 | 0 | 10 | 1 | 18 | 2 | 2 | 3 | 10 |
| 2020-09-25 | 18 | 42 | 27 | 70 | 12 | 20 | 13 | 19 | 4 | 10 | 8 | 23 | 0 | 10 | 3 | 18 | 2 | 2 | 3 | 10 |
| 2020-09-26 | 17 | 42 | 26 | 70 | 11 | 20 | 13 | 19 | 4 | 10 | 8 | 23 | 1 | 10 | 3 | 18 | 1 | 2 | 2 | 10 |
| 2020-09-27 | 21 | 42 | 25 | 57 | 14 | 20 | 13 | 19 | 4 | 10 | 8 | 10 | 1 | 10 | 2 | 18 | 2 | 2 | 2 | 10 |
| 2020-09-28 | 19 | 42 | 22 | 57 | 14 | 20 | 11 | 19 | 3 | 10 | 9 | 10 | 0 | 10 | 1 | 18 | 2 | 2 | 1 | 10 |
| 2020-09-29 | 17 | 42 | 18 | 57 | 13 | 20 | 10 | 19 | 3 | 10 | 5 | 10 | 0 | 10 | 1 | 18 | 1 | 2 | 2 | 10 |
| 2020-09-30 | 16 | 42 | 21 | 57 | 12 | 20 | 11 | 19 | 2 | 10 | 6 | 10 | 1 | 10 | 2 | 18 | 1 | 2 | 2 | 10 |
| 2020-10-02 | 16 | 42 | 14 | 57 | 11 | 20 | 6 | 19 | 2 | 10 | 6 | 10 | 2 | 10 | 2 | 18 | 1 | 2 | 0 | 10 |
| 2020-10-06 | 18 | 42 | 13 | 57 | 12 | 20 | 4 | 19 | 5 | 10 | 7 | 10 | 1 | 10 | 1 | 18 | 0 | 2 | 1 | 10 |
| 2020-10-07 | 16 | 42 | 7 | 57 | 11 | 20 | 3 | 19 | 3 | 10 | 2 | 10 | 2 | 10 | 1 | 18 | 0 | 2 | 1 | 10 |
| 2020-10-09 | 15 | 42 | 10 | 57 | 9 | 20 | 1 | 19 | 5 | 10 | 5 | 10 | 1 | 10 | 2 | 18 | 0 | 2 | 2 | 10 |
| 2020-10-10 | 18 | 42 | 10 | 57 | 10 | 20 | 2 | 19 | 5 | 10 | 3 | 10 | 2 | 10 | 3 | 18 | 1 | 2 | 2 | 10 |
| 2020-10-11 | 15 | 42 | 15 | 57 | 8 | 20 | 2 | 19 | 4 | 10 | 8 | 10 | 2 | 10 | 3 | 18 | 1 | 2 | 2 | 10 |
| 2020-10-12 | 17 | 42 | 16 | 57 | 8 | 20 | 2 | 19 | 5 | 10 | 8 | 10 | 3 | 10 | 4 | 18 | 1 | 2 | 2 | 10 |
| 2020-10-13 | 15 | 42 | 17 | 57 | 7 | 20 | 3 | 19 | 4 | 10 | 10 | 10 | 3 | 10 | 2 | 18 | 1 | 2 | 2 | 10 |
| 2020-10-14 | 12 | 42 | 13 | 57 | 5 | 20 | 0 | 19 | 4 | 10 | 9 | 10 | 1 | 10 | 2 | 18 | 2 | 2 | 2 | 10 |
| 2020-10-15 | 14 | 42 | 15 | 57 | 6 | 20 | 1 | 19 | 6 | 10 | 9 | 10 | 0 | 10 | 3 | 18 | 2 | 2 | 2 | 10 |
| 2020-10-16 | 13 | 42 | 17 | 57 | 3 | 20 | 4 | 19 | 8 | 10 | 9 | 10 | 0 | 10 | 2 | 18 | 2 | 2 | 2 | 10 |
| 2020-10-17 | 16 | 42 | 20 | 57 | 4 | 20 | 4 | 19 | 9 | 10 | 10 | 10 | 1 | 10 | 3 | 18 | 2 | 2 | 3 | 10 |
| 2020-10-18 | 15 | 42 | 21 | 57 | 4 | 20 | 6 | 19 | 8 | 10 | 10 | 10 | 1 | 10 | 3 | 18 | 2 | 2 | 2 | 10 |
| 2020-10-19 | 13 | 42 | 18 | 57 | 5 | 20 | 5 | 19 | 6 | 10 | 10 | 10 | 0 | 10 | 2 | 18 | 2 | 2 | 1 | 10 |
| 2020-10-20 | 15 | 42 | 16 | 57 | 5 | 20 | 6 | 19 | 7 | 10 | 8 | 10 | 1 | 10 | 1 | 18 | 2 | 2 | 1 | 10 |
| 2020-10-21 | 14 | 42 | 18 | 57 | 5 | 20 | 6 | 19 | 7 | 10 | 8 | 10 | 1 | 10 | 3 | 18 | 1 | 2 | 1 | 10 |
| 2020-10-22 | 11 | 42 | 18 | 57 | 4 | 20 | 7 | 19 | 5 | 10 | 7 | 10 | 1 | 10 | 3 | 18 | 1 | 2 | 1 | 10 |
| 2020-10-23 | 11 | 42 | 18 | 57 | 4 | 20 | 7 | 19 | 5 | 10 | 7 | 10 | 2 | 10 | 3 | 18 | 0 | 2 | 1 | 10 |
| 2020-10-24 | 12 | 42 | 16 | 57 | 5 | 20 | 6 | 19 | 5 | 10 | 6 | 10 | 2 | 10 | 3 | 18 | 0 | 2 | 1 | 10 |
| 2020-10-25 | 12 | 42 | 17 | 57 | 5 | 20 | 6 | 19 | 5 | 10 | 8 | 10 | 2 | 10 | 3 | 18 | 0 | 2 | 0 | 10 |
| 2020-10-26 | 14 | 42 | 18 | 57 | 5 | 20 | 6 | 19 | 5 | 10 | 9 | 10 | 4 | 10 | 3 | 18 | 0 | 2 | 0 | 10 |
| 2020-10-27 | 14 | 42 | 15 | 57 | 6 | 20 | 3 | 19 | 5 | 10 | 7 | 10 | 3 | 10 | 4 | 18 | 0 | 2 | 1 | 10 |
| 2020-10-28 | 11 | 42 | 14 | 57 | 5 | 20 | 2 | 19 | 3 | 10 | 7 | 10 | 3 | 10 | 3 | 18 | 0 | 2 | 2 | 10 |
| 2020-10-29 | 12 | 42 | 14 | 57 | 4 | 20 | 3 | 19 | 5 | 10 | 6 | 10 | 3 | 10 | 4 | 18 | 0 | 2 | 1 | 10 |
| 2020-10-30 | 11 | 42 | 19 | 57 | 3 | 20 | 3 | 19 | 5 | 10 | 8 | 10 | 3 | 10 | 5 | 18 | 0 | 2 | 3 | 10 |
| 2020-10-31 | 13 | 42 | 18 | 57 | 4 | 20 | 4 | 19 | 5 | 10 | 8 | 10 | 4 | 10 | 2 | 18 | 0 | 2 | 4 | 10 |
| 2020-11-01 | 17 | 42 | 17 | 57 | 5 | 20 | 5 | 19 | 8 | 10 | 5 | 10 | 4 | 10 | 3 | 18 | 0 | 2 | 4 | 10 |
| 2020-11-02 | 14 | 42 | 18 | 57 | 5 | 20 | 6 | 19 | 5 | 10 | 4 | 10 | 4 | 10 | 3 | 18 | 0 | 2 | 5 | 10 |
| 2020-11-03 | 15 | 42 | 19 | 57 | 4 | 20 | 6 | 19 | 6 | 10 | 4 | 10 | 4 | 10 | 5 | 18 | 1 | 2 | 4 | 10 |
| 2020-11-04 | 11 | 42 | 17 | 57 | 3 | 20 | 5 | 19 | 4 | 10 | 6 | 10 | 3 | 10 | 3 | 18 | 1 | 2 | 3 | 10 |

| | | | | | | | | | | | | | | | | | | | | |
|---|---|---|---|---|---|---|---|---|---|---|---|---|---|---|---|---|---|---|---|---|
| 2020-11-05 | 11 | 42 | 21 | 57 | 3 | 20 | 6 | 19 | 2 | 10 | 7 | 10 | 4 | 10 | 4 | 18 | 2 | 2 | 4 | 10 |
| 2020-11-06 | 13 | 42 | 11 | 57 | 4 | 20 | 4 | 19 | 2 | 10 | 5 | 10 | 5 | 10 | 1 | 18 | 2 | 2 | 1 | 10 |
| 2020-11-07 | 11 | 42 | 13 | 57 | 3 | 20 | 5 | 19 | 2 | 10 | 7 | 10 | 5 | 10 | 1 | 18 | 1 | 2 | 0 | 10 |
| 2020-11-08 | 13 | 42 | 16 | 57 | 4 | 20 | 5 | 19 | 4 | 10 | 8 | 10 | 4 | 10 | 3 | 18 | 1 | 2 | 0 | 10 |
| 2020-11-09 | 10 | 42 | 13 | 57 | 4 | 20 | 5 | 19 | 2 | 10 | 4 | 10 | 3 | 10 | 4 | 18 | 1 | 2 | 0 | 10 |
| 2020-11-10 | 10 | 42 | 19 | 57 | 4 | 20 | 5 | 19 | 2 | 10 | 4 | 10 | 3 | 10 | 6 | 18 | 1 | 2 | 4 | 10 |
| 2020-11-11 | 10 | 42 | 21 | 57 | 4 | 20 | 4 | 19 | 3 | 10 | 6 | 10 | 2 | 10 | 7 | 18 | 1 | 2 | 4 | 10 |
| 2020-11-12 | 11 | 42 | 26 | 60 | 4 | 20 | 3 | 19 | 3 | 10 | 13 | 13 | 2 | 10 | 7 | 18 | 2 | 2 | 3 | 10 |
| 2020-11-13 | 11 | 42 | 23 | 60 | 4 | 20 | 3 | 19 | 2 | 10 | 13 | 13 | 3 | 10 | 4 | 18 | 2 | 2 | 3 | 10 |
| 2020-11-14 | 13 | 42 | 19 | 60 | 4 | 20 | 2 | 19 | 3 | 10 | 13 | 13 | 4 | 10 | 2 | 18 | 2 | 2 | 2 | 10 |
| 2020-11-15 | 18 | 42 | 18 | 60 | 8 | 20 | 2 | 19 | 3 | 10 | 13 | 13 | 5 | 10 | 2 | 18 | 2 | 2 | 1 | 10 |
| 2020-11-16 | 18 | 42 | 17 | 59 | 9 | 20 | 1 | 19 | 3 | 10 | 12 | 12 | 5 | 10 | 2 | 18 | 1 | 2 | 2 | 10 |
| 2020-11-17 | 16 | 42 | 17 | 57 | 7 | 20 | 5 | 19 | 3 | 10 | 9 | 10 | 5 | 10 | 2 | 18 | 1 | 2 | 1 | 10 |
| 2020-11-18 | 17 | 42 | 19 | 57 | 7 | 20 | 7 | 19 | 4 | 10 | 8 | 10 | 4 | 10 | 1 | 18 | 2 | 2 | 3 | 10 |
| 2020-12-31 | 28 | 42 | 51 | 74 | 15 | 20 | 15 | 19 | 7 | 10 | 18 | 25 | 4 | 10 | 12 | 20 | 2 | 2 | 6 | 10 |
| 2021-01-01 | 28 | 42 | 47 | 74 | 15 | 20 | 14 | 19 | 6 | 10 | 16 | 25 | 5 | 10 | 11 | 20 | 2 | 2 | 6 | 10 |
| 2021-01-02 | 29 | 42 | 43 | 74 | 14 | 20 | 15 | 19 | 8 | 10 | 14 | 25 | 5 | 10 | 11 | 20 | 2 | 2 | 3 | 10 |
| 2021-01-03 | 29 | 42 | 48 | 74 | 16 | 20 | 16 | 19 | 6 | 10 | 19 | 25 | 5 | 10 | 10 | 20 | 2 | 2 | 3 | 10 |
| 2021-01-04 | 25 | 42 | 45 | 74 | 15 | 20 | 16 | 19 | 7 | 10 | 18 | 25 | 2 | 10 | 9 | 20 | 1 | 2 | 2 | 10 |
| 2021-01-05 | 29 | 42 | 44 | 74 | 16 | 20 | 12 | 19 | 8 | 10 | 16 | 25 | 3 | 10 | 11 | 20 | 2 | 2 | 5 | 10 |
| 2021-01-06 | 30 | 42 | 36 | 74 | 17 | 20 | 5 | 19 | 8 | 10 | 17 | 25 | 3 | 10 | 10 | 20 | 2 | 2 | 4 | 10 |
| 2021-01-07 | 33 | 42 | 42 | 74 | 18 | 20 | 8 | 19 | 10 | 10 | 20 | 25 | 3 | 10 | 10 | 20 | 2 | 2 | 4 | 10 |
| 2021-01-08 | 34 | 42 | 46 | 74 | 18 | 20 | 12 | 19 | 9 | 10 | 21 | 25 | 5 | 10 | 9 | 20 | 2 | 2 | 4 | 10 |
| 2021-01-11 | 33 | 42 | 52 | 74 | 16 | 20 | 17 | 19 | 10 | 10 | 19 | 25 | 5 | 10 | 12 | 20 | 2 | 2 | 4 | 10 |
| 2021-01-12 | 31 | 42 | 51 | 74 | 13 | 20 | 18 | 19 | 10 | 10 | 15 | 25 | 7 | 10 | 13 | 20 | 1 | 2 | 5 | 10 |
| 2021-01-13 | 27 | 42 | 50 | 74 | 12 | 20 | 16 | 19 | 7 | 10 | 19 | 25 | 7 | 10 | 11 | 20 | 1 | 2 | 4 | 10 |
| 2021-01-14 | 27 | 42 | 55 | 74 | 14 | 20 | 19 | 19 | 5 | 10 | 20 | 25 | 7 | 10 | 11 | 20 | 1 | 2 | 5 | 10 |
| 2021-01-15 | 30 | 42 | 55 | 74 | 16 | 20 | 17 | 19 | 7 | 10 | 22 | 25 | 6 | 10 | 12 | 20 | 1 | 2 | 4 | 10 |
| 2021-01-16 | 32 | 42 | 61 | 74 | 16 | 20 | 19 | 19 | 9 | 10 | 25 | 25 | 5 | 10 | 15 | 20 | 2 | 2 | 2 | 10 |
| 2021-01-17 | 38 | 42 | 54 | 74 | 20 | 20 | 19 | 19 | 10 | 10 | 21 | 25 | 6 | 10 | 12 | 20 | 2 | 2 | 2 | 10 |
| 2021-01-18 | 39 | 42 | 52 | 74 | 20 | 20 | 19 | 19 | 10 | 10 | 21 | 25 | 8 | 10 | 10 | 20 | 1 | 2 | 2 | 10 |
| 2021-01-19 | 39 | 42 | 58 | 74 | 20 | 20 | 19 | 19 | 10 | 10 | 21 | 25 | 8 | 10 | 12 | 20 | 1 | 2 | 6 | 10 |
| 2021-01-20 | 38 | 42 | 56 | 74 | 20 | 20 | 16 | 19 | 9 | 10 | 24 | 25 | 8 | 10 | 12 | 20 | 1 | 2 | 4 | 10 |
| 2021-01-21 | 37 | 42 | 49 | 74 | 19 | 20 | 19 | 19 | 10 | 10 | 16 | 25 | 7 | 10 | 10 | 20 | 1 | 2 | 4 | 10 |
| 2021-01-22 | 35 | 42 | 44 | 74 | 19 | 20 | 16 | 19 | 8 | 10 | 16 | 25 | 7 | 10 | 9 | 20 | 1 | 2 | 3 | 10 |
| 2021-01-23 | 37 | 42 | 44 | 74 | 20 | 20 | 18 | 19 | 10 | 10 | 16 | 25 | 6 | 10 | 8 | 20 | 1 | 2 | 2 | 10 |
| 2021-01-24 | 35 | 42 | 43 | 74 | 19 | 20 | 19 | 19 | 9 | 10 | 13 | 25 | 6 | 10 | 9 | 20 | 1 | 2 | 2 | 10 |
| 2021-01-25 | 30 | 42 | 39 | 74 | 18 | 20 | 19 | 19 | 5 | 10 | 12 | 25 | 6 | 10 | 8 | 20 | 1 | 2 | 0 | 10 |
| 2021-01-26 | 28 | 42 | 35 | 74 | 16 | 20 | 17 | 19 | 4 | 10 | 13 | 25 | 6 | 10 | 5 | 20 | 2 | 2 | 0 | 10 |
| 2021-01-27 | 26 | 42 | 35 | 74 | 15 | 20 | 15 | 19 | 4 | 10 | 17 | 25 | 5 | 10 | 3 | 20 | 2 | 2 | 0 | 10 |
| 2021-01-28 | 24 | 42 | 34 | 74 | 13 | 20 | 14 | 19 | 4 | 10 | 16 | 25 | 5 | 10 | 4 | 20 | 2 | 2 | 0 | 10 |
| 2021-01-29 | 22 | 42 | 29 | 74 | 13 | 20 | 11 | 19 | 4 | 10 | 13 | 25 | 4 | 10 | 3 | 20 | 1 | 2 | 2 | 10 |

| | | | | | | | | | | | | | | | | | | | | |
|---|---|---|---|---|---|---|---|---|---|---|---|---|---|---|---|---|---|---|---|---|
| 2021-01-30 | 19 | 42 | 35 | 74 | 11 | 20 | 13 | 19 | 3 | 10 | 17 | 25 | 4 | 10 | 3 | 20 | 1 | 2 | 2 | 10 |
| 2021-01-31 | 19 | 42 | 36 | 74 | 10 | 20 | 14 | 19 | 4 | 10 | 16 | 25 | 4 | 10 | 4 | 20 | 1 | 2 | 2 | 10 |
| 2021-02-01 | 14 | 42 | 31 | 74 | 5 | 20 | 13 | 19 | 5 | 10 | 11 | 25 | 3 | 10 | 5 | 20 | 1 | 2 | 2 | 10 |
| 2021-02-02 | 18 | 42 | 36 | 74 | 6 | 20 | 18 | 19 | 6 | 10 | 13 | 25 | 5 | 10 | 3 | 20 | 1 | 2 | 2 | 10 |
| 2021-02-03 | 20 | 42 | 42 | 77 | 8 | 20 | 21 | 22 | 5 | 10 | 15 | 25 | 6 | 10 | 4 | 20 | 1 | 2 | 2 | 10 |
| 2021-02-04 | 24 | 42 | 30 | 77 | 11 | 20 | 8 | 22 | 7 | 10 | 17 | 25 | 6 | 10 | 4 | 20 | 0 | 2 | 1 | 10 |
| 2021-02-05 | 23 | 42 | 29 | 77 | 10 | 20 | 11 | 22 | 7 | 10 | 15 | 25 | 6 | 10 | 2 | 20 | 0 | 2 | 1 | 10 |
| 2021-02-06 | 25 | 42 | 35 | 77 | 15 | 20 | 16 | 22 | 5 | 10 | 17 | 25 | 5 | 10 | 2 | 20 | 0 | 2 | 0 | 10 |
| 2021-02-07 | 24 | 42 | 31 | 77 | 14 | 20 | 18 | 22 | 5 | 10 | 12 | 25 | 5 | 10 | 1 | 20 | 0 | 2 | 0 | 10 |
| 2021-02-08 | 20 | 42 | 31 | 77 | 9 | 20 | 20 | 22 | 5 | 10 | 10 | 25 | 6 | 10 | 1 | 20 | 0 | 2 | 0 | 10 |
| 2021-02-09 | 21 | 42 | 28 | 77 | 9 | 20 | 16 | 22 | 5 | 10 | 11 | 25 | 6 | 10 | 1 | 20 | 1 | 2 | 0 | 10 |
| 2021-02-10 | 19 | 42 | 30 | 77 | 10 | 20 | 16 | 22 | 1 | 10 | 11 | 25 | 7 | 10 | 2 | 20 | 1 | 2 | 1 | 10 |
| 2021-02-11 | 19 | 42 | 26 | 77 | 10 | 20 | 11 | 22 | 1 | 10 | 12 | 25 | 7 | 10 | 1 | 20 | 1 | 2 | 2 | 10 |
| 2021-02-12 | 19 | 42 | 25 | 77 | 10 | 20 | 13 | 22 | 1 | 10 | 9 | 25 | 7 | 10 | 1 | 20 | 1 | 2 | 2 | 10 |
| 2021-02-13 | 17 | 42 | 30 | 77 | 7 | 20 | 17 | 22 | 2 | 10 | 10 | 25 | 8 | 10 | 1 | 20 | 0 | 2 | 2 | 10 |
| 2021-02-14 | 22 | 42 | 35 | 77 | 10 | 20 | 19 | 22 | 4 | 10 | 12 | 25 | 8 | 10 | 2 | 20 | 0 | 2 | 2 | 10 |
| 2021-02-15 | 24 | 42 | 38 | 77 | 12 | 20 | 20 | 22 | 4 | 10 | 12 | 25 | 7 | 10 | 4 | 20 | 1 | 2 | 2 | 10 |
| 2021-02-16 | 24 | 42 | 33 | 77 | 12 | 20 | 21 | 22 | 6 | 10 | 8 | 25 | 5 | 10 | 2 | 20 | 1 | 2 | 2 | 10 |
| 2021-02-17 | 24 | 42 | 28 | 77 | 13 | 20 | 19 | 22 | 5 | 10 | 6 | 25 | 4 | 10 | 2 | 20 | 2 | 2 | 1 | 10 |
| 2021-02-18 | 22 | 42 | 23 | 77 | 11 | 20 | 13 | 22 | 5 | 10 | 6 | 25 | 4 | 10 | 3 | 20 | 2 | 2 | 1 | 10 |
| 2021-02-19 | 22 | 42 | 21 | 77 | 12 | 20 | 10 | 22 | 5 | 10 | 5 | 25 | 4 | 10 | 3 | 20 | 1 | 2 | 3 | 10 |
| 2021-02-20 | 25 | 42 | 18 | 77 | 12 | 20 | 9 | 22 | 7 | 10 | 4 | 25 | 4 | 10 | 3 | 20 | 2 | 2 | 2 | 10 |
| 2021-02-21 | 25 | 42 | 22 | 77 | 12 | 20 | 10 | 22 | 7 | 10 | 7 | 25 | 4 | 10 | 2 | 20 | 2 | 2 | 3 | 10 |
| 2021-02-22 | 23 | 42 | 19 | 77 | 11 | 20 | 11 | 22 | 7 | 10 | 4 | 25 | 3 | 10 | 3 | 20 | 2 | 2 | 1 | 10 |
| 2021-02-23 | 23 | 42 | 23 | 77 | 10 | 20 | 11 | 22 | 8 | 10 | 5 | 25 | 3 | 10 | 4 | 20 | 2 | 2 | 3 | 10 |
| 2021-02-24 | 22 | 42 | 24 | 77 | 11 | 20 | 8 | 22 | 6 | 10 | 8 | 25 | 3 | 10 | 5 | 20 | 2 | 2 | 3 | 10 |
| 2021-02-25 | 23 | 42 | 21 | 77 | 13 | 20 | 6 | 22 | 5 | 10 | 6 | 25 | 3 | 10 | 5 | 20 | 2 | 2 | 4 | 10 |
| 2021-02-26 | 25 | 42 | 22 | 77 | 13 | 20 | 6 | 22 | 8 | 10 | 7 | 25 | 3 | 10 | 5 | 20 | 1 | 2 | 4 | 10 |
| 2021-02-27 | 25 | 42 | 16 | 77 | 13 | 20 | 4 | 22 | 7 | 10 | 4 | 25 | 4 | 10 | 4 | 20 | 1 | 2 | 4 | 10 |
| 2021-02-28 | 21 | 42 | 20 | 77 | 12 | 20 | 4 | 22 | 4 | 10 | 9 | 25 | 4 | 10 | 4 | 20 | 1 | 2 | 3 | 10 |
| 2021-03-01 | 20 | 42 | 27 | 77 | 11 | 20 | 8 | 22 | 4 | 10 | 9 | 25 | 4 | 10 | 6 | 20 | 1 | 2 | 4 | 10 |
| 2021-03-02 | 20 | 42 | 21 | 77 | 10 | 20 | 6 | 22 | 5 | 10 | 7 | 25 | 4 | 10 | 6 | 20 | 1 | 2 | 2 | 10 |
| 2021-03-03 | 20 | 42 | 19 | 77 | 10 | 20 | 6 | 22 | 5 | 10 | 6 | 25 | 4 | 10 | 4 | 20 | 1 | 2 | 3 | 10 |
| 2021-03-04 | 21 | 42 | 20 | 77 | 9 | 20 | 6 | 22 | 6 | 10 | 9 | 25 | 5 | 10 | 3 | 20 | 1 | 2 | 2 | 10 |
| 2021-03-05 | 17 | 42 | 23 | 77 | 7 | 20 | 9 | 22 | 5 | 10 | 10 | 25 | 4 | 10 | 3 | 20 | 1 | 2 | 1 | 10 |
| 2021-03-06 | 20 | 42 | 25 | 77 | 7 | 20 | 12 | 22 | 7 | 10 | 12 | 25 | 4 | 10 | 1 | 20 | 2 | 2 | 0 | 10 |
| 2021-03-07 | 23 | 42 | 26 | 77 | 9 | 20 | 14 | 22 | 9 | 10 | 9 | 25 | 3 | 10 | 2 | 20 | 2 | 2 | 1 | 10 |
| 2021-03-08 | 19 | 42 | 31 | 77 | 6 | 20 | 19 | 22 | 9 | 10 | 8 | 25 | 3 | 10 | 3 | 20 | 1 | 2 | 1 | 10 |
| 2021-03-09 | 22 | 42 | 24 | 77 | 9 | 20 | 12 | 22 | 7 | 10 | 9 | 25 | 5 | 10 | 2 | 20 | 1 | 2 | 1 | 10 |
| 2021-03-10 | 21 | 42 | 29 | 77 | 6 | 20 | 13 | 22 | 9 | 10 | 12 | 25 | 5 | 10 | 3 | 20 | 1 | 2 | 1 | 10 |
| 2021-03-11 | 23 | 42 | 30 | 77 | 7 | 20 | 12 | 22 | 10 | 10 | 13 | 25 | 5 | 10 | 4 | 20 | 1 | 2 | 1 | 10 |
| 2021-03-12 | 21 | 42 | 28 | 77 | 8 | 20 | 11 | 22 | 9 | 10 | 13 | 25 | 3 | 10 | 3 | 20 | 1 | 2 | 1 | 10 |

| | | | | | | | | | | | | | | | | | | | | |
|---|---|---|---|---|---|---|---|---|---|---|---|---|---|---|---|---|---|---|---|---|
| 2021-03-13 | 23 | 42 | 33 | 77 | 9 | 20 | 14 | 22 | 8 | 10 | 13 | 25 | 4 | 10 | 4 | 20 | 2 | 2 | 2 | 10 |
| 2021-03-14 | 23 | 42 | 42 | 77 | 11 | 20 | 19 | 22 | 7 | 10 | 19 | 25 | 3 | 10 | 3 | 20 | 2 | 2 | 1 | 10 |
| 2021-03-15 | 23 | 42 | 40 | 77 | 12 | 20 | 20 | 22 | 7 | 10 | 16 | 25 | 3 | 10 | 3 | 20 | 1 | 2 | 1 | 10 |
| 2021-03-16 | 28 | 42 | 44 | 77 | 18 | 20 | 20 | 22 | 6 | 10 | 18 | 25 | 2 | 10 | 2 | 20 | 2 | 2 | 4 | 10 |
| 2021-03-17 | 30 | 42 | 52 | 77 | 19 | 20 | 22 | 22 | 7 | 10 | 21 | 25 | 2 | 10 | 3 | 20 | 2 | 2 | 6 | 10 |
| 2021-03-18 | 31 | 42 | 52 | 77 | 19 | 20 | 22 | 22 | 6 | 10 | 18 | 25 | 4 | 10 | 4 | 20 | 2 | 2 | 8 | 10 |
| 2021-03-19 | 30 | 42 | 53 | 77 | 19 | 20 | 22 | 22 | 5 | 10 | 17 | 25 | 4 | 10 | 4 | 20 | 2 | 2 | 10 | 10 |
| 2021-03-20 | 33 | 42 | 58 | 77 | 19 | 20 | 22 | 22 | 8 | 10 | 17 | 25 | 4 | 10 | 9 | 20 | 2 | 2 | 10 | 10 |
| 2021-03-21 | 33 | 42 | 53 | 77 | 20 | 20 | 20 | 22 | 7 | 10 | 14 | 25 | 4 | 10 | 9 | 20 | 2 | 2 | 10 | 10 |
| 2021-03-22 | 31 | 42 | 57 | 77 | 19 | 20 | 22 | 22 | 6 | 10 | 17 | 25 | 4 | 10 | 8 | 20 | 2 | 2 | 10 | 10 |
| 2021-03-23 | 30 | 42 | 63 | 80 | 18 | 20 | 25 | 25 | 7 | 10 | 20 | 25 | 3 | 10 | 8 | 20 | 2 | 2 | 10 | 10 |
| 2021-03-24 | 33 | 42 | 68 | 80 | 20 | 20 | 25 | 25 | 8 | 10 | 22 | 25 | 3 | 10 | 11 | 20 | 2 | 2 | 10 | 10 |
| 2021-03-25 | 31 | 42 | 64 | 80 | 17 | 20 | 25 | 25 | 9 | 10 | 21 | 25 | 3 | 10 | 8 | 20 | 2 | 2 | 10 | 10 |
| 2021-03-26 | 35 | 42 | 69 | 88 | 20 | 20 | 24 | 25 | 9 | 10 | 27 | 33 | 4 | 10 | 8 | 20 | 2 | 2 | 10 | 10 |
| 2021-03-27 | 32 | 42 | 75 | 88 | 17 | 20 | 25 | 25 | 10 | 10 | 30 | 33 | 3 | 10 | 10 | 20 | 2 | 2 | 10 | 10 |
| 2021-03-28 | 39 | 46 | 70 | 86 | 20 | 20 | 23 | 25 | 9 | 10 | 32 | 33 | 4 | 10 | 7 | 20 | 6 | 6 | 8 | 8 |
| 2021-03-29 | 37 | 46 | 68 | 86 | 18 | 20 | 25 | 25 | 9 | 10 | 29 | 33 | 4 | 10 | 9 | 20 | 6 | 6 | 5 | 8 |
| 2021-03-30 | 43 | 46 | 71 | 86 | 20 | 20 | 25 | 25 | 10 | 10 | 31 | 33 | 7 | 10 | 10 | 20 | 6 | 6 | 5 | 8 |
| 2021-03-31 | 43 | 46 | 70 | 89 | 19 | 20 | 25 | 25 | 10 | 10 | 28 | 36 | 8 | 10 | 11 | 20 | 6 | 6 | 6 | 8 |
| 2021-04-01 | 43 | 46 | 76 | 89 | 18 | 20 | 25 | 25 | 10 | 10 | 30 | 36 | 9 | 10 | 13 | 20 | 6 | 6 | 8 | 8 |
| 2021-04-02 | 40 | 46 | 66 | 86 | 17 | 20 | 25 | 25 | 10 | 10 | 22 | 33 | 7 | 10 | 11 | 20 | 6 | 6 | 8 | 8 |
| 2021-04-03 | 47 | 50 | 68 | 90 | 20 | 20 | 25 | 25 | 10 | 10 | 25 | 33 | 7 | 10 | 11 | 20 | 10 | 10 | 7 | 12 |
| 2021-04-04 | 49 | 50 | 68 | 90 | 20 | 20 | 25 | 25 | 10 | 10 | 27 | 33 | 9 | 10 | 10 | 20 | 10 | 10 | 6 | 12 |
| 2021-04-05 | 49 | 50 | 66 | 90 | 20 | 20 | 25 | 25 | 10 | 10 | 26 | 33 | 9 | 10 | 9 | 20 | 10 | 10 | 6 | 12 |
| 2021-04-06 | 49 | 50 | 69 | 93 | 20 | 20 | 25 | 25 | 10 | 10 | 26 | 36 | 9 | 10 | 9 | 20 | 10 | 10 | 9 | 12 |
| 2021-04-07 | 44 | 50 | 94 | 110 | 20 | 20 | 42 | 42 | 10 | 10 | 35 | 36 | 7 | 10 | 9 | 20 | 7 | 10 | 8 | 12 |
| 2021-04-08 | 44 | 50 | 86 | 110 | 20 | 20 | 34 | 42 | 10 | 10 | 35 | 36 | 6 | 10 | 12 | 20 | 8 | 10 | 5 | 12 |
| 2021-04-09 | 44 | 50 | 90 | 110 | 19 | 20 | 37 | 42 | 10 | 10 | 34 | 36 | 6 | 10 | 12 | 20 | 9 | 10 | 7 | 12 |
| 2021-04-10 | 46 | 50 | 95 | 110 | 20 | 20 | 42 | 42 | 10 | 10 | 34 | 36 | 6 | 10 | 12 | 20 | 10 | 10 | 7 | 12 |
| 2021-04-11 | 47 | 50 | 93 | 110 | 20 | 20 | 42 | 42 | 10 | 10 | 33 | 36 | 7 | 10 | 10 | 20 | 10 | 10 | 8 | 12 |
| 2021-04-12 | 44 | 50 | 108 | 126 | 20 | 20 | 58 | 58 | 10 | 10 | 35 | 36 | 6 | 10 | 9 | 20 | 8 | 10 | 6 | 12 |
| 2021-04-13 | 44 | 50 | 110 | 126 | 20 | 20 | 58 | 58 | 10 | 10 | 33 | 36 | 7 | 10 | 9 | 20 | 7 | 10 | 10 | 12 |
| 2021-04-14 | 42 | 50 | 101 | 126 | 17 | 20 | 54 | 58 | 10 | 10 | 33 | 36 | 7 | 10 | 9 | 20 | 8 | 10 | 5 | 12 |
| 2021-04-15 | 44 | 50 | 105 | 126 | 19 | 20 | 52 | 58 | 10 | 10 | 33 | 36 | 7 | 10 | 12 | 20 | 8 | 10 | 8 | 12 |
| 2021-04-16 | 45 | 50 | 110 | 133 | 20 | 20 | 61 | 65 | 10 | 10 | 27 | 36 | 7 | 10 | 14 | 20 | 8 | 10 | 8 | 12 |
| 2021-04-17 | 42 | 50 | 103 | 133 | 19 | 20 | 56 | 65 | 10 | 10 | 25 | 36 | 7 | 10 | 15 | 20 | 6 | 10 | 7 | 12 |
| 2021-04-18 | 45 | 50 | 107 | 133 | 20 | 20 | 58 | 65 | 10 | 10 | 28 | 36 | 6 | 10 | 14 | 20 | 9 | 10 | 7 | 12 |
| 2021-04-19 | 45 | 50 | 111 | 133 | 20 | 20 | 59 | 65 | 10 | 10 | 28 | 36 | 7 | 10 | 17 | 20 | 8 | 10 | 7 | 12 |
| 2021-04-20 | 48 | 50 | 109 | 133 | 20 | 20 | 60 | 65 | 10 | 10 | 27 | 36 | 9 | 10 | 15 | 20 | 9 | 10 | 7 | 12 |
| 2021-04-21 | 49 | 50 | 100 | 133 | 20 | 20 | 57 | 65 | 10 | 10 | 24 | 36 | 9 | 10 | 12 | 20 | 10 | 10 | 7 | 12 |
| 2021-04-22 | 49 | 50 | 110 | 133 | 20 | 20 | 62 | 65 | 10 | 10 | 25 | 36 | 10 | 10 | 16 | 20 | 9 | 10 | 7 | 12 |
| 2021-04-23 | 48 | 50 | 109 | 133 | 20 | 20 | 61 | 65 | 10 | 10 | 29 | 36 | 10 | 10 | 15 | 20 | 8 | 10 | 4 | 12 |

| | | | | | | | | | | | | | | | | | | | | |
|---|---|---|---|---|---|---|---|---|---|---|---|---|---|---|---|---|---|---|---|---|
| 2021-04-24 | 45 | 50 | 116 | 133 | 19 | 20 | 63 | 65 | 10 | 10 | 30 | 36 | 10 | 10 | 16 | 20 | 6 | 10 | 7 | 12 |
| 2021-04-25 | 46 | 50 | 117 | 133 | 20 | 20 | 60 | 65 | 10 | 10 | 33 | 36 | 10 | 10 | 17 | 20 | 6 | 10 | 7 | 12 |
| 2021-04-26 | 46 | 50 | 117 | 133 | 20 | 20 | 65 | 65 | 10 | 10 | 31 | 36 | 10 | 10 | 14 | 20 | 6 | 10 | 7 | 12 |
| 2021-04-27 | 43 | 50 | 118 | 133 | 20 | 20 | 61 | 65 | 10 | 10 | 34 | 36 | 7 | 10 | 17 | 20 | 6 | 10 | 6 | 12 |
| 2021-04-28 | 39 | 50 | 115 | 133 | 18 | 20 | 58 | 65 | 10 | 10 | 36 | 36 | 5 | 10 | 16 | 20 | 6 | 10 | 5 | 12 |
| 2021-04-29 | 40 | 50 | 108 | 133 | 20 | 20 | 55 | 65 | 10 | 10 | 36 | 36 | 6 | 10 | 12 | 20 | 4 | 10 | 5 | 12 |
| 2021-04-30 | 42 | 50 | 111 | 140 | 20 | 20 | 52 | 65 | 10 | 10 | 43 | 43 | 8 | 10 | 12 | 20 | 4 | 10 | 4 | 12 |
| 2021-05-01 | 40 | 50 | 109 | 138 | 20 | 20 | 52 | 65 | 10 | 10 | 41 | 41 | 8 | 10 | 11 | 20 | 2 | 10 | 5 | 12 |
| 2021-05-02 | 40 | 50 | 115 | 137 | 20 | 20 | 60 | 65 | 10 | 10 | 40 | 40 | 8 | 10 | 10 | 20 | 2 | 10 | 5 | 12 |
| 2021-05-03 | 41 | 50 | 119 | 137 | 20 | 20 | 61 | 65 | 10 | 10 | 40 | 40 | 9 | 10 | 13 | 20 | 2 | 10 | 5 | 12 |
| 2021-05-04 | 41 | 50 | 107 | 133 | 20 | 20 | 62 | 65 | 10 | 10 | 35 | 35 | 8 | 10 | 5 | 20 | 3 | 10 | 5 | 12 |
| 2021-05-05 | 41 | 50 | 101 | 133 | 20 | 20 | 55 | 65 | 10 | 10 | 33 | 36 | 8 | 10 | 8 | 20 | 3 | 10 | 5 | 12 |
| 2021-05-06 | 41 | 50 | 97 | 133 | 20 | 20 | 55 | 65 | 10 | 10 | 26 | 36 | 8 | 10 | 10 | 20 | 3 | 10 | 6 | 12 |
| 2021-05-07 | 41 | 50 | 86 | 133 | 20 | 20 | 44 | 65 | 10 | 10 | 25 | 36 | 8 | 10 | 11 | 20 | 3 | 10 | 6 | 12 |
| 2021-05-08 | 39 | 50 | 97 | 133 | 18 | 20 | 52 | 65 | 10 | 10 | 28 | 36 | 8 | 10 | 10 | 20 | 3 | 10 | 7 | 12 |
| 2021-05-09 | 42 | 50 | 97 | 133 | 20 | 20 | 55 | 65 | 10 | 10 | 27 | 36 | 9 | 10 | 9 | 20 | 3 | 10 | 6 | 12 |
| 2021-05-10 | 44 | 50 | 103 | 133 | 20 | 20 | 57 | 65 | 10 | 10 | 29 | 36 | 10 | 10 | 10 | 20 | 4 | 10 | 7 | 12 |
| 2021-05-11 | 45 | 50 | 103 | 133 | 20 | 20 | 59 | 65 | 10 | 10 | 30 | 36 | 10 | 10 | 7 | 20 | 5 | 10 | 7 | 12 |
| 2021-05-12 | 42 | 50 | 88 | 133 | 18 | 20 | 49 | 65 | 10 | 10 | 29 | 36 | 9 | 10 | 6 | 20 | 5 | 10 | 4 | 12 |
| 2021-05-13 | 42 | 50 | 76 | 133 | 19 | 20 | 35 | 65 | 10 | 10 | 28 | 36 | 8 | 10 | 8 | 20 | 5 | 10 | 5 | 12 |
| 2021-05-14 | 37 | 52 | 84 | 129 | 16 | 20 | 44 | 63 | 10 | 10 | 28 | 36 | 7 | 10 | 8 | 20 | 4 | 12 | 4 | 10 |
| 2021-05-15 | 42 | 50 | 75 | 131 | 20 | 20 | 38 | 63 | 10 | 10 | 25 | 36 | 6 | 10 | 8 | 20 | 6 | 10 | 4 | 12 |
| 2021-05-16 | 41 | 50 | 83 | 131 | 19 | 20 | 48 | 63 | 10 | 10 | 23 | 36 | 6 | 10 | 8 | 20 | 6 | 10 | 4 | 12 |
| 2021-05-17 | 37 | 50 | 76 | 131 | 19 | 20 | 39 | 63 | 10 | 10 | 23 | 36 | 6 | 10 | 7 | 20 | 2 | 10 | 7 | 12 |
| 2021-05-18 | 36 | 50 | 96 | 131 | 20 | 20 | 60 | 63 | 10 | 10 | 21 | 36 | 5 | 10 | 7 | 20 | 1 | 10 | 8 | 12 |
| 2021-05-19 | 36 | 50 | 85 | 131 | 20 | 20 | 60 | 63 | 10 | 10 | 18 | 36 | 5 | 10 | 4 | 20 | 1 | 10 | 3 | 12 |
| 2021-05-20 | 38 | 50 | 90 | 131 | 20 | 20 | 63 | 63 | 10 | 10 | 19 | 36 | 6 | 10 | 6 | 20 | 2 | 10 | 2 | 12 |
| 2021-05-21 | 41 | 50 | 93 | 131 | 20 | 20 | 61 | 63 | 10 | 10 | 21 | 36 | 6 | 10 | 8 | 20 | 5 | 10 | 3 | 12 |
| 2021-05-22 | 40 | 50 | 94 | 131 | 20 | 20 | 60 | 63 | 10 | 10 | 20 | 36 | 5 | 10 | 10 | 20 | 5 | 10 | 4 | 12 |
| 2021-05-23 | 41 | 50 | 97 | 131 | 20 | 20 | 60 | 63 | 10 | 10 | 21 | 36 | 5 | 10 | 12 | 20 | 6 | 10 | 4 | 12 |
| 2021-05-24 | 42 | 50 | 100 | 131 | 20 | 20 | 62 | 63 | 10 | 10 | 23 | 36 | 5 | 10 | 12 | 20 | 7 | 10 | 3 | 12 |
| 2021-05-25 | 42 | 50 | 97 | 131 | 20 | 20 | 61 | 63 | 8 | 10 | 22 | 36 | 5 | 10 | 10 | 20 | 9 | 10 | 4 | 12 |
| 2021-05-26 | 41 | 50 | 99 | 131 | 20 | 20 | 60 | 63 | 8 | 10 | 25 | 36 | 5 | 10 | 9 | 20 | 8 | 10 | 5 | 12 |
| 2021-05-27 | 42 | 50 | 97 | 131 | 20 | 20 | 56 | 63 | 8 | 10 | 24 | 36 | 6 | 10 | 12 | 20 | 8 | 10 | 5 | 12 |
| 2021-05-28 | 43 | 50 | 95 | 131 | 20 | 20 | 48 | 63 | 9 | 10 | 30 | 36 | 6 | 10 | 11 | 20 | 8 | 10 | 6 | 12 |
| 2021-05-29 | 45 | 50 | 92 | 131 | 20 | 20 | 43 | 63 | 10 | 10 | 30 | 36 | 7 | 10 | 10 | 20 | 8 | 10 | 9 | 12 |
| 2021-05-30 | 44 | 50 | 100 | 131 | 20 | 20 | 50 | 63 | 10 | 10 | 32 | 36 | 7 | 10 | 8 | 20 | 7 | 10 | 10 | 12 |
| 2021-05-31 | 44 | 50 | 97 | 131 | 20 | 20 | 52 | 63 | 10 | 10 | 30 | 36 | 7 | 10 | 8 | 20 | 7 | 10 | 7 | 12 |
| 2021-06-01 | 43 | 50 | 107 | 131 | 19 | 20 | 60 | 63 | 10 | 10 | 33 | 36 | 6 | 10 | 7 | 20 | 8 | 10 | 7 | 12 |
| 2021-06-02 | 42 | 50 | 96 | 131 | 20 | 20 | 53 | 63 | 10 | 10 | 30 | 36 | 5 | 10 | 7 | 20 | 7 | 10 | 6 | 12 |
| 2021-06-03 | 42 | 50 | 91 | 131 | 19 | 20 | 51 | 63 | 10 | 10 | 32 | 36 | 5 | 10 | 4 | 20 | 8 | 10 | 4 | 12 |
| 2021-06-04 | 42 | 50 | 74 | 131 | 20 | 20 | 36 | 63 | 10 | 10 | 31 | 36 | 4 | 10 | 4 | 20 | 8 | 10 | 3 | 12 |

| | | | | | | | | | | | | | | | | | | | | |
|---|---|---|---|---|---|---|---|---|---|---|---|---|---|---|---|---|---|---|---|---|
| 2021-06-05 | 41 | 50 | 73 | 131 | 19 | 20 | 39 | 63 | 10 | 10 | 28 | 36 | 4 | 10 | 3 | 20 | 8 | 10 | 3 | 12 |
| 2021-06-06 | 39 | 50 | 67 | 131 | 18 | 20 | 38 | 63 | 10 | 10 | 23 | 36 | 4 | 10 | 5 | 20 | 7 | 10 | 1 | 12 |
| 2021-06-07 | 41 | 50 | 59 | 131 | 19 | 20 | 31 | 63 | 10 | 10 | 22 | 36 | 4 | 10 | 4 | 20 | 8 | 10 | 2 | 12 |
| 2021-06-08 | 31 | 50 | 72 | 131 | 11 | 20 | 41 | 63 | 10 | 10 | 18 | 36 | 3 | 10 | 8 | 20 | 7 | 10 | 5 | 12 |
| 2021-06-09 | 31 | 50 | 68 | 131 | 11 | 20 | 41 | 63 | 10 | 10 | 15 | 36 | 3 | 10 | 7 | 20 | 7 | 10 | 5 | 12 |
| 2021-06-10 | 35 | 50 | 56 | 131 | 13 | 20 | 27 | 63 | 10 | 10 | 15 | 36 | 7 | 10 | 8 | 20 | 5 | 10 | 6 | 12 |
| 2021-06-11 | 32 | 50 | 55 | 131 | 16 | 20 | 25 | 63 | 9 | 10 | 12 | 36 | 3 | 10 | 11 | 20 | 4 | 10 | 7 | 12 |
| 2021-06-12 | 32 | 50 | 48 | 131 | 18 | 20 | 17 | 63 | 8 | 10 | 11 | 36 | 3 | 10 | 12 | 20 | 3 | 10 | 8 | 12 |
| 2021-06-13 | 30 | 50 | 48 | 131 | 20 | 20 | 16 | 63 | 5 | 10 | 11 | 36 | 2 | 10 | 13 | 20 | 3 | 10 | 8 | 12 |
| 2021-06-14 | 29 | 50 | 39 | 131 | 19 | 20 | 10 | 63 | 5 | 10 | 10 | 36 | 2 | 10 | 13 | 20 | 3 | 10 | 6 | 12 |
| 2021-06-15 | 30 | 50 | 45 | 113 | 20 | 20 | 18 | 63 | 5 | 10 | 9 | 18 | 2 | 10 | 13 | 20 | 3 | 10 | 5 | 12 |
| 2021-06-16 | 28 | 50 | 48 | 113 | 19 | 20 | 20 | 63 | 5 | 10 | 9 | 18 | 2 | 10 | 13 | 20 | 2 | 10 | 6 | 12 |
| 2021-06-17 | 31 | 50 | 48 | 122 | 16 | 20 | 24 | 63 | 5 | 10 | 12 | 18 | 8 | 10 | 8 | 29 | 2 | 10 | 4 | 12 |
| 2021-06-18 | 26 | 50 | 43 | 113 | 15 | 20 | 21 | 63 | 7 | 10 | 11 | 18 | 1 | 10 | 8 | 20 | 3 | 10 | 3 | 12 |
| 2021-06-19 | 25 | 50 | 40 | 113 | 13 | 20 | 24 | 63 | 6 | 10 | 7 | 18 | 3 | 10 | 6 | 20 | 3 | 10 | 3 | 12 |
| 2021-06-20 | 26 | 50 | 40 | 113 | 15 | 20 | 25 | 63 | 6 | 10 | 9 | 18 | 2 | 10 | 4 | 20 | 3 | 10 | 2 | 12 |
| 2021-06-21 | 22 | 50 | 37 | 113 | 13 | 20 | 21 | 63 | 6 | 10 | 9 | 18 | 0 | 10 | 5 | 20 | 3 | 10 | 2 | 12 |
| 2021-06-22 | 28 | 50 | 42 | 113 | 17 | 20 | 27 | 63 | 6 | 10 | 9 | 18 | 2 | 10 | 4 | 20 | 3 | 10 | 2 | 12 |
| 2021-06-23 | 21 | 50 | 38 | 113 | 14 | 20 | 26 | 63 | 3 | 10 | 7 | 18 | 2 | 10 | 4 | 20 | 2 | 10 | 1 | 12 |
| 2021-06-24 | 22 | 50 | 33 | 113 | 15 | 20 | 22 | 63 | 3 | 10 | 4 | 18 | 2 | 10 | 6 | 20 | 2 | 10 | 1 | 12 |
| 2021-06-25 | 20 | 46 | 34 | 113 | 14 | 20 | 21 | 63 | 3 | 10 | 3 | 18 | 2 | 6 | 8 | 20 | 1 | 10 | 2 | 12 |
| 2021-06-26 | 21 | 46 | 32 | 97 | 15 | 20 | 20 | 47 | 3 | 10 | 3 | 18 | 2 | 6 | 7 | 20 | 1 | 10 | 2 | 12 |
| 2021-06-27 | 23 | 46 | 35 | 97 | 16 | 20 | 22 | 47 | 4 | 10 | 5 | 18 | 2 | 6 | 7 | 20 | 1 | 10 | 1 | 12 |
| 2021-06-28 | 22 | 46 | 30 | 97 | 15 | 20 | 20 | 47 | 5 | 10 | 4 | 18 | 1 | 6 | 6 | 20 | 1 | 10 | 0 | 12 |
| 2021-06-29 | 20 | 46 | 26 | 97 | 16 | 20 | 16 | 47 | 2 | 10 | 5 | 18 | 1 | 6 | 5 | 20 | 1 | 10 | 0 | 12 |
| 2021-06-30 | 25 | 46 | 33 | 97 | 17 | 20 | 20 | 47 | 3 | 10 | 8 | 18 | 1 | 6 | 5 | 20 | 4 | 10 | 0 | 12 |
| 2021-07-01 | 25 | 46 | 35 | 97 | 17 | 20 | 23 | 47 | 2 | 10 | 9 | 18 | 1 | 6 | 3 | 20 | 5 | 10 | 0 | 12 |
| 2021-07-02 | 24 | 46 | 43 | 97 | 15 | 20 | 29 | 47 | 4 | 10 | 11 | 18 | 2 | 6 | 3 | 20 | 3 | 10 | 0 | 12 |
| 2021-07-03 | 23 | 46 | 39 | 97 | 15 | 20 | 27 | 47 | 3 | 10 | 9 | 18 | 2 | 6 | 3 | 20 | 3 | 10 | 0 | 12 |
| 2021-07-04 | 23 | 46 | 39 | 97 | 16 | 20 | 25 | 47 | 3 | 10 | 11 | 18 | 2 | 6 | 3 | 20 | 2 | 10 | 0 | 12 |
| 2021-07-05 | 22 | 46 | 39 | 97 | 16 | 20 | 25 | 47 | 3 | 10 | 11 | 18 | 1 | 6 | 3 | 20 | 2 | 10 | 0 | 12 |
| 2021-07-06 | 21 | 46 | 40 | 97 | 18 | 20 | 24 | 47 | 3 | 10 | 12 | 18 | 0 | 6 | 3 | 20 | 0 | 10 | 1 | 12 |
| 2021-07-07 | 23 | 46 | 40 | 97 | 17 | 20 | 29 | 47 | 6 | 10 | 6 | 18 | 0 | 6 | 4 | 20 | 0 | 10 | 1 | 12 |
| 2021-07-08 | 25 | 46 | 35 | 97 | 19 | 20 | 22 | 47 | 6 | 10 | 7 | 18 | 0 | 6 | 5 | 20 | 0 | 10 | 1 | 12 |
| 2021-07-09 | 22 | 46 | 48 | 97 | 16 | 20 | 31 | 47 | 6 | 10 | 9 | 18 | 0 | 6 | 6 | 20 | 0 | 10 | 2 | 12 |
| 2021-07-10 | 23 | 46 | 53 | 97 | 16 | 20 | 36 | 47 | 7 | 10 | 8 | 18 | 0 | 6 | 7 | 20 | 0 | 10 | 2 | 12 |
| 2021-07-11 | 25 | 46 | 52 | 97 | 18 | 20 | 35 | 47 | 7 | 10 | 9 | 18 | 0 | 6 | 6 | 20 | 0 | 10 | 2 | 12 |
| 2021-07-12 | 27 | 46 | 51 | 97 | 19 | 20 | 34 | 47 | 7 | 10 | 12 | 18 | 0 | 6 | 4 | 20 | 1 | 10 | 1 | 12 |
| 2021-07-13 | 28 | 46 | 56 | 97 | 20 | 20 | 36 | 47 | 7 | 10 | 14 | 18 | 0 | 6 | 5 | 20 | 1 | 10 | 1 | 12 |
| 2021-07-14 | 24 | 46 | 61 | 97 | 15 | 20 | 41 | 47 | 8 | 10 | 15 | 18 | 0 | 6 | 4 | 20 | 1 | 10 | 1 | 12 |
| 2021-07-15 | 28 | 46 | 57 | 97 | 20 | 20 | 38 | 47 | 7 | 10 | 15 | 18 | 0 | 6 | 3 | 20 | 1 | 10 | 1 | 12 |
| 2021-07-16 | 24 | 46 | 56 | 97 | 17 | 20 | 39 | 47 | 5 | 10 | 14 | 18 | 0 | 6 | 3 | 20 | 2 | 10 | 0 | 12 |

| | | | | | | | | | | | | | | | | | | | | |
|---|---|---|---|---|---|---|---|---|---|---|---|---|---|---|---|---|---|---|---|---|
| 2021-07-17 | 23 | 46 | 59 | 97 | 16 | 20 | 38 | 47 | 4 | 10 | 16 | 18 | 1 | 6 | 5 | 20 | 2 | 10 | 0 | 12 |
| 2021-07-18 | 25 | 46 | 62 | 99 | 19 | 20 | 38 | 47 | 3 | 10 | 20 | 20 | 1 | 6 | 4 | 20 | 2 | 10 | 0 | 12 |
| 2021-07-19 | 27 | 46 | 63 | 100 | 20 | 20 | 39 | 47 | 4 | 10 | 21 | 21 | 1 | 6 | 3 | 20 | 2 | 10 | 0 | 12 |
| 2021-07-20 | 27 | 46 | 64 | 97 | 20 | 20 | 43 | 47 | 5 | 10 | 16 | 18 | 1 | 6 | 4 | 20 | 1 | 10 | 1 | 12 |
| 2021-07-21 | 23 | 46 | 56 | 97 | 19 | 20 | 35 | 47 | 2 | 10 | 14 | 18 | 1 | 6 | 6 | 20 | 1 | 10 | 1 | 12 |
| 2021-07-22 | 21 | 46 | 49 | 88 | 17 | 20 | 31 | 47 | 2 | 10 | 11 | 18 | 1 | 6 | 6 | 11 | 1 | 10 | 1 | 12 |
| 2021-07-23 | 17 | 46 | 50 | 88 | 13 | 20 | 33 | 47 | 2 | 10 | 10 | 18 | 1 | 6 | 6 | 11 | 1 | 10 | 1 | 12 |
| 2021-07-24 | 20 | 46 | 45 | 88 | 14 | 20 | 31 | 47 | 3 | 10 | 10 | 18 | 2 | 6 | 4 | 11 | 1 | 10 | 0 | 12 |
| 2021-07-25 | 25 | 46 | 52 | 88 | 16 | 20 | 38 | 47 | 4 | 10 | 10 | 18 | 3 | 6 | 3 | 11 | 2 | 10 | 1 | 12 |
| 2021-07-26 | 24 | 46 | 52 | 88 | 14 | 20 | 37 | 47 | 5 | 10 | 11 | 18 | 3 | 6 | 3 | 11 | 2 | 10 | 1 | 12 |
| 2021-07-27 | 29 | 46 | 56 | 88 | 20 | 20 | 43 | 47 | 4 | 10 | 9 | 18 | 3 | 6 | 2 | 11 | 2 | 10 | 2 | 12 |
| 2021-07-28 | 28 | 46 | 52 | 88 | 19 | 20 | 36 | 47 | 4 | 10 | 11 | 18 | 3 | 6 | 2 | 11 | 2 | 10 | 3 | 12 |
| 2021-07-29 | 25 | 46 | 50 | 88 | 18 | 20 | 31 | 47 | 5 | 10 | 11 | 18 | 2 | 6 | 2 | 11 | 0 | 10 | 6 | 12 |
| 2021-07-30 | 21 | 46 | 49 | 88 | 11 | 20 | 37 | 47 | 6 | 10 | 7 | 18 | 2 | 6 | 2 | 11 | 2 | 10 | 3 | 12 |
| 2021-07-31 | 23 | 46 | 46 | 88 | 13 | 20 | 31 | 47 | 6 | 10 | 7 | 18 | 2 | 6 | 4 | 11 | 2 | 10 | 4 | 12 |
| 2021-08-01 | 25 | 46 | 48 | 88 | 13 | 20 | 32 | 47 | 7 | 10 | 10 | 18 | 2 | 6 | 2 | 11 | 3 | 10 | 4 | 12 |
| 2021-08-02 | 26 | 46 | 48 | 88 | 14 | 20 | 32 | 47 | 6 | 10 | 9 | 18 | 4 | 6 | 4 | 11 | 2 | 10 | 3 | 12 |
| 2021-08-03 | 25 | 46 | 51 | 88 | 14 | 20 | 35 | 47 | 6 | 10 | 9 | 18 | 3 | 6 | 5 | 11 | 2 | 10 | 2 | 12 |
| 2021-08-04 | 19 | 46 | 55 | 88 | 11 | 20 | 37 | 47 | 4 | 10 | 11 | 18 | 2 | 6 | 5 | 11 | 2 | 10 | 2 | 12 |
| 2021-08-05 | 18 | 46 | 52 | 88 | 13 | 20 | 32 | 47 | 3 | 10 | 12 | 18 | 1 | 6 | 7 | 11 | 1 | 10 | 1 | 12 |
| 2021-08-06 | 21 | 46 | 54 | 88 | 13 | 20 | 34 | 47 | 3 | 10 | 13 | 18 | 3 | 6 | 6 | 11 | 2 | 10 | 1 | 12 |
| 2021-08-07 | 23 | 46 | 42 | 88 | 15 | 20 | 27 | 47 | 3 | 10 | 10 | 18 | 4 | 6 | 5 | 11 | 1 | 10 | 0 | 12 |
| 2021-08-08 | 27 | 46 | 44 | 88 | 19 | 20 | 28 | 47 | 2 | 10 | 9 | 18 | 3 | 6 | 7 | 11 | 3 | 10 | 0 | 12 |
| 2021-08-09 | 30 | 46 | 42 | 88 | 20 | 20 | 27 | 47 | 4 | 10 | 9 | 18 | 3 | 6 | 6 | 11 | 3 | 10 | 0 | 12 |
| 2021-08-10 | 28 | 46 | 47 | 88 | 19 | 20 | 30 | 47 | 4 | 10 | 11 | 18 | 2 | 6 | 6 | 11 | 3 | 10 | 0 | 12 |
| 2021-08-11 | 26 | 46 | 46 | 88 | 17 | 20 | 26 | 47 | 4 | 10 | 14 | 18 | 2 | 6 | 6 | 11 | 3 | 10 | 0 | 12 |
| 2021-08-12 | 28 | 46 | 43 | 88 | 19 | 20 | 23 | 47 | 6 | 10 | 12 | 18 | 1 | 6 | 7 | 11 | 2 | 10 | 1 | 12 |
| 2021-08-13 | 27 | 46 | 53 | 90 | 18 | 20 | 25 | 47 | 6 | 10 | 20 | 20 | 2 | 6 | 7 | 11 | 1 | 10 | 1 | 12 |
| 2021-08-14 | 26 | 46 | 52 | 90 | 17 | 20 | 24 | 47 | 7 | 10 | 20 | 20 | 2 | 6 | 7 | 11 | 0 | 10 | 1 | 12 |
| 2021-08-15 | 30 | 46 | 50 | 88 | 20 | 20 | 25 | 47 | 7 | 10 | 16 | 18 | 2 | 6 | 6 | 11 | 1 | 10 | 3 | 12 |
| 2021-08-16 | 30 | 46 | 48 | 88 | 20 | 20 | 27 | 47 | 9 | 10 | 12 | 18 | 1 | 6 | 6 | 11 | 0 | 10 | 3 | 12 |
| 2021-08-17 | 29 | 46 | 53 | 88 | 20 | 20 | 30 | 47 | 7 | 10 | 13 | 18 | 0 | 6 | 7 | 11 | 2 | 10 | 3 | 12 |
| 2021-08-18 | 28 | 46 | 50 | 88 | 20 | 20 | 27 | 47 | 7 | 10 | 16 | 18 | 0 | 6 | 5 | 11 | 1 | 10 | 2 | 12 |
| 2021-08-19 | 28 | 46 | 46 | 95 | 19 | 20 | 21 | 47 | 7 | 10 | 20 | 25 | 1 | 6 | 4 | 11 | 1 | 10 | 1 | 12 |
| 2021-08-20 | 30 | 46 | 51 | 95 | 20 | 20 | 25 | 47 | 8 | 10 | 19 | 25 | 1 | 6 | 5 | 11 | 1 | 10 | 2 | 12 |
| 2021-08-21 | 30 | 46 | 52 | 95 | 18 | 20 | 23 | 47 | 8 | 10 | 22 | 25 | 3 | 6 | 5 | 11 | 1 | 10 | 2 | 12 |
| 2021-08-22 | 33 | 46 | 50 | 95 | 20 | 20 | 24 | 47 | 8 | 10 | 19 | 25 | 3 | 6 | 5 | 11 | 2 | 10 | 2 | 12 |
| 2021-08-23 | 34 | 46 | 44 | 95 | 20 | 20 | 22 | 47 | 5 | 10 | 16 | 25 | 5 | 6 | 3 | 11 | 4 | 10 | 3 | 12 |
| 2021-08-24 | 36 | 46 | 48 | 95 | 20 | 20 | 27 | 47 | 8 | 10 | 14 | 25 | 4 | 6 | 5 | 11 | 4 | 10 | 2 | 12 |
| 2021-08-25 | 29 | 46 | 39 | 95 | 16 | 20 | 20 | 47 | 6 | 10 | 11 | 25 | 3 | 6 | 6 | 11 | 4 | 10 | 2 | 12 |
| 2021-08-26 | 30 | 46 | 39 | 95 | 16 | 20 | 18 | 47 | 7 | 10 | 13 | 25 | 3 | 6 | 5 | 11 | 4 | 10 | 3 | 12 |
| 2021-08-27 | 27 | 46 | 43 | 95 | 13 | 20 | 20 | 47 | 8 | 10 | 13 | 25 | 3 | 6 | 6 | 11 | 3 | 10 | 4 | 12 |

| | | | | | | | | | | | | | | | | | | | | |
|---|---|---|---|---|---|---|---|---|---|---|---|---|---|---|---|---|---|---|---|---|
| 2021-08-28 | 25 | 46 | 41 | 95 | 14 | 20 | 20 | 47 | 6 | 10 | 13 | 25 | 3 | 6 | 3 | 11 | 2 | 10 | 5 | 12 |
| 2021-08-29 | 23 | 46 | 35 | 95 | 12 | 20 | 20 | 47 | 6 | 10 | 12 | 25 | 3 | 6 | 1 | 11 | 2 | 10 | 2 | 12 |
| 2021-08-30 | 17 | 46 | 32 | 95 | 9 | 20 | 19 | 47 | 6 | 10 | 9 | 25 | 2 | 6 | 1 | 11 | 0 | 10 | 3 | 12 |
| 2021-08-31 | 16 | 46 | 36 | 95 | 10 | 20 | 20 | 47 | 4 | 10 | 12 | 25 | 2 | 6 | 1 | 11 | 0 | 10 | 3 | 12 |
| 2021-09-01 | 15 | 46 | 33 | 88 | 8 | 20 | 24 | 47 | 5 | 10 | 6 | 18 | 2 | 6 | 1 | 11 | 0 | 10 | 2 | 12 |
| 2021-09-02 | 18 | 46 | 27 | 88 | 11 | 20 | 16 | 47 | 5 | 10 | 6 | 18 | 2 | 6 | 3 | 11 | 0 | 10 | 2 | 12 |
| 2021-09-03 | 18 | 46 | 25 | 88 | 9 | 20 | 14 | 47 | 7 | 10 | 5 | 18 | 2 | 6 | 4 | 11 | 0 | 10 | 2 | 12 |
| 2021-09-04 | 18 | 46 | 25 | 88 | 9 | 20 | 13 | 47 | 6 | 10 | 6 | 18 | 2 | 6 | 4 | 11 | 1 | 10 | 2 | 12 |
| 2021-09-05 | 17 | 46 | 22 | 88 | 8 | 20 | 14 | 47 | 6 | 10 | 3 | 18 | 2 | 6 | 4 | 11 | 1 | 10 | 1 | 12 |
| 2021-09-06 | 17 | 46 | 26 | 88 | 8 | 20 | 16 | 47 | 6 | 10 | 6 | 18 | 3 | 6 | 3 | 11 | 0 | 10 | 1 | 12 |
| 2021-09-07 | 18 | 46 | 33 | 88 | 10 | 20 | 23 | 47 | 5 | 10 | 7 | 18 | 2 | 6 | 2 | 11 | 1 | 10 | 1 | 12 |
| 2021-09-08 | 17 | 46 | 32 | 88 | 8 | 20 | 23 | 47 | 5 | 10 | 7 | 18 | 2 | 6 | 1 | 11 | 2 | 10 | 1 | 12 |
| 2021-09-09 | 16 | 46 | 31 | 88 | 9 | 20 | 20 | 47 | 5 | 10 | 8 | 18 | 0 | 6 | 2 | 11 | 2 | 10 | 1 | 12 |
| 2021-09-10 | 15 | 46 | 27 | 88 | 8 | 20 | 18 | 47 | 5 | 10 | 6 | 18 | 0 | 6 | 3 | 11 | 2 | 10 | 0 | 12 |
| 2021-09-11 | 15 | 46 | 26 | 88 | 8 | 20 | 18 | 47 | 5 | 10 | 6 | 18 | 0 | 6 | 2 | 11 | 2 | 10 | 0 | 12 |
| 2021-09-12 | 13 | 46 | 24 | 88 | 5 | 20 | 16 | 47 | 6 | 10 | 6 | 18 | 0 | 6 | 2 | 11 | 2 | 10 | 0 | 12 |
| 2021-09-13 | 13 | 46 | 24 | 88 | 3 | 20 | 16 | 47 | 8 | 10 | 7 | 18 | 0 | 6 | 1 | 11 | 2 | 10 | 0 | 12 |
| 2021-09-14 | 16 | 46 | 24 | 88 | 6 | 20 | 15 | 47 | 8 | 10 | 7 | 18 | 0 | 6 | 1 | 11 | 2 | 10 | 1 | 12 |
| 2021-09-15 | 16 | 46 | 18 | 88 | 6 | 20 | 10 | 47 | 8 | 10 | 6 | 18 | 0 | 6 | 1 | 11 | 2 | 10 | 1 | 12 |
| 2021-09-16 | 16 | 46 | 20 | 88 | 6 | 20 | 12 | 47 | 8 | 10 | 6 | 18 | 0 | 6 | 1 | 11 | 2 | 10 | 1 | 12 |
| 2021-09-17 | 17 | 46 | 19 | 88 | 7 | 20 | 13 | 47 | 8 | 10 | 5 | 18 | 1 | 6 | 1 | 11 | 1 | 10 | 0 | 12 |
| 2021-09-18 | 13 | 46 | 21 | 79 | 4 | 20 | 14 | 47 | 7 | 10 | 6 | 9 | 1 | 6 | 1 | 11 | 1 | 10 | 0 | 12 |
| 2021-09-19 | 16 | 46 | 18 | 79 | 4 | 20 | 13 | 47 | 9 | 10 | 4 | 9 | 2 | 6 | 1 | 11 | 1 | 10 | 0 | 12 |
| 2021-09-20 | 13 | 46 | 19 | 79 | 4 | 20 | 12 | 47 | 8 | 10 | 6 | 9 | 1 | 6 | 1 | 11 | 0 | 10 | 0 | 12 |
| 2021-09-21 | 13 | 46 | 22 | 60 | 5 | 20 | 13 | 28 | 6 | 10 | 8 | 9 | 2 | 6 | 0 | 11 | 0 | 10 | 1 | 12 |
| 2021-09-22 | 13 | 46 | 24 | 60 | 5 | 20 | 15 | 28 | 6 | 10 | 8 | 9 | 2 | 6 | 0 | 11 | 0 | 10 | 1 | 12 |
| 2021-09-23 | 13 | 46 | 26 | 60 | 5 | 20 | 16 | 28 | 6 | 10 | 8 | 9 | 2 | 6 | 1 | 11 | 0 | 10 | 1 | 12 |
| 2021-09-24 | 10 | 51 | 28 | 59 | 4 | 20 | 15 | 28 | 4 | 10 | 9 | 13 | 2 | 11 | 3 | 6 | 0 | 10 | 1 | 12 |
| 2021-09-25 | 11 | 46 | 23 | 64 | 4 | 20 | 15 | 28 | 5 | 10 | 8 | 13 | 2 | 6 | 0 | 11 | 0 | 10 | 0 | 12 |
| 2021-09-26 | 12 | 46 | 23 | 64 | 4 | 20 | 15 | 28 | 5 | 10 | 9 | 13 | 3 | 6 | 0 | 11 | 0 | 10 | 1 | 12 |
| 2021-09-27 | 16 | 46 | 23 | 64 | 4 | 20 | 15 | 28 | 6 | 10 | 8 | 13 | 5 | 6 | 0 | 11 | 1 | 10 | 0 | 12 |
| 2021-09-28 | 15 | 46 | 19 | 64 | 4 | 20 | 13 | 28 | 5 | 10 | 5 | 13 | 5 | 6 | 1 | 11 | 1 | 10 | 0 | 12 |
| 2021-09-29 | 10 | 45 | 16 | 60 | 4 | 20 | 11 | 28 | 3 | 10 | 4 | 9 | 2 | 5 | 1 | 11 | 1 | 10 | 0 | 12 |
| 2021-10-01 | 14 | 45 | 12 | 60 | 7 | 20 | 7 | 28 | 2 | 10 | 3 | 9 | 4 | 5 | 2 | 11 | 1 | 10 | 0 | 12 |
| 2021-10-02 | 15 | 45 | 11 | 60 | 8 | 20 | 6 | 28 | 2 | 10 | 3 | 9 | 4 | 5 | 2 | 11 | 1 | 10 | 0 | 12 |
| 2021-10-03 | 16 | 45 | 8 | 60 | 8 | 20 | 6 | 28 | 3 | 10 | 2 | 9 | 4 | 5 | 2 | 11 | 1 | 10 | 0 | 12 |
| 2021-10-04 | 16 | 45 | 8 | 60 | 8 | 20 | 4 | 28 | 3 | 10 | 3 | 9 | 4 | 5 | 1 | 11 | 1 | 10 | 0 | 12 |
| 2021-10-05 | 10 | 45 | 10 | 60 | 5 | 20 | 4 | 28 | 2 | 10 | 5 | 9 | 2 | 5 | 0 | 11 | 1 | 10 | 1 | 12 |
| 2021-10-06 | 6 | 45 | 10 | 60 | 3 | 20 | 5 | 28 | 2 | 10 | 4 | 9 | 1 | 5 | 0 | 11 | 0 | 10 | 1 | 12 |
| 2021-10-07 | 5 | 45 | 12 | 60 | 3 | 20 | 6 | 28 | 2 | 10 | 6 | 9 | 0 | 5 | 0 | 11 | 0 | 10 | 0 | 12 |
| 2021-10-08 | 5 | 45 | 13 | 60 | 3 | 20 | 7 | 28 | 1 | 10 | 6 | 9 | 0 | 5 | 0 | 11 | 1 | 10 | 0 | 12 |
| 2021-10-09 | 8 | 45 | 11 | 43 | 4 | 20 | 7 | 11 | 1 | 10 | 4 | 9 | 2 | 5 | 0 | 11 | 1 | 10 | 0 | 12 |

| | | | | | | | | | | | | | | | | | | | | |
|---|---|---|---|---|---|---|---|---|---|---|---|---|---|---|---|---|---|---|---|---|
| 2021-10-10 | 8 | 45 | 10 | 43 | 4 | 20 | 6 | 11 | 1 | 10 | 4 | 9 | 2 | 5 | 0 | 11 | 1 | 10 | 0 | 12 |
| 2021-10-11 | 6 | 45 | 12 | 43 | 4 | 20 | 8 | 11 | 1 | 10 | 4 | 9 | 0 | 5 | 0 | 11 | 1 | 10 | 0 | 12 |
| 2021-10-12 | 10 | 45 | 10 | 43 | 3 | 20 | 8 | 11 | 4 | 10 | 2 | 9 | 2 | 5 | 0 | 11 | 1 | 10 | 0 | 12 |
| 2021-10-13 | 9 | 45 | 7 | 43 | 3 | 20 | 5 | 11 | 5 | 10 | 2 | 9 | 0 | 5 | 0 | 11 | 1 | 10 | 0 | 12 |
| 2021-10-14 | 6 | 45 | 7 | 43 | 3 | 20 | 5 | 11 | 2 | 10 | 2 | 9 | 0 | 5 | 0 | 11 | 1 | 10 | 0 | 12 |
| 2021-10-15 | 6 | 45 | 6 | 43 | 3 | 20 | 4 | 11 | 2 | 10 | 2 | 9 | 0 | 5 | 0 | 11 | 1 | 10 | 0 | 12 |
| 2021-10-16 | 6 | 45 | 6 | 43 | 3 | 20 | 3 | 11 | 2 | 10 | 3 | 9 | 0 | 5 | 0 | 11 | 1 | 10 | 0 | 12 |
| 2021-10-17 | 5 | 45 | 5 | 43 | 3 | 20 | 3 | 11 | 1 | 10 | 2 | 9 | 0 | 5 | 0 | 11 | 1 | 10 | 0 | 12 |
| 2021-10-18 | 4 | 45 | 4 | 43 | 2 | 20 | 2 | 11 | 1 | 10 | 2 | 9 | 0 | 5 | 0 | 11 | 1 | 10 | 0 | 12 |
| 2021-10-19 | 5 | 45 | 6 | 43 | 2 | 20 | 2 | 11 | 2 | 10 | 3 | 9 | 0 | 5 | 1 | 11 | 1 | 10 | 0 | 12 |
| 2021-10-20 | 3 | 45 | 7 | 43 | 1 | 20 | 1 | 11 | 2 | 10 | 5 | 9 | 0 | 5 | 1 | 11 | 0 | 10 | 0 | 12 |
| 2021-10-21 | 4 | 37 | 2 | 43 | 1 | 20 | 1 | 11 | 3 | 10 | 0 | 9 | 0 | 5 | 1 | 11 | 0 | 2 | 0 | 12 |
| 2021-10-22 | 3 | 37 | 2 | 43 | 1 | 20 | 0 | 11 | 2 | 10 | 1 | 9 | 0 | 5 | 1 | 11 | 0 | 2 | 0 | 12 |
| 2021-10-26 | 2 | 37 | 1 | 43 | 1 | 20 | 1 | 11 | 1 | 10 | 0 | 9 | 0 | 5 | 0 | 11 | 0 | 2 | 0 | 12 |
| 2021-10-27 | 3 | 37 | 0 | 43 | 2 | 20 | 0 | 11 | 1 | 10 | 0 | 9 | 0 | 5 | 0 | 11 | 0 | 2 | 0 | 12 |
| 2021-10-28 | 3 | 37 | 0 | 43 | 2 | 20 | 0 | 11 | 1 | 10 | 0 | 9 | 0 | 5 | 0 | 11 | 0 | 2 | 0 | 12 |
| 2021-10-29 | 3 | 37 | 0 | 43 | 2 | 20 | 0 | 11 | 1 | 10 | 0 | 9 | 0 | 5 | 0 | 11 | 0 | 2 | 0 | 12 |
| 2021-10-30 | 3 | 45 | 0 | 43 | 2 | 20 | 0 | 11 | 1 | 10 | 0 | 9 | 0 | 5 | 0 | 11 | 0 | 10 | 0 | 12 |
| 2021-10-31 | 3 | 37 | 0 | 43 | 2 | 20 | 0 | 11 | 1 | 10 | 0 | 9 | 0 | 5 | 0 | 11 | 0 | 2 | 0 | 12 |
| 2021-11-01 | 3 | 37 | 1 | 39 | 2 | 20 | 1 | 11 | 1 | 2 | 0 | 5 | 0 | 5 | 0 | 11 | 0 | 10 | 0 | 12 |
| 2021-11-02 | 3 | 29 | 1 | 39 | 3 | 20 | 0 | 11 | 0 | 2 | 0 | 5 | 0 | 5 | 1 | 11 | 0 | 2 | 0 | 12 |
| 2021-11-03 | 1 | 29 | 1 | 39 | 1 | 20 | 0 | 11 | 0 | 2 | 0 | 5 | 0 | 5 | 1 | 11 | 0 | 2 | 0 | 12 |
| 2021-11-04 | 1 | 29 | 1 | 39 | 1 | 20 | 0 | 11 | 0 | 2 | 0 | 5 | 0 | 5 | 1 | 11 | 0 | 2 | 0 | 12 |
| 2021-11-05 | 1 | 29 | 1 | 39 | 1 | 20 | 0 | 11 | 0 | 2 | 0 | 5 | 0 | 5 | 1 | 11 | 0 | 2 | 0 | 12 |
| 2021-11-06 | 0 | 29 | 0 | 39 | 0 | 20 | 0 | 11 | 0 | 2 | 0 | 5 | 0 | 5 | 0 | 11 | 0 | 2 | 0 | 12 |
| 2021-11-07 | 0 | 29 | 0 | 39 | 0 | 20 | 0 | 11 | 0 | 2 | 0 | 5 | 0 | 5 | 0 | 11 | 0 | 2 | 0 | 12 |
| 2021-11-08 | 0 | 29 | 0 | 39 | 0 | 20 | 0 | 11 | 0 | 2 | 0 | 5 | 0 | 5 | 0 | 11 | 0 | 2 | 0 | 12 |
| 2021-11-09 | 0 | 39 | 0 | 29 | 0 | 20 | 0 | 11 | 0 | 2 | 0 | 5 | 0 | 5 | 0 | 11 | 0 | 12 | 0 | 2 |
| 2021-11-10 | 0 | 29 | 0 | 39 | 0 | 20 | 0 | 11 | 0 | 2 | 0 | 5 | 0 | 5 | 0 | 11 | 0 | 2 | 0 | 12 |
| 2021-11-11 | 0 | 29 | 2 | 39 | 0 | 20 | 1 | 11 | 0 | 2 | 1 | 5 | 0 | 5 | 0 | 11 | 0 | 2 | 0 | 12 |
| 2021-11-13 | 2 | 29 | 3 | 32 | 0 | 20 | 2 | 11 | 2 | 2 | 1 | 5 | 0 | 5 | 0 | 4 | 0 | 2 | 0 | 12 |
| 2021-11-14 | 0 | 29 | 4 | 32 | 0 | 20 | 2 | 11 | 0 | 2 | 1 | 5 | 0 | 5 | 1 | 4 | 0 | 2 | 0 | 12 |
| 2021-11-15 | 0 | 29 | 4 | 32 | 0 | 20 | 2 | 11 | 0 | 2 | 1 | 5 | 0 | 5 | 1 | 4 | 0 | 2 | 0 | 12 |
| 2021-11-16 | 0 | 29 | 4 | 32 | 0 | 20 | 2 | 11 | 0 | 2 | 1 | 5 | 0 | 5 | 1 | 4 | 0 | 2 | 0 | 12 |
| 2021-11-17 | 0 | 29 | 2 | 32 | 0 | 20 | 2 | 11 | 0 | 2 | 0 | 5 | 0 | 5 | 0 | 4 | 0 | 2 | 0 | 12 |
| 2021-11-18 | 0 | 29 | 5 | 32 | 0 | 20 | 4 | 11 | 0 | 2 | 0 | 5 | 0 | 5 | 1 | 4 | 0 | 2 | 0 | 12 |
| 2021-11-19 | 0 | 29 | 4 | 32 | 0 | 20 | 3 | 11 | 0 | 2 | 0 | 5 | 0 | 5 | 1 | 4 | 0 | 2 | 0 | 12 |
| 2021-11-20 | 0 | 29 | 4 | 32 | 0 | 20 | 3 | 11 | 0 | 2 | 0 | 5 | 0 | 5 | 1 | 4 | 0 | 2 | 0 | 12 |
| 2021-11-21 | 0 | 29 | 4 | 32 | 0 | 20 | 3 | 11 | 0 | 2 | 0 | 5 | 0 | 5 | 1 | 4 | 0 | 2 | 0 | 12 |
| 2021-11-22 | 0 | 29 | 3 | 32 | 0 | 20 | 3 | 11 | 0 | 2 | 0 | 5 | 0 | 5 | 0 | 4 | 0 | 2 | 0 | 12 |
| 2021-11-23 | 0 | 29 | 1 | 32 | 0 | 20 | 1 | 11 | 0 | 5 | 0 | 4 | 0 | 2 | 0 | 5 | 0 | 2 | 0 | 12 |
| 2021-11-24 | 0 | 29 | 2 | 32 | 0 | 20 | 1 | 11 | 0 | 2 | 1 | 5 | 0 | 5 | 0 | 4 | 0 | 2 | 0 | 12 |

| | | | | | | | | | | | | | | | | | | | | |
|---|---|---|---|---|---|---|---|---|---|---|---|---|---|---|---|---|---|---|---|---|
| 2021-11-25 | 0 | 29 | 1 | 32 | 0 | 20 | 1 | 11 | 0 | 2 | 0 | 5 | 0 | 5 | 0 | 4 | 0 | 2 | 0 | 12 |
| 2021-11-26 | 0 | 29 | 1 | 32 | 0 | 20 | 1 | 11 | 0 | 2 | 0 | 5 | 0 | 5 | 0 | 4 | 0 | 2 | 0 | 12 |
| 2021-11-27 | 0 | 29 | 1 | 32 | 0 | 20 | 0 | 11 | 0 | 2 | 1 | 5 | 0 | 5 | 0 | 4 | 0 | 2 | 0 | 12 |
| 2021-11-28 | 0 | 29 | 1 | 32 | 0 | 20 | 0 | 11 | 0 | 2 | 1 | 5 | 0 | 5 | 0 | 4 | 0 | 2 | 0 | 12 |
| 2021-11-29 | 0 | 29 | 0 | 32 | 0 | 20 | 0 | 11 | 0 | 2 | 0 | 5 | 0 | 5 | 0 | 4 | 0 | 2 | 0 | 12 |
| 2021-11-30 | 0 | 29 | 1 | 32 | 0 | 20 | 0 | 11 | 0 | 2 | 0 | 5 | 0 | 5 | 1 | 4 | 0 | 2 | 0 | 12 |
| 2021-12-01 | 0 | 29 | 1 | 32 | 0 | 20 | 0 | 11 | 0 | 2 | 0 | 5 | 0 | 5 | 1 | 4 | 0 | 2 | 0 | 12 |
| 2021-12-02 | 0 | 29 | 2 | 32 | 0 | 20 | 1 | 11 | 0 | 2 | 0 | 5 | 0 | 5 | 1 | 4 | 0 | 2 | 0 | 12 |
| 2021-12-03 | 0 | 29 | 2 | 32 | 0 | 20 | 1 | 11 | 0 | 2 | 0 | 5 | 0 | 5 | 1 | 4 | 0 | 2 | 0 | 12 |
| 2021-12-04 | 0 | 29 | 2 | 32 | 0 | 20 | 1 | 11 | 0 | 2 | 0 | 5 | 0 | 5 | 1 | 4 | 0 | 2 | 0 | 12 |
| 2021-12-05 | 0 | 29 | 2 | 32 | 0 | 20 | 1 | 11 | 0 | 2 | 0 | 5 | 0 | 5 | 1 | 4 | 0 | 2 | 0 | 12 |
| 2021-12-06 | 0 | 29 | 2 | 32 | 0 | 20 | 1 | 11 | 0 | 2 | 0 | 5 | 0 | 5 | 1 | 4 | 0 | 2 | 0 | 12 |
| 2021-12-07 | 0 | 29 | 2 | 32 | 0 | 20 | 1 | 11 | 0 | 2 | 0 | 5 | 0 | 5 | 1 | 4 | 0 | 2 | 0 | 12 |
| 2021-12-08 | 0 | 29 | 2 | 32 | 0 | 20 | 1 | 11 | 0 | 2 | 0 | 5 | 0 | 5 | 1 | 4 | 0 | 2 | 0 | 12 |
| 2021-12-09 | 0 | 29 | 1 | 32 | 0 | 20 | 0 | 11 | 0 | 2 | 0 | 5 | 0 | 5 | 1 | 4 | 0 | 2 | 0 | 12 |
| 2021-12-10 | 0 | 29 | 1 | 27 | 0 | 20 | 0 | 6 | 0 | 2 | 0 | 5 | 0 | 5 | 1 | 4 | 0 | 2 | 0 | 12 |
| 2021-12-11 | 0 | 28 | 1 | 27 | 0 | 20 | 0 | 6 | 0 | 2 | 0 | 5 | 0 | 4 | 1 | 4 | 0 | 2 | 0 | 12 |
| 2021-12-12 | 0 | 28 | 1 | 27 | 0 | 20 | 0 | 6 | 0 | 2 | 0 | 5 | 0 | 4 | 1 | 4 | 0 | 2 | 0 | 12 |
| 2021-12-13 | 0 | 28 | 1 | 27 | 0 | 20 | 0 | 6 | 0 | 2 | 0 | 5 | 0 | 4 | 1 | 4 | 0 | 2 | 0 | 12 |
| 2021-12-14 | 0 | 28 | 2 | 27 | 0 | 20 | 0 | 6 | 0 | 2 | 0 | 5 | 0 | 4 | 2 | 4 | 0 | 2 | 0 | 12 |
| 2021-12-15 | 0 | 28 | 1 | 27 | 0 | 20 | 0 | 6 | 0 | 2 | 0 | 5 | 0 | 4 | 1 | 4 | 0 | 2 | 0 | 12 |
| 2021-12-16 | 0 | 28 | 1 | 27 | 0 | 20 | 0 | 6 | 0 | 2 | 0 | 5 | 0 | 4 | 1 | 4 | 0 | 2 | 0 | 12 |
| 2021-12-18 | 0 | 28 | 1 | 27 | 0 | 20 | 0 | 6 | 0 | 2 | 0 | 5 | 0 | 4 | 1 | 4 | 0 | 2 | 0 | 12 |
| 2021-12-19 | 0 | 28 | 1 | 27 | 0 | 20 | 0 | 6 | 0 | 2 | 0 | 5 | 0 | 4 | 1 | 4 | 0 | 2 | 0 | 12 |
| 2021-12-20 | 0 | 28 | 1 | 27 | 0 | 20 | 0 | 6 | 0 | 2 | 0 | 5 | 0 | 4 | 1 | 4 | 0 | 2 | 0 | 12 |
| 2021-12-21 | 0 | 28 | 1 | 27 | 0 | 20 | 0 | 6 | 0 | 2 | 0 | 5 | 0 | 4 | 1 | 4 | 0 | 2 | 0 | 12 |
| 2021-12-22 | 0 | 28 | 1 | 27 | 0 | 20 | 0 | 6 | 0 | 2 | 0 | 5 | 0 | 4 | 1 | 4 | 0 | 2 | 0 | 12 |
| 2021-12-23 | 0 | 28 | 1 | 27 | 0 | 20 | 0 | 6 | 0 | 2 | 0 | 5 | 0 | 4 | 1 | 4 | 0 | 2 | 0 | 12 |
| 2021-12-25 | 0 | 28 | 0 | 27 | 0 | 20 | 0 | 6 | 0 | 2 | 0 | 5 | 0 | 4 | 0 | 4 | 0 | 2 | 0 | 12 |
| 2021-12-26 | 0 | 28 | 0 | 27 | 0 | 20 | 0 | 6 | 0 | 2 | 0 | 5 | 0 | 4 | 0 | 4 | 0 | 2 | 0 | 12 |
| 2021-12-27 | 0 | 28 | 1 | 27 | 0 | 20 | 0 | 6 | 0 | 2 | 0 | 5 | 0 | 4 | 1 | 4 | 0 | 2 | 0 | 12 |
| 2021-12-29 | 0 | 28 | 2 | 27 | 0 | 20 | 0 | 6 | 0 | 2 | 1 | 5 | 0 | 4 | 1 | 4 | 0 | 2 | 0 | 12 |
| 2021-12-30 | 0 | 28 | 1 | 27 | 0 | 20 | 0 | 6 | 0 | 2 | 1 | 5 | 0 | 4 | 0 | 4 | 0 | 2 | 0 | 12 |
| 2021-12-31 | 0 | 28 | 0 | 27 | 0 | 20 | 0 | 6 | 0 | 2 | 0 | 5 | 0 | 4 | 0 | 4 | 0 | 2 | 0 | 12 |
| 2022-01-01 | 0 | 28 | 0 | 27 | 0 | 20 | 0 | 6 | 0 | 2 | 0 | 5 | 0 | 4 | 0 | 4 | 0 | 2 | 0 | 12 |
| 2022-01-02 | 0 | 28 | 0 | 27 | 0 | 20 | 0 | 6 | 0 | 2 | 0 | 5 | 0 | 4 | 0 | 4 | 0 | 2 | 0 | 12 |
| 2022-01-03 | 0 | 28 | 0 | 27 | 0 | 20 | 0 | 6 | 0 | 2 | 0 | 5 | 0 | 4 | 0 | 4 | 0 | 2 | 0 | 12 |
| 2022-01-04 | 0 | 28 | 0 | 27 | 0 | 20 | 0 | 6 | 0 | 2 | 0 | 5 | 0 | 4 | 0 | 4 | 0 | 2 | 0 | 12 |
| 2022-01-05 | 0 | 28 | 1 | 27 | 0 | 20 | 1 | 6 | 0 | 2 | 0 | 5 | 0 | 4 | 0 | 4 | 0 | 2 | 0 | 12 |
| 2022-01-06 | 0 | 28 | 1 | 27 | 0 | 20 | 0 | 6 | 0 | 2 | 0 | 5 | 0 | 4 | 1 | 4 | 0 | 2 | 0 | 12 |
| 2022-01-07 | 0 | 28 | 1 | 27 | 0 | 20 | 0 | 6 | 0 | 2 | 0 | 5 | 0 | 4 | 1 | 4 | 0 | 2 | 0 | 12 |
| 2022-01-08 | 0 | 28 | 1 | 27 | 0 | 20 | 0 | 6 | 0 | 2 | 0 | 5 | 0 | 4 | 1 | 4 | 0 | 2 | 0 | 12 |

| | | | | | | | | | | | | | | | | | | | | |
|---|---|---|---|---|---|---|---|---|---|---|---|---|---|---|---|---|---|---|---|---|
| 2022-01-09 | 0 | 28 | 0 | 27 | 0 | 20 | 0 | 6 | 0 | 2 | 0 | 5 | 0 | 4 | 0 | 4 | 0 | 2 | 0 | 12 |
| 2022-01-10 | 0 | 28 | 5 | 27 | 0 | 20 | 3 | 6 | 0 | 2 | 2 | 5 | 0 | 4 | 0 | 4 | 0 | 2 | 0 | 12 |
| 2022-01-11 | 0 | 28 | 5 | 27 | 0 | 20 | 3 | 6 | 0 | 2 | 2 | 5 | 0 | 4 | 0 | 4 | 0 | 2 | 0 | 12 |
| 2022-01-12 | 0 | 28 | 10 | 27 | 0 | 20 | 6 | 6 | 0 | 2 | 2 | 5 | 0 | 4 | 0 | 4 | 0 | 2 | 2 | 12 |
| 2022-01-13 | 0 | 28 | 9 | 32 | 0 | 20 | 7 | 11 | 0 | 2 | 2 | 5 | 0 | 4 | 0 | 4 | 0 | 2 | 0 | 12 |
| 2022-01-14 | 2 | 28 | 13 | 32 | 0 | 20 | 8 | 11 | 0 | 2 | 2 | 5 | 0 | 4 | 2 | 4 | 2 | 2 | 1 | 12 |
| 2022-01-15 | 2 | 28 | 17 | 32 | 1 | 20 | 10 | 11 | 1 | 2 | 5 | 5 | 0 | 4 | 2 | 4 | 0 | 2 | 0 | 12 |
| 2022-01-16 | 1 | 28 | 17 | 32 | 0 | 20 | 11 | 11 | 1 | 2 | 5 | 5 | 0 | 4 | 1 | 4 | 0 | 2 | 0 | 12 |
| 2022-01-17 | 1 | 28 | 15 | 32 | 0 | 20 | 9 | 11 | 1 | 2 | 4 | 5 | 0 | 4 | 2 | 4 | 0 | 2 | 0 | 12 |
| 2022-01-18 | 3 | 28 | 17 | 33 | 0 | 20 | 10 | 11 | 1 | 2 | 6 | 6 | 1 | 4 | 1 | 4 | 1 | 2 | 0 | 12 |
| 2022-01-19 | 4 | 28 | 28 | 45 | 1 | 20 | 20 | 23 | 1 | 2 | 6 | 6 | 1 | 4 | 2 | 4 | 1 | 2 | 0 | 12 |
| 2022-01-20 | 6 | 28 | 24 | 44 | 1 | 20 | 17 | 23 | 2 | 2 | 5 | 5 | 1 | 4 | 2 | 4 | 2 | 2 | 0 | 12 |
| 2022-01-21 | 6 | 28 | 33 | 44 | 2 | 20 | 21 | 23 | 2 | 2 | 8 | 8 | 0 | 4 | 3 | 4 | 2 | 2 | 1 | 12 |
| 2022-01-22 | 11 | 31 | 36 | 55 | 7 | 20 | 22 | 23 | 2 | 5 | 10 | 16 | 0 | 4 | 2 | 4 | 2 | 2 | 2 | 12 |
| 2022-01-23 | 12 | 21 | 36 | 55 | 8 | 10 | 23 | 23 | 2 | 5 | 9 | 16 | 0 | 4 | 2 | 4 | 2 | 2 | 2 | 12 |
| 2022-01-24 | 12 | 21 | 43 | 55 | 8 | 10 | 23 | 23 | 3 | 5 | 13 | 16 | 0 | 4 | 4 | 4 | 1 | 2 | 3 | 12 |
| 2022-01-25 | 11 | 21 | 47 | 56 | 7 | 10 | 22 | 23 | 4 | 5 | 16 | 16 | 0 | 4 | 5 | 5 | 0 | 2 | 4 | 12 |
| 2022-01-26 | 13 | 25 | 48 | 62 | 7 | 10 | 23 | 23 | 3 | 5 | 16 | 16 | 0 | 4 | 6 | 11 | 3 | 6 | 3 | 12 |
| 2022-01-27 | 12 | 25 | 58 | 87 | 7 | 10 | 31 | 44 | 3 | 5 | 18 | 20 | 0 | 4 | 6 | 11 | 2 | 6 | 3 | 12 |
| 2022-01-28 | 16 | 23 | 59 | 87 | 10 | 10 | 30 | 44 | 3 | 5 | 20 | 20 | 1 | 4 | 6 | 11 | 2 | 4 | 3 | 12 |
| 2022-01-29 | 18 | 23 | 62 | 87 | 9 | 10 | 34 | 44 | 5 | 5 | 20 | 20 | 1 | 4 | 8 | 11 | 3 | 4 | 0 | 12 |
| 2022-01-30 | 15 | 23 | 63 | 87 | 8 | 10 | 33 | 44 | 5 | 5 | 20 | 20 | 1 | 4 | 9 | 11 | 1 | 4 | 1 | 12 |
| 2022-01-31 | 15 | 23 | 65 | 87 | 7 | 10 | 35 | 44 | 4 | 5 | 20 | 20 | 1 | 4 | 10 | 11 | 3 | 4 | 0 | 12 |
| 2022-02-01 | 13 | 19 | 65 | 75 | 7 | 10 | 35 | 44 | 5 | 5 | 20 | 20 | 1 | 4 | 10 | 11 | - | 0 | - | 0 |
| 2022-02-02 | 14 | 23 | 61 | 87 | 6 | 10 | 30 | 44 | 5 | 5 | 20 | 20 | 1 | 4 | 10 | 11 | 2 | 4 | 1 | 12 |
| 2022-02-04 | 13 | 23 | 53 | 87 | 7 | 10 | 28 | 44 | 5 | 5 | 19 | 20 | 1 | 4 | 6 | 11 | 0 | 4 | 0 | 12 |
| 2022-02-05 | 14 | 23 | 53 | 87 | 7 | 10 | 28 | 44 | 5 | 5 | 20 | 20 | 1 | 4 | 4 | 11 | 1 | 4 | 1 | 12 |
| 2022-02-06 | 15 | 23 | 51 | 87 | 9 | 10 | 28 | 44 | 4 | 5 | 20 | 20 | 2 | 4 | 4 | 11 | 0 | 4 | 1 | 12 |
| 2022-02-07 | 17 | 23 | 48 | 87 | 9 | 10 | 29 | 44 | 5 | 5 | 15 | 20 | 3 | 4 | 4 | 11 | 0 | 4 | 0 | 12 |
| 2022-02-08 | 17 | 23 | 55 | 87 | 9 | 10 | 35 | 44 | 5 | 5 | 12 | 20 | 3 | 4 | 7 | 11 | 0 | 4 | 1 | 12 |
| 2022-02-09 | 15 | 23 | 59 | 87 | 7 | 10 | 34 | 44 | 5 | 5 | 16 | 20 | 3 | 4 | 8 | 11 | 0 | 4 | 1 | 12 |
| 2022-02-10 | 13 | 23 | 46 | 87 | 6 | 10 | 29 | 44 | 5 | 5 | 9 | 20 | 2 | 4 | 7 | 11 | 0 | 4 | 1 | 12 |
| 2022-02-11 | 15 | 23 | 51 | 87 | 7 | 10 | 32 | 44 | 5 | 5 | 10 | 20 | 3 | 4 | 8 | 11 | 0 | 4 | 1 | 12 |
| 2022-02-12 | 13 | 23 | 48 | 87 | 7 | 10 | 31 | 44 | 3 | 5 | 11 | 20 | 2 | 4 | 6 | 11 | 1 | 4 | 0 | 12 |
| 2022-02-13 | 14 | 23 | 42 | 87 | 8 | 10 | 28 | 44 | 3 | 5 | 10 | 20 | 2 | 4 | 4 | 11 | 1 | 4 | 0 | 12 |
| 2022-02-14 | 13 | 23 | 35 | 87 | 6 | 10 | 24 | 44 | 4 | 5 | 8 | 20 | 2 | 4 | 3 | 11 | 1 | 4 | 0 | 12 |
| 2022-02-15 | 8 | 23 | 35 | 87 | 4 | 10 | 24 | 44 | 2 | 5 | 10 | 20 | 2 | 4 | 1 | 11 | 0 | 4 | 0 | 12 |
| 2022-02-16 | 9 | 23 | 35 | 87 | 4 | 10 | 24 | 44 | 3 | 5 | 10 | 20 | 2 | 4 | 1 | 11 | 0 | 4 | 0 | 12 |
| 2022-02-17 | 10 | 23 | 31 | 87 | 5 | 10 | 25 | 44 | 3 | 5 | 5 | 20 | 2 | 4 | 1 | 11 | 0 | 4 | 0 | 12 |
| 2022-02-18 | 11 | 23 | 24 | 87 | 6 | 10 | 21 | 44 | 3 | 5 | 3 | 20 | 2 | 4 | 0 | 11 | 0 | 4 | 0 | 12 |
| 2022-02-19 | 10 | 23 | 27 | 75 | 6 | 10 | 23 | 44 | 3 | 5 | 3 | 8 | 1 | 4 | 1 | 11 | 0 | 4 | 0 | 12 |
| 2022-02-20 | 12 | 23 | 23 | 75 | 6 | 10 | 21 | 44 | 2 | 5 | 1 | 8 | 4 | 4 | 1 | 11 | 0 | 4 | 0 | 12 |

| | | | | | | | | | | | | | | | | | | | | |
|---|---|---|---|---|---|---|---|---|---|---|---|---|---|---|---|---|---|---|---|---|
| 2022-02-21 | 10 | 23 | 23 | 75 | 6 | 10 | 20 | 44 | 1 | 5 | 1 | 8 | 3 | 4 | 2 | 11 | 0 | 4 | 0 | 12 |
| 2022-02-22 | 5 | 23 | 23 | 75 | 2 | 10 | 21 | 44 | 1 | 5 | 1 | 8 | 2 | 4 | 1 | 11 | 0 | 4 | 0 | 12 |
| 2022-02-23 | 4 | 23 | 8 | 43 | 3 | 10 | 6 | 12 | 0 | 5 | 1 | 8 | 1 | 4 | 1 | 11 | 0 | 4 | 0 | 12 |
| 2022-02-24 | 3 | 23 | 6 | 43 | 3 | 10 | 5 | 12 | 0 | 5 | 1 | 8 | 0 | 4 | 0 | 11 | 0 | 4 | 0 | 12 |
| 2022-02-25 | 5 | 23 | 3 | 43 | 5 | 10 | 3 | 12 | 0 | 5 | 0 | 8 | 0 | 4 | 0 | 11 | 0 | 4 | 3 | 12 |
| 2022-02-26 | 3 | 23 | 3 | 43 | 3 | 10 | 3 | 12 | 0 | 5 | 0 | 8 | 0 | 4 | 0 | 11 | 0 | 4 | 0 | 12 |
| 2022-02-27 | 3 | 23 | 3 | 43 | 3 | 10 | 2 | 12 | 0 | 5 | 1 | 8 | 0 | 4 | 0 | 11 | 0 | 4 | 0 | 12 |
| 2022-02-28 | 2 | 23 | 2 | 43 | 2 | 10 | 1 | 12 | 0 | 5 | 1 | 8 | 0 | 4 | 0 | 11 | 0 | 4 | 0 | 12 |
| 2022-03-01 | 1 | 23 | 1 | 43 | 1 | 10 | 1 | 12 | 0 | 5 | 0 | 8 | 0 | 4 | 0 | 11 | 0 | 4 | 0 | 12 |
| 2022-03-02 | 1 | 23 | 2 | 43 | 1 | 10 | 1 | 12 | 0 | 5 | 1 | 8 | 0 | 4 | 0 | 11 | 0 | 4 | 0 | 12 |
| 2022-03-03 | 2 | 23 | 3 | 43 | 1 | 10 | 2 | 12 | 1 | 5 | 1 | 8 | 0 | 4 | 0 | 11 | 0 | 4 | 0 | 12 |
| 2022-03-04 | 2 | 23 | 4 | 43 | 1 | 10 | 2 | 12 | 1 | 5 | 2 | 8 | 0 | 4 | 0 | 11 | 0 | 4 | 0 | 12 |
| 2022-03-05 | 2 | 23 | 4 | 43 | 2 | 10 | 1 | 12 | 0 | 5 | 2 | 8 | 0 | 4 | 1 | 11 | 0 | 4 | 0 | 12 |
| 2022-03-06 | 2 | 23 | 5 | 43 | 2 | 10 | 1 | 12 | 0 | 5 | 3 | 8 | 0 | 4 | 1 | 11 | 0 | 4 | 0 | 12 |
| 2022-03-07 | 1 | 23 | 2 | 43 | 1 | 10 | 1 | 12 | 0 | 5 | 1 | 8 | 0 | 4 | 0 | 11 | 0 | 4 | 0 | 12 |
| 2022-03-08 | 3 | 23 | 2 | 43 | 2 | 10 | 1 | 12 | 1 | 5 | 1 | 8 | 0 | 4 | 0 | 11 | 0 | 4 | 0 | 12 |
| 2022-03-09 | 2 | 23 | 3 | 43 | 2 | 10 | 1 | 12 | 0 | 5 | 2 | 8 | 0 | 4 | 0 | 11 | 0 | 4 | 0 | 12 |
| 2022-03-10 | 2 | 23 | 3 | 43 | 2 | 10 | 1 | 12 | 0 | 5 | 2 | 8 | 0 | 4 | 0 | 11 | 0 | 4 | 0 | 12 |
| 2022-03-11 | 2 | 23 | 1 | 43 | 2 | 10 | 1 | 12 | 0 | 5 | 0 | 8 | 0 | 4 | 0 | 11 | 0 | 4 | 0 | 12 |
| 2022-03-12 | 0 | 23 | 3 | 39 | 0 | 10 | 1 | 12 | 0 | 5 | 1 | 4 | 0 | 4 | 0 | 11 | 0 | 4 | 1 | 12 |
| 2022-03-13 | 0 | 23 | 3 | 39 | 0 | 10 | 1 | 12 | 0 | 5 | 1 | 4 | 0 | 4 | 0 | 11 | 0 | 4 | 1 | 12 |
| 2022-03-14 | 0 | 23 | 3 | 39 | 0 | 10 | 1 | 12 | 0 | 5 | 1 | 4 | 0 | 4 | 0 | 11 | 0 | 4 | 1 | 12 |
| 2022-03-15 | 0 | 23 | 3 | 39 | 0 | 10 | 1 | 12 | 0 | 5 | 1 | 4 | 0 | 4 | 0 | 11 | 0 | 4 | 1 | 12 |
| 2022-03-16 | 0 | 23 | 3 | 39 | 0 | 10 | 1 | 12 | 0 | 5 | 1 | 4 | 0 | 4 | 0 | 11 | 0 | 4 | 1 | 12 |
| 2022-03-17 | 0 | 23 | 3 | 39 | 0 | 10 | 1 | 12 | 0 | 5 | 1 | 4 | 0 | 4 | 0 | 11 | 0 | 4 | 1 | 12 |
| 2022-03-18 | 0 | 23 | 3 | 29 | 0 | 10 | 1 | 2 | 0 | 5 | 1 | 4 | 0 | 4 | 0 | 11 | 0 | 4 | 1 | 12 |
| 2022-03-19 | 0 | 23 | 1 | 29 | 0 | 10 | 1 | 2 | 0 | 5 | 0 | 4 | 0 | 4 | 0 | 11 | 0 | 4 | 0 | 12 |
| 2022-03-20 | 0 | 23 | 1 | 29 | 0 | 10 | 1 | 2 | 0 | 5 | 0 | 4 | 0 | 4 | 0 | 11 | 0 | 4 | 0 | 12 |
| 2022-03-21 | 0 | 23 | 1 | 29 | 0 | 10 | 1 | 2 | 0 | 5 | 0 | 4 | 0 | 4 | 0 | 11 | 0 | 4 | 0 | 12 |
| 2022-03-22 | 0 | 23 | 0 | 29 | 0 | 10 | 0 | 2 | 0 | 5 | 0 | 4 | 0 | 4 | 0 | 11 | 0 | 4 | 0 | 12 |
| 2022-03-23 | 0 | 23 | 0 | 29 | 0 | 10 | 0 | 2 | 0 | 5 | 0 | 4 | 0 | 4 | 0 | 11 | 0 | 4 | 0 | 12 |
| 2022-03-24 | 0 | 23 | 0 | 29 | 0 | 10 | 0 | 2 | 0 | 5 | 0 | 4 | 0 | 4 | 0 | 11 | 0 | 4 | 0 | 12 |
| 2022-03-25 | 0 | 23 | 0 | 29 | 0 | 10 | 0 | 2 | 0 | 5 | 0 | 4 | 0 | 4 | 0 | 11 | 0 | 4 | 0 | 12 |
| 2022-03-26 | 0 | 23 | 0 | 29 | 0 | 10 | 0 | 2 | 0 | 5 | 0 | 4 | 0 | 4 | 0 | 11 | 0 | 4 | 0 | 12 |
| 2022-03-27 | 0 | 23 | 0 | 29 | 0 | 10 | 0 | 2 | 0 | 5 | 0 | 4 | 0 | 4 | 0 | 11 | 0 | 4 | 0 | 12 |
| 2022-03-28 | 1 | 23 | 1 | 29 | 0 | 10 | 0 | 2 | 1 | 5 | 0 | 4 | 0 | 4 | 0 | 11 | 0 | 4 | 1 | 12 |
| 2022-03-29 | 1 | 23 | 0 | 29 | 0 | 10 | 0 | 2 | 1 | 5 | 0 | 4 | 0 | 4 | 0 | 11 | 0 | 4 | 0 | 12 |
| 2022-03-30 | 1 | 23 | 0 | 29 | 0 | 10 | 0 | 2 | 1 | 5 | 0 | 4 | 0 | 4 | 0 | 11 | 0 | 4 | 0 | 12 |
| 2022-03-31 | 1 | 23 | 1 | 29 | 0 | 10 | 1 | 2 | 1 | 5 | 0 | 4 | 0 | 4 | 0 | 11 | 0 | 4 | 0 | 12 |
| 2022-04-01 | 1 | 23 | 2 | 29 | 0 | 10 | 1 | 2 | 1 | 5 | 0 | 4 | 0 | 4 | 1 | 11 | 0 | 4 | 0 | 12 |
| 2022-04-02 | 1 | 29 | 1 | 29 | 0 | 2 | 1 | 2 | 1 | 4 | 0 | 4 | 0 | 11 | 0 | 11 | 0 | 12 | 0 | 12 |

### 4 Database table of vaccination progress in Nova Friburgo

| Date | VACCINE | | | | | |
|---|---|---|---|---|---|---|
| | Unique dose | First dose | Second dose | Booster dose | Booster Janssen | ≥11 |
| 2021-02-15 | | 5635 | 996 | | | |
| 2021-02-17 | | 5782 | 1163 | | | |
| 2021-02-18 | | 5821 | 1562 | | | |
| 2021-02-19 | | 5883 | 1640 | | | |
| 2021-02-23 | | 5929 | 1640 | | | |
| 2021-02-24 | | 6019 | 1808 | | | |
| 2021-02-25 | | 6043 | 1855 | | | |
| 2021-02-26 | | 6155 | 1872 | | | |
| 2021-03-01 | | 7195 | 1872 | | | |
| 2021-03-03 | | 7422 | 1971 | | | |
| 2021-03-05 | | 7423 | 3035 | | | |
| 2021-03-08 | | 8399 | 3433 | | | |
| 2021-03-09 | | 8749 | 3659 | | | |
| 2021-03-10 | | 8769 | 3679 | | | |
| 2021-03-11 | | 8776 | 3717 | | | |
| 2021-03-15 | | 9494 | 3805 | | | |
| 2021-03-16 | | 9944 | 3829 | | | |
| 2021-03-17 | | 9981 | 3866 | | | |
| 2021-03-18 | | 10013 | 3899 | | | |
| 2021-03-19 | | 10058 | 3990 | | | |
| 2021-03-22 | | 12034 | 3999 | | | |
| 2021-03-23 | | 13340 | 4230 | | | |
| 2021-03-29 | | 19509 | 4244 | | | |
| 2021-03-30 | | 21392 | 4316 | | | |
| 2021-03-31 | | 21525 | 5051 | | | |
| 2021-04-05 | | 22982 | 5449 | | | |
| 2021-04-06 | | 23545 | 5474 | | | |
| 2021-04-07 | | 23552 | 6338 | | | |
| 2021-04-12 | | 25424 | 6936 | | | |
| 2021-04-13 | | 26688 | 7046 | | | |

| | | | |
|---|---|---|---|
| 2021-04-14 | | 26770 | 8319 |
| 2021-04-15 | | 26773 | 8830 |
| 2021-04-17 | | 26782 | 11192 |
| 2021-04-19 | | 28708 | 12490 |
| 2021-04-20 | | 29939 | 13354 |
| 2021-04-21 | | 29951 | 15341 |
| 2021-04-23 | | 30077 | 15936 |
| 2021-04-26 | | 32546 | 16461 |
| 2021-04-27 | | 33706 | 16643 |
| 2021-04-28 | | 33706 | 17596 |
| 2021-04-29 | | 33926 | 17596 |
| 2021-05-03 | | 37524 | 18963 |
| 2021-05-04 | | 39738 | 19786 |
| 2021-05-05 | | 40162 | 19787 |
| 2021-05-06 | | 40829 | 19876 |
| 2021-05-10 | | 41156 | 20191 |
| 2021-05-11 | | 41802 | 20191 |
| 2021-05-12 | | 41802 | 20393 |
| 2021-05-13 | | 41806 | 20448 |
| 2021-05-17 | | 44108 | 20451 |
| 2021-05-18 | | 45147 | 20478 |
| 2021-05-20 | | 46000 | 22034 |
| 2021-05-24 | | 48999 | 22052 |
| 2021-05-25 | | 50629 | 22052 |
| 2021-05-26 | | 51140 | 22052 |
| 2021-05-27 | | 51159 | 22055 |
| 2021-06-02 | | 53829 | 22679 |
| 2021-06-04 | | 54351 | 22679 |
| 2021-06-09 | | 57246 | 22786 |
| 2021-06-10 | | 57264 | 22875 |
| 2021-06-14 | | 60594 | 22888 |
| 2021-06-15 | | 62032 | 22895 |
| 2021-06-16 | | 62396 | 22919 |
| 2021-06-21 | | 62985 | 22920 |
| 2021-06-22 | | 67060 | 22931 |
| 2021-06-23 | | 67135 | 22969 |
| 2021-06-25 | | 67135 | 22969 |
| 2021-06-28 | | 68783 | 22969 |
| 2021-06-29 | | 70802 | 22973 |
| 2021-06-30 | | 71326 | 23736 |
| 2021-07-01 | | 71508 | 24443 |
| 2021-07-05 | | 74994 | 24462 |

| | | | | |
|---|---|---|---|---|
| 2021-07-06 | | 76451 | 24807 | |
| 2021-07-07 | | 76517 | 26711 | |
| 2021-07-08 | | 76654 | 27438 | |
| 2021-07-10 | 1570 | 78196 | 27573 | |
| 2021-07-12 | 1570 | 82302 | 27580 | |
| 2021-07-13 | 3718 | 83979 | 27655 | |
| 2021-07-14 | 3822 | 86128 | 29357 | |
| 2021-07-15 | 3833 | 88141 | 29965 | |
| 2021-07-16 | 3913 | 88147 | 30177 | |
| 2021-07-19 | 3954 | 90944 | 30276 | |
| 2021-07-20 | 3995 | 94672 | 30350 | |
| 2021-07-21 | 3996 | 96572 | 33394 | |
| 2021-07-22 | 4214 | 98106 | 35637 | |
| 2021-07-25 | 4214 | 98106 | 35637 | |
| 2021-07-26 | 4214 | 101226 | 35644 | |
| 2021-07-27 | 4239 | 104329 | 35673 | |
| 2021-07-28 | 4239 | 104330 | 36816 | |
| 2021-07-29 | 4239 | 104330 | 37328 | |
| 2021-08-02 | 4246 | 106664 | 40035 | |
| 2021-08-04 | 4246 | 110403 | 41751 | |
| 2021-08-09 | 4246 | 113574 | 41814 | |
| 2021-08-10 | 4246 | 117248 | 44088 | |
| 2021-08-12 | 4246 | 117248 | 46602 | |
| 2021-08-13 | 4246 | 117248 | 47873 | |
| 2021-08-16 | 4246 | 120683 | 48103 | |
| 2021-08-17 | 4440 | 124323 | 48150 | |
| 2021-08-18 | 4440 | 126496 | 48244 | |
| 2021-08-19 | 4440 | 128655 | 49554 | |
| 2021-08-21 | 4440 | 132757 | 50674 | |
| 2021-08-25 | 4520 | 134395 | 57002 | |
| 2021-08-26 | 4520 | 134395 | 57370 | |
| 2021-08-30 | 4520 | 134411 | 58182 | |
| 2021-09-01 | 4360 | 134417 | 62931 | |
| 2021-09-02 | 4360 | 134577 | 64763 | |
| 2021-09-08 | 4360 | 135579 | 67835 | |
| 2021-09-09 | 4360 | 135614 | 68293 | |
| 2021-09-10 | 4360 | 139384 | 68293 | |
| 2021-09-13 | 4360 | 143500 | 68294 | |
| 2021-09-14 | 4360 | 147818 | 68294 | 253 |
| 2021-09-15 | 4360 | 147925 | 71383 | 253 |
| 2021-09-17 | 4360 | 149842 | 75221 | 253 |
| 2021-09-21 | 4360 | 149845 | 76539 | 253 |

| | | | | | |
|---|---|---|---|---|---|
| 2021-09-23 | 4360 | 149857 | 80195 | 253 | |
| 2021-09-24 | 4360 | 149880 | 82077 | 253 | |
| 2021-09-28 | 4360 | 151207 | 82213 | 253 | |
| 2021-09-29 | 4360 | 151222 | 88120 | 254 | |
| 2021-10-01 | 4360 | 151237 | 88841 | 253 | |
| 2021-10-06 | 4360 | 151286 | 95135 | 439 | |
| 2021-10-07 | 4360 | 151335 | 98917 | 456 | |
| 2021-10-13 | 4360 | 151395 | 102644 | 1170 | |
| 2021-10-15 | 4360 | 151403 | 108654 | 1898 | |
| 2021-10-18 | 4360 | 152331 | 114597 | 2773 | |
| 2021-10-19 | 4360 | 152350 | 116346 | 3052 | |
| 2021-10-22 | 4021 | 152546 | 117301 | 4021 | |
| 2021-10-25 | 4360 | 152586 | 119390 | 5164 | |
| 2021-10-26 | 4360 | 152648 | 122462 | 6119 | |
| 2021-11-04 | 4360 | 153711 | 124555 | 12383 | |
| 2021-11-05 | 4360 | 153905 | 125163 | 13214 | |
| 2021-11-08 | 4360 | 153941 | 127656 | 13775 | |
| 2021-11-09 | 4360 | 153950 | 130705 | 14593 | |
| 2021-11-10 | 4360 | 153977 | 131191 | 15280 | |
| 2021-11-11 | 4360 | 154017 | 131563 | 15916 | |
| 2021-11-12 | 4360 | 154105 | 133347 | 16040 | |
| 2021-11-16 | 4360 | 154133 | 133862 | 16374 | |
| 2021-11-18 | 4360 | 154227 | 134405 | 17082 | |
| 2021-11-23 | 4360 | 154306 | 135002 | 18028 | |
| 2021-11-24 | 4360 | 154362 | 135370 | 18591 | |
| 2021-11-25 | 4360 | 154452 | 135585 | 18767 | |
| 2021-11-30 | 4360 | 154472 | 137390 | 18798 | |
| 2021-12-01 | 4360 | 154501 | 140970 | 18806 | |
| 2021-12-02 | 4360 | 154538 | 145070 | 18817 | |
| 2021-12-03 | 4360 | 154711 | 146355 | 18817 | |
| 2021-12-07 | 4360 | 154815 | 147558 | 20675 | |
| 2021-12-08 | 4360 | 154844 | 148184 | 22117 | |
| 2021-12-09 | 4360 | 154907 | 148603 | 22242 | |
| 2021-12-13 | 4360 | 154956 | 148912 | 24534 | |
| 2021-12-16 | 4360 | 155055 | 149526 | 27921 | |
| 2021-12-20 | 4360 | 155113 | 149988 | 30827 | |
| 2022-01-04 | 4360 | 155235 | 150520 | 36448 | 1583 |
| 2022-01-05 | 4360 | 155269 | 150804 | 40076 | 1583 |
| 2022-01-06 | 4360 | 155314 | 150970 | 40337 | 2065 |
| 2022-01-11 | 4360 | 155355 | 151209 | 43601 | 2065 |
| 2022-01-12 | 4360 | 155413 | 151279 | 43601 | 2378 |
| 2022-01-18 | 4360 | 155537 | 151961 | 50119 | 2378 |

| | | | | | | |
|---|---|---|---|---|---|---|
| 2022-01-19 | 4360 | 155603 | 152057 | 50124 | 2612 | |
| 2022-01-20 | 4360 | 155644 | 152337 | 52526 | 2612 | |
| 2022-01-25 | 4360 | 155682 | 152553 | 55585 | 2612 | |
| 2022-02-03 | | | | | | 2270 |
| 2022-02-08 | 4360 | 155933 | 154272 | 64107 | 2612 | |
| 2022-02-09 | | | | | | 4044 |
| 2022-02-10 | | | | | | 5077 |
| 2022-02-11 | 4360 | 156021 | 154451 | 64112 | 2612 | |
| 2022-02-15 | 4360 | 156054 | 154631 | 67665 | 2612 | 5083 |
| 2022-02-17 | 4360 | 156054 | 154632 | 67665 | 2612 | 8221 |
| 2022-02-18 | 4360 | 156090 | 154728 | 67685 | 2612 | |
| 2022-02-22 | 4360 | 156119 | 154891 | 71082 | 2612 | |
| 2022-02-23 | 4360 | 156122 | 154892 | 71161 | 2612 | |
| 2022-02-24 | | | | | | 9641 |
| 2022-02-25 | 4360 | 156150 | 154951 | 71174 | 2612 | |
| 2022-03-04 | 4360 | 156182 | 155061 | 72421 | 2612 | 9926 |
| 2022-03-12 | 4360 | 156188 | 155139 | 74112 | 2612 | 11591 |

### 5 Database table of community mobility in Nova Friburgo

| Date | Retail_and_recreation_percent_change_from_baseline | Grocery_and_pharmacy_percent_change_from_baseline | Parks_percent_change_from_baseline | Transit_stations_percent_change_from_baseline | Workplaces_percent_change_from_baseline | Residential_percent_change_from_baseline |
|---|---|---|---|---|---|---|
| 2020-03-29 | -71 | -46 | -73 | -79 | -44 | 17 |
| 2020-03-30 | -65 | -39 | -65 | -73 | -56 | 23 |
| 2020-03-31 | -62 | -29 | -42 | -67 | -56 | 22 |
| 2020-04-01 | -62 | -30 | -65 | -67 | -55 | 23 |
| 2020-04-02 | -59 | -25 | -57 | -67 | -53 | 22 |
| 2020-04-03 | -61 | -26 | -64 | -66 | -51 | 24 |
| 2020-04-04 | -65 | -27 | -73 | -63 | -45 | 18 |
| 2020-04-05 | -65 | -36 | -70 | -71 | -40 | 16 |
| 2020-04-06 | -52 | -22 | -60 | -63 | -44 | 19 |
| 2020-04-07 | -52 | -19 | -60 | -60 | -46 | 19 |
| 2020-04-08 | -49 | -11 | -64 | -61 | -45 | 19 |
| 2020-04-09 | -47 | 2 | -54 | -56 | -46 | 18 |
| 2020-04-10 | -75 | -41 | -75 | -79 | -73 | 29 |
| 2020-04-11 | -66 | -27 | -77 | -71 | -45 | 19 |
| 2020-04-12 | -64 | -29 | -74 | -73 | -33 | 15 |

| | | | | | | |
|---|---|---|---|---|---|---|
| 2020-04-13 | -51 | -25 | -47 | -68 | -41 | 18 |
| 2020-04-14 | -55 | -26 | -40 | -61 | -43 | 19 |
| 2020-04-15 | -52 | -21 | -56 | -63 | -42 | 19 |
| 2020-04-16 | -54 | -22 | -49 | -66 | -41 | 20 |
| 2020-04-17 | -54 | -20 | -50 | -64 | -40 | 20 |
| 2020-04-18 | -61 | -23 | -67 | -67 | -37 | 17 |
| 2020-04-19 | -60 | -26 | -58 | -65 | -33 | 15 |
| 2020-04-20 | -48 | -21 | -48 | -58 | -39 | 17 |
| 2020-04-21 | -68 | -43 | -55 | -75 | -65 | 23 |
| 2020-04-22 | -45 | -13 | -38 | -60 | -40 | 18 |
| 2020-04-23 | -64 | -32 | -60 | -71 | -58 | 23 |
| 2020-04-24 | -49 | -13 | -47 | -59 | -37 | 19 |
| 2020-04-25 | -57 | -20 | -61 | -61 | -31 | 15 |
| 2020-04-26 | -58 | -24 | -55 | -66 | -32 | 14 |
| 2020-04-27 | -50 | -24 | -54 | -61 | -36 | 17 |
| 2020-04-28 | -51 | -20 | -54 | -61 | -38 | 18 |
| 2020-04-29 | -47 | -16 | -52 | -61 | -38 | 18 |
| 2020-04-30 | -42 | 8 | -47 | -53 | -36 | 16 |
| 2020-05-01 | -69 | -64 | -53 | -72 | -69 | 27 |
| 2020-05-02 | -57 | -1 | -63 | -59 | -33 | 15 |
| 2020-05-03 | -61 | -16 | -67 | -71 | -32 | 14 |
| 2020-05-04 | -51 | -5 | -47 | -61 | -34 | 16 |
| 2020-05-05 | -51 | -6 | -57 | -59 | -37 | 17 |
| 2020-05-06 | -47 | 0 | -49 | -60 | -35 | 17 |
| 2020-05-07 | -51 | 0 | -53 | -62 | -37 | 18 |
| 2020-05-08 | -44 | 7 | -45 | -52 | -32 | 18 |
| 2020-05-09 | -48 | 5 | -59 | -57 | -26 | 14 |
| 2020-05-10 | -51 | -4 | -64 | -65 | -10 | 10 |
| 2020-05-11 | -48 | -12 | -46 | -63 | -33 | 16 |
| 2020-05-12 | -51 | -12 | -49 | -62 | -37 | 17 |
| 2020-05-13 | -49 | -10 | -54 | -61 | -36 | 18 |
| 2020-05-14 | -50 | -9 | -49 | -61 | -36 | 17 |
| 2020-05-15 | -54 | -11 | -60 | -63 | -35 | 20 |
| 2020-05-16 | -64 | -21 | -72 | -67 | -40 | 18 |
| 2020-05-17 | -56 | -19 | -56 | -67 | -23 | 14 |
| 2020-05-18 | -46 | -11 | -43 | -59 | -32 | 16 |
| 2020-05-19 | -50 | -11 | -47 | -58 | -36 | 17 |
| 2020-05-20 | -46 | -6 | -48 | -53 | -35 | 18 |
| 2020-05-21 | -49 | -6 | -39 | -58 | -35 | 18 |
| 2020-05-22 | -48 | -5 | -44 | -51 | -31 | 19 |
| 2020-05-23 | -53 | -11 | -63 | -55 | -25 | 15 |
| 2020-05-24 | -62 | -25 | -72 | -65 | -29 | 15 |

| | | | | | | |
|---|---|---|---|---|---|---|
| 2020-05-25 | -48 | -9 | -48 | -59 | -32 | 16 |
| 2020-05-26 | -48 | -10 | -51 | -62 | -33 | 16 |
| 2020-05-27 | -44 | -2 | -48 | -56 | -32 | 17 |
| 2020-05-28 | -46 | 0 | -50 | -57 | -31 | 17 |
| 2020-05-29 | -44 | 6 | -45 | -48 | -29 | 17 |
| 2020-05-30 | -49 | -3 | -60 | -55 | -20 | 13 |
| 2020-05-31 | -48 | -7 | -47 | -57 | -12 | 11 |
| 2020-06-01 | -43 | -8 | -44 | -53 | -26 | 14 |
| 2020-06-02 | -43 | -7 | -50 | -47 | -30 | 16 |
| 2020-06-03 | -40 | -3 | -47 | -45 | -28 | 17 |
| 2020-06-04 | -41 | -2 | -48 | -52 | -28 | 16 |
| 2020-06-05 | -39 | 5 | -33 | -45 | -26 | 17 |
| 2020-06-06 | -45 | -3 | -61 | -43 | -18 | 13 |
| 2020-06-07 | -51 | -18 | -57 | -47 | -16 | 13 |
| 2020-06-08 | -38 | -5 | -49 | -40 | -25 | 15 |
| 2020-06-09 | -39 | -3 | -48 | -32 | -28 | 15 |
| 2020-06-10 | -30 | 9 | -42 | -34 | -25 | 14 |
| 2020-06-11 | -56 | -21 | -41 | -51 | -54 | 22 |
| 2020-06-12 | -30 | 15 | -35 | -31 | -30 | 16 |
| 2020-06-13 | -45 | -7 | -46 | -31 | -14 | 12 |
| 2020-06-14 | -47 | -12 | -40 | -38 | -7 | 11 |
| 2020-06-15 | -40 | -10 | -45 | -35 | -25 | 15 |
| 2020-06-16 | -43 | -9 | -51 | -38 | -27 | 16 |
| 2020-06-17 | -37 | -1 | -44 | -28 | -25 | 16 |
| 2020-06-18 | -40 | -3 | -46 | -40 | -25 | 16 |
| 2020-06-19 | -40 | 1 | -41 | -23 | -21 | 16 |
| 2020-06-20 | -45 | -5 | -50 | -27 | -14 | 13 |
| 2020-06-21 | -44 | -9 | -29 | -31 | -5 | 11 |
| 2020-06-22 | -40 | -8 | -42 | -33 | -24 | 14 |
| 2020-06-23 | -42 | -8 | -49 | -35 | -28 | 15 |
| 2020-06-24 | -37 | -1 | -43 | -30 | -24 | 15 |
| 2020-06-25 | -39 | -2 | -34 | -27 | -23 | 15 |
| 2020-06-26 | -39 | 0 | -30 | -24 | -21 | 15 |
| 2020-06-27 | -46 | -7 | -47 | -24 | -14 | 13 |
| 2020-06-28 | -48 | -14 | -43 | -32 | -9 | 12 |
| 2020-06-29 | -42 | -11 | -45 | -32 | -22 | 15 |
| 2020-06-30 | -42 | -5 | -51 | -29 | -26 | 15 |
| 2020-07-01 | -35 | 6 | -42 | -22 | -22 | 14 |
| 2020-07-02 | -37 | 3 | -38 | -20 | -22 | 14 |
| 2020-07-03 | -33 | 8 | -41 | -11 | -18 | 14 |
| 2020-07-04 | -37 | 5 | -14 | -10 | -10 | 11 |
| 2020-07-05 | -38 | -5 | -27 | -27 | -5 | 10 |

| | | | | | | |
|---|---|---|---|---|---|---|
| 2020-07-06 | -26 | 2 | -36 | -12 | -18 | 12 |
| 2020-07-07 | -27 | 7 | -34 | -12 | -21 | 12 |
| 2020-07-08 | -23 | 8 | -32 | -11 | -18 | 12 |
| 2020-07-09 | -26 | 8 | -32 | -13 | -18 | 12 |
| 2020-07-10 | -27 | 7 | -30 | -1 | -16 | 13 |
| 2020-07-11 | -27 | 7 | -24 | 4 | -5 | 10 |
| 2020-07-12 | -32 | -2 | -5 | -23 | -5 | 9 |
| 2020-07-13 | -28 | -2 | -33 | -18 | -16 | 11 |
| 2020-07-14 | -29 | 0 | -41 | -18 | -21 | 12 |
| 2020-07-15 | -25 | 9 | -41 | -11 | -17 | 13 |
| 2020-07-16 | -27 | 4 | -35 | -15 | -17 | 13 |
| 2020-07-17 | -24 | 11 | -22 | -6 | -13 | 13 |
| 2020-07-18 | -26 | 7 | -29 | 0 | -7 | 11 |
| 2020-07-19 | -34 | -7 | -1 | -18 | -7 | 10 |
| 2020-07-20 | -37 | -6 | -38 | -25 | -17 | 12 |
| 2020-07-21 | -39 | -5 | -42 | -21 | -19 | 13 |
| 2020-07-22 | -34 | 0 | -41 | -15 | -19 | 14 |
| 2020-07-23 | -36 | 0 | -34 | -16 | -18 | 13 |
| 2020-07-24 | -34 | 7 | -11 | -10 | -14 | 13 |
| 2020-07-25 | -38 | 3 | -29 | -5 | -10 | 11 |
| 2020-07-26 | -33 | 0 | -1 | -16 | -7 | 9 |
| 2020-07-27 | -31 | -4 | -39 | -15 | -15 | 12 |
| 2020-07-28 | -29 | -1 | -29 | -9 | -17 | 12 |
| 2020-07-29 | -25 | 6 | -33 | -6 | -17 | 12 |
| 2020-07-30 | -29 | 0 | -35 | -10 | -17 | 13 |
| 2020-07-31 | -24 | 10 | -5 | -3 | -13 | 12 |
| 2020-08-01 | -28 | 7 | -19 | 11 | -8 | 11 |
| 2020-08-02 | -28 | 1 | 7 | -5 | -6 | 10 |
| 2020-08-03 | -26 | 1 | -37 | -9 | -15 | 11 |
| 2020-08-04 | -25 | 2 | -31 | -2 | -17 | 12 |
| 2020-08-05 | -20 | 19 | -31 | -4 | -14 | 11 |
| 2020-08-06 | -19 | 16 | -27 | 7 | -14 | 11 |
| 2020-08-07 | -17 | 20 | -18 | 12 | -10 | 10 |
| 2020-08-08 | -16 | 22 | -8 | 24 | -3 | 8 |
| 2020-08-09 | -31 | 10 | -6 | -4 | 5 | 6 |
| 2020-08-10 | -24 | 3 | -33 | -3 | -13 | 10 |
| 2020-08-11 | -27 | 4 | -29 | -5 | -16 | 11 |
| 2020-08-12 | -23 | 11 | -29 | -4 | -14 | 11 |
| 2020-08-13 | -24 | 10 | -26 | 0 | -14 | 11 |
| 2020-08-14 | -20 | 16 | -7 | 9 | -10 | 11 |
| 2020-08-15 | -22 | 12 | 13 | 12 | -7 | 8 |
| 2020-08-16 | -24 | 10 | 38 | 3 | -7 | 8 |

| | | | | | | |
|---|---|---|---|---|---|---|
| 2020-08-17 | -45 | -46 | | | -30 | 14 |
| 2020-08-18 | -21 | 17 | | | -12 | 10 |
| 2020-08-19 | -21 | 13 | | | -11 | 10 |
| 2020-08-20 | -28 | 8 | | | -12 | 11 |
| 2020-08-21 | -33 | 11 | | | -10 | 12 |
| 2020-08-22 | -41 | 2 | | | -9 | 13 |
| 2020-08-23 | -38 | | | | -8 | 11 |
| 2020-08-24 | -32 | 3 | | | -10 | 10 |
| 2020-08-25 | -32 | 3 | | | -12 | 11 |
| 2020-08-26 | -27 | 14 | | | -9 | 11 |
| 2020-08-27 | -25 | 14 | | | -9 | 10 |
| 2020-08-28 | -22 | 18 | | | -5 | 10 |
| 2020-08-29 | -19 | 16 | | | -2 | 7 |
| 2020-08-30 | -21 | | | | -2 | 7 |
| 2020-08-31 | -27 | 8 | | | -9 | 8 |
| 2020-09-01 | -27 | 12 | | | -9 | 9 |
| 2020-09-02 | -20 | 20 | | | -7 | 8 |
| 2020-09-03 | -20 | 15 | | | -6 | 8 |
| 2020-09-04 | -14 | 22 | | | -4 | 7 |
| 2020-09-05 | -12 | 24 | | | -2 | 4 |
| 2020-09-06 | -6 | | | | -4 | 4 |
| 2020-09-07 | -41 | -17 | | | -58 | 11 |
| 2020-09-08 | -19 | 14 | | | -7 | 6 |
| 2020-09-09 | -17 | 20 | | | -4 | 8 |
| 2020-09-10 | -19 | 14 | | | -5 | 8 |
| 2020-09-11 | -18 | 12 | -4 | 31 | -1 | 8 |
| 2020-09-12 | -16 | 19 | 16 | 48 | 4 | 5 |
| 2020-09-13 | -18 | 22 | 51 | 33 | -2 | 5 |
| 2020-09-14 | -22 | 5 | -29 | 11 | -6 | 7 |
| 2020-09-15 | -23 | 7 | -36 | 15 | -4 | 9 |
| 2020-09-16 | -21 | 16 | -26 | 12 | -4 | 9 |
| 2020-09-17 | -23 | 9 | -20 | 17 | -4 | 9 |
| 2020-09-18 | -20 | 14 | 2 | 27 | -1 | 8 |
| 2020-09-19 | -18 | 16 | 19 | 47 | 2 | 6 |
| 2020-09-20 | -28 | 12 | 13 | 20 | -3 | 7 |
| 2020-09-21 | -29 | 0 | -37 | 16 | -9 | 9 |
| 2020-09-22 | -38 | -8 | -48 | -4 | -9 | 11 |
| 2020-09-23 | -25 | 15 | -32 | 2 | -5 | 10 |
| 2020-09-24 | -25 | 9 | -26 | 5 | -5 | 10 |
| 2020-09-25 | -19 | 16 | -4 | 30 | 0 | 9 |
| 2020-09-26 | -20 | 18 | 14 | 37 | 3 | 7 |
| 2020-09-27 | -21 | 19 | 38 | 16 | -2 | 6 |

| | | | | | | |
|---|---|---|---|---|---|---|
| 2020-09-28 | -28 | 1 | -32 | 4 | -7 | 9 |
| 2020-09-29 | -29 | 6 | -26 | 17 | -5 | 8 |
| 2020-09-30 | -19 | 23 | -23 | 18 | -3 | 8 |
| 2020-10-01 | -20 | 20 | -16 | 27 | -3 | 6 |
| 2020-10-02 | -20 | 18 | -5 | 49 | -2 | 1 |
| 2020-10-03 | -13 | 28 | 4 | 77 | 2 | 1 |
| 2020-10-04 | -21 | 32 | 8 | 33 | -3 | 5 |
| 2020-10-05 | -21 | 16 | -30 | 38 | -7 | 6 |
| 2020-10-06 | -17 | 19 | -19 | 43 | -5 | 6 |
| 2020-10-07 | -12 | 30 | -17 | 40 | -2 | 6 |
| 2020-10-08 | -16 | 19 | -25 | 33 | -3 | 7 |
| 2020-10-09 | -13 | 22 | 31 | 55 | 1 | 7 |
| 2020-10-10 | -6 | 31 | 5 | 76 | 2 | 4 |
| 2020-10-11 | -8 | 34 | 62 | 28 | 0 | 7 |
| 2020-10-12 | -35 | -12 | 31 | 8 | -59 | 13 |
| 2020-10-13 | -17 | 14 | -27 | 32 | -3 | 6 |
| 2020-10-14 | -14 | 21 | -27 | 22 | -2 | 8 |
| 2020-10-15 | -14 | 19 | -14 | 34 | -3 | 8 |
| 2020-10-16 | -13 | 19 | -7 | 37 | 2 | 8 |
| 2020-10-17 | -10 | 21 | 5 | 65 | 5 | 7 |
| 2020-10-18 | -17 | 26 | 33 | 22 | -2 | 8 |
| 2020-10-19 | -19 | 9 | -21 | 32 | -6 | 8 |
| 2020-10-20 | -19 | 14 | -19 | 33 | -2 | 8 |
| 2020-10-21 | -16 | 21 | -25 | 27 | 0 | 9 |
| 2020-10-22 | -14 | 24 | -11 | 33 | 0 | 8 |
| 2020-10-23 | -11 | 26 | -2 | 49 | 4 | 7 |
| 2020-10-24 | -24 | 9 | -26 | 43 | 0 | 9 |
| 2020-10-25 | -28 | 18 | -16 | 24 | -4 | 9 |
| 2020-10-26 | -25 | 8 | -32 | 21 | -4 | 8 |
| 2020-10-27 | -17 | 20 | -20 | 25 | 0 | 8 |
| 2020-10-28 | -15 | 20 | -17 | 21 | 1 | 7 |
| 2020-10-29 | -16 | 17 | -23 | 34 | -1 | 7 |
| 2020-10-30 | -12 | 26 | -13 | 47 | 0 | 7 |
| 2020-10-31 | -15 | 26 | -14 | 69 | 2 | 6 |
| 2020-11-01 | -16 | 34 | 13 | 16 | -2 | 8 |
| 2020-11-02 | -47 | -15 | -6 | -8 | -58 | 15 |
| 2020-11-03 | -17 | 21 | -35 | 38 | 0 | 6 |
| 2020-11-04 | -16 | 26 | -37 | 23 | 0 | 8 |
| 2020-11-05 | -13 | 26 | 13 | 33 | 1 | 7 |
| 2020-11-06 | -9 | 29 | -13 | 44 | 4 | 6 |
| 2020-11-07 | -2 | 37 | 6 | 76 | 9 | 4 |
| 2020-11-08 | -8 | 38 | 24 | 36 | 2 | 6 |

| | | | | | | |
|---|---|---|---|---|---|---|
| 2020-11-09 | -17 | 16 | -25 | 35 | -2 | 6 |
| 2020-11-10 | -17 | 16 | -32 | 32 | 0 | 7 |
| 2020-11-11 | -10 | 29 | -27 | 27 | 1 | 7 |
| 2020-11-12 | -16 | 16 | -28 | 27 | 1 | 8 |
| 2020-11-13 | -8 | 28 | -6 | 44 | 5 | 6 |
| 2020-11-14 | -8 | 25 | -9 | 81 | 8 | 5 |
| 2020-11-15 | 20 | 79 | 33 | 63 | 12 | 1 |
| 2020-11-16 | -26 | -2 | -40 | 20 | -3 | 8 |
| 2020-11-17 | -16 | 20 | -26 | 35 | 1 | 6 |
| 2020-11-18 | -18 | 19 | -34 | 28 | 3 | 8 |
| 2020-11-19 | -11 | 30 | -26 | 43 | 1 | 6 |
| 2020-11-20 | -29 | 12 | -3 | 23 | -35 | 14 |
| 2020-11-21 | -12 | 22 | 0 | 49 | 2 | 6 |
| 2020-11-22 | -17 | 26 | -1 | 50 | -3 | 6 |
| 2020-11-23 | -19 | 17 | -32 | 18 | -2 | 7 |
| 2020-11-24 | -19 | 16 | -30 | 22 | 1 | 7 |
| 2020-11-25 | -14 | 38 | -25 | 17 | 4 | 7 |
| 2020-11-26 | -13 | 41 | -12 | 35 | 4 | 6 |
| 2020-11-27 | -4 | 51 | 9 | 43 | 6 | 5 |
| 2020-11-28 | -7 | 32 | 8 | 68 | 13 | 4 |
| 2020-11-29 | -11 | 40 | 20 | 41 | 3 | 5 |
| 2020-11-30 | -17 | 20 | -27 | 24 | 0 | 6 |
| 2020-12-01 | -24 | 10 | -33 | 24 | 1 | 8 |
| 2020-12-02 | -13 | 28 | -32 | 19 | 3 | 7 |
| 2020-12-03 | -12 | 29 | -21 | 24 | 3 | 6 |
| 2020-12-04 | -9 | 30 | -9 | 42 | 7 | 6 |
| 2020-12-05 | -13 | 24 | -14 | 68 | 9 | 5 |
| 2020-12-06 | -15 | 42 | 1 | 27 | 2 | 6 |
| 2020-12-07 | -21 | 13 | -38 | 17 | -1 | 7 |
| 2020-12-08 | -32 | 1 | -48 | 1 | -9 | 12 |
| 2020-12-09 | -11 | 32 | -30 | 16 | 5 | 7 |
| 2020-12-10 | -15 | 18 | 3 | 22 | 2 | 7 |
| 2020-12-11 | -9 | 25 | -6 | 25 | 7 | 6 |
| 2020-12-12 | -13 | 24 | -14 | 42 | 11 | 6 |
| 2020-12-13 | -5 | 46 | 18 | 31 | 9 | 7 |
| 2020-12-14 | -12 | 18 | -22 | 21 | 0 | 5 |
| 2020-12-15 | -9 | 23 | -19 | 22 | 3 | 5 |
| 2020-12-16 | -7 | 28 | -23 | 17 | 4 | 7 |
| 2020-12-17 | -8 | 29 | -16 | 21 | 3 | 6 |
| 2020-12-18 | -9 | 27 | -10 | 25 | 3 | 7 |
| 2020-12-19 | -5 | 35 | 11 | 59 | 13 | 4 |
| 2020-12-20 | 10 | 81 | 27 | 42 | 26 | 4 |

| | | | | | | |
|---|---|---|---|---|---|---|
| 2020-12-21 | 0 | 39 | -5 | 19 | -8 | 5 |
| 2020-12-22 | -8 | 38 | -11 | 31 | -10 | 7 |
| 2020-12-23 | 13 | 79 | -25 | 56 | -12 | 6 |
| 2020-12-24 | -12 | 68 | -35 | 25 | -41 | 11 |
| 2020-12-25 | -75 | -74 | -63 | -49 | -75 | 20 |
| 2020-12-26 | -31 | 1 | -25 | 27 | -20 | 11 |
| 2020-12-27 | -25 | 8 | -4 | 55 | -5 | 9 |
| 2020-12-28 | -17 | 18 | -16 | 17 | -31 | 12 |
| 2020-12-29 | -16 | 25 | -15 | 20 | -30 | 11 |
| 2020-12-30 | -2 | 65 | 15 | 52 | -29 | 9 |
| 2020-12-31 | -36 | 39 | 5 | 8 | -53 | 12 |
| 2021-01-01 | -75 | -78 | -22 | -47 | -77 | 20 |
| 2021-01-02 | -49 | -21 | -16 | 15 | -38 | 13 |
| 2021-01-03 | -33 | 6 | -6 | 47 | -12 | 8 |
| 2021-01-04 | -18 | 17 | -17 | 23 | -16 | 9 |
| 2021-01-05 | -21 | 14 | -21 | 9 | -12 | 9 |
| 2021-01-06 | -21 | 15 | -29 | 8 | -10 | 10 |
| 2021-01-07 | -18 | 19 | -14 | 17 | -8 | 9 |
| 2021-01-08 | -18 | 15 | -10 | 20 | -5 | 9 |
| 2021-01-09 | -22 | 12 | -17 | 25 | -3 | 8 |
| 2021-01-10 | -22 | 25 | 14 | 25 | -2 | 7 |
| 2021-01-11 | -18 | 14 | -12 | 13 | -4 | 7 |
| 2021-01-12 | -20 | 12 | -11 | 12 | -4 | 6 |
| 2021-01-13 | -18 | 22 | -12 | -1 | -4 | 5 |
| 2021-01-14 | -23 | 8 | -14 | 4 | -8 | 5 |
| 2021-01-15 | -22 | 10 | 1 | 14 | -6 | 5 |
| 2021-01-16 | -26 | 4 | 2 | 17 | -7 | 5 |
| 2021-01-17 | -22 | 19 | 41 | 2 | -7 | 3 |
| 2021-01-18 | -29 | 2 | -10 | -1 | -11 | 5 |
| 2021-01-19 | -28 | 6 | -17 | -3 | -9 | 5 |
| 2021-01-20 | -27 | 18 | -15 | 4 | -6 | 6 |
| 2021-01-21 | -32 | 8 | -8 | 1 | -8 | 6 |
| 2021-01-22 | -31 | 13 | -6 | 19 | -4 | 6 |
| 2021-01-23 | -51 | -1 | -16 | 4 | -17 | 8 |
| 2021-01-24 | -45 | 13 | 9 | 0 | -14 | 5 |
| 2021-01-25 | -33 | 3 | -15 | 6 | -7 | 5 |
| 2021-01-26 | -33 | 1 | -17 | 6 | -6 | 6 |
| 2021-01-27 | -29 | 11 | -14 | 0 | -4 | 6 |
| 2021-01-28 | -30 | 6 | -8 | 2 | -6 | 6 |
| 2021-01-29 | -27 | 10 | 4 | 29 | -2 | 5 |
| 2021-01-30 | -47 | -3 | -12 | 8 | -15 | 6 |
| 2021-01-31 | -43 | 15 | 20 | 6 | -14 | 4 |

| | | | | | | |
|---|---|---|---|---|---|---|
| 2021-02-01 | -25 | 2 | -11 | 7 | -6 | 4 |
| 2021-02-02 | -25 | 1 | -31 | 3 | -5 | 5 |
| 2021-02-03 | -18 | 13 | -20 | 7 | -3 | 5 |
| 2021-02-04 | -21 | 8 | -11 | 15 | -3 | 5 |
| 2021-02-05 | -19 | 11 | -11 | 17 | -1 | 5 |
| 2021-02-06 | -37 | -6 | -53 | 15 | -9 | 9 |
| 2021-02-07 | -37 | 12 | -30 | 2 | -13 | 8 |
| 2021-02-08 | -25 | 0 | -31 | 8 | -7 | 5 |
| 2021-02-09 | -25 | 1 | -33 | 9 | -4 | 6 |
| 2021-02-10 | -12 | 18 | -21 | 18 | -1 | 5 |
| 2021-02-11 | -14 | 8 | -9 | 42 | -3 | 5 |
| 2021-02-12 | -14 | 15 | -12 | 27 | -1 | 5 |
| 2021-02-13 | -18 | 15 | -15 | 37 | -4 | 4 |
| 2021-02-14 | -18 | 19 | 23 | 17 | -5 | 6 |
| 2021-02-15 | -22 | 5 | 17 | 8 | -38 | 10 |
| 2021-02-16 | -44 | -25 | 28 | -8 | -54 | 14 |
| 2021-02-17 | -14 | 13 | -13 | 20 | -9 | 6 |
| 2021-02-18 | -20 | 7 | -25 | 3 | -5 | 5 |
| 2021-02-19 | -17 | 9 | -8 | 21 | -1 | 6 |
| 2021-02-20 | -22 | 6 | -23 | 26 | 1 | 6 |
| 2021-02-21 | -24 | 18 | -10 | 22 | -5 | 6 |
| 2021-02-22 | -22 | 3 | -30 | 13 | -3 | 5 |
| 2021-02-23 | -20 | 7 | -27 | 10 | -1 | 5 |
| 2021-02-24 | -14 | 15 | -18 | 10 | 1 | 5 |
| 2021-02-25 | -18 | 13 | -12 | 6 | 0 | 5 |
| 2021-02-26 | -16 | 13 | -1 | 25 | 4 | 5 |
| 2021-02-27 | -18 | 16 | -10 | 37 | 1 | 5 |
| 2021-02-28 | -23 | 22 | -8 | 22 | -3 | 6 |
| 2021-03-01 | -21 | 10 | -16 | 21 | -2 | 5 |
| 2021-03-02 | -21 | 7 | -13 | 21 | -1 | 5 |
| 2021-03-03 | -15 | 19 | -19 | 20 | 0 | 6 |
| 2021-03-04 | -17 | 15 | -9 | 12 | 0 | 6 |
| 2021-03-05 | -14 | 20 | -1 | 31 | 3 | 5 |
| 2021-03-06 | -18 | 18 | -23 | 43 | 1 | 4 |
| 2021-03-07 | -31 | 18 | -29 | 23 | -9 | 7 |
| 2021-03-08 | -18 | 8 | -23 | 12 | -2 | 4 |
| 2021-03-09 | -21 | 8 | -28 | 10 | -1 | 6 |
| 2021-03-10 | -13 | 23 | -20 | 12 | 2 | 5 |
| 2021-03-11 | -17 | 13 | -18 | 10 | 1 | 6 |
| 2021-03-12 | -16 | 12 | -3 | 23 | 5 | 5 |
| 2021-03-13 | -18 | 15 | -7 | 34 | 1 | 5 |
| 2021-03-14 | -15 | 25 | 20 | 33 | 0 | 5 |

| | | | | | | |
|---|---|---|---|---|---|---|
| 2021-03-15 | -20 | 14 | -16 | 12 | -2 | 5 |
| 2021-03-16 | -28 | 7 | -26 | 6 | -1 | 6 |
| 2021-03-17 | -19 | 18 | -12 | 8 | 2 | 6 |
| 2021-03-18 | -21 | 9 | -12 | 9 | 1 | 6 |
| 2021-03-19 | -21 | 15 | -7 | 17 | 4 | 6 |
| 2021-03-20 | -24 | 14 | -12 | 38 | 1 | 6 |
| 2021-03-21 | -24 | 20 | 7 | 19 | -4 | 6 |
| 2021-03-22 | -24 | 6 | -14 | 8 | -1 | 5 |
| 2021-03-23 | -25 | 4 | -21 | 7 | -1 | 7 |
| 2021-03-24 | -20 | 17 | -13 | 3 | 1 | 7 |
| 2021-03-25 | -22 | 14 | -15 | 13 | -1 | 7 |
| 2021-03-26 | -24 | 19 | -5 | 7 | -6 | 9 |
| 2021-03-27 | -35 | 5 | -26 | 7 | -3 | 8 |
| 2021-03-28 | -36 | 19 | -22 | -22 | -7 | 9 |
| 2021-03-29 | -48 | -3 | -27 | -28 | -32 | 14 |
| 2021-03-30 | -51 | -2 | -39 | -26 | -32 | 15 |
| 2021-03-31 | -40 | 23 | -28 | -20 | -29 | 14 |
| 2021-04-01 | -35 | 35 | -16 | -5 | -31 | 12 |
| 2021-04-02 | -64 | -18 | -36 | -41 | -65 | 23 |
| 2021-04-03 | -52 | 5 | -43 | -22 | -30 | 13 |
| 2021-04-04 | -53 | 12 | -43 | -14 | -13 | 9 |
| 2021-04-05 | -39 | 3 | -31 | -4 | -10 | 9 |
| 2021-04-06 | -39 | 6 | -27 | -4 | -10 | 10 |
| 2021-04-07 | -33 | 18 | -30 | 1 | -9 | 11 |
| 2021-04-08 | -35 | 14 | -29 | 8 | -8 | 10 |
| 2021-04-09 | -35 | 24 | -19 | 11 | -5 | 10 |
| 2021-04-10 | -42 | 4 | -35 | 1 | -11 | 10 |
| 2021-04-11 | -44 | 39 | -27 | -12 | -11 | 9 |
| 2021-04-12 | -36 | 5 | -19 | -1 | -12 | 9 |
| 2021-04-13 | -41 | 16 | -37 | -12 | -15 | 12 |
| 2021-04-14 | -32 | 13 | -23 | 3 | -12 | 11 |
| 2021-04-15 | -34 | 31 | -25 | 3 | -12 | 11 |
| 2021-04-16 | -34 | 14 | -16 | 11 | -9 | 12 |
| 2021-04-17 | -42 | 16 | -30 | 20 | -4 | 9 |
| 2021-04-18 | -41 | 23 | -22 | 5 | -4 | 10 |
| 2021-04-19 | -39 | 15 | -29 | -4 | -12 | 10 |
| 2021-04-20 | -35 | 10 | -27 | -3 | -12 | 10 |
| 2021-04-21 | -41 | 17 | -34 | -22 | -26 | 15 |
| 2021-04-22 | -32 | 13 | -18 | 2 | -13 | 11 |
| 2021-04-23 | -38 | 19 | -10 | -1 | -22 | 14 |
| 2021-04-24 | -39 | 5 | -24 | 8 | -6 | 9 |
| 2021-04-25 | -40 | 30 | -6 | 14 | -5 | 9 |

| | | | | | | |
|---|---|---|---|---|---|---|
| 2021-04-26 | -29 | 14 | -22 | 3 | -2 | 7 |
| 2021-04-27 | -32 | 14 | -31 | -2 | -2 | 9 |
| 2021-04-28 | -23 | 25 | -33 | -2 | -1 | 8 |
| 2021-04-29 | -31 | 14 | -33 | -3 | -4 | 10 |
| 2021-04-30 | -19 | 32 | -9 | 26 | 5 | 7 |
| 2021-05-01 | -35 | 14 | -29 | 21 | -20 | 11 |
| 2021-05-02 | -25 | 31 | -8 | 18 | -2 | 9 |
| 2021-05-03 | -21 | 24 | -18 | 8 | 0 | 6 |
| 2021-05-04 | -22 | 22 | -23 | 8 | 1 | 7 |
| 2021-05-05 | -19 | 29 | -21 | 12 | 2 | 7 |
| 2021-05-06 | -18 | 30 | -17 | 10 | 2 | 7 |
| 2021-05-07 | -12 | 42 | 6 | 34 | 6 | 6 |
| 2021-05-08 | -14 | 43 | -26 | 45 | 3 | 6 |
| 2021-05-09 | -24 | 47 | -9 | 39 | 14 | 4 |
| 2021-05-10 | -21 | 21 | -21 | 15 | -2 | 6 |
| 2021-05-11 | -21 | 24 | -25 | 10 | 2 | 7 |
| 2021-05-12 | -18 | 31 | -15 | 4 | 3 | 7 |
| 2021-05-13 | -24 | 23 | -27 | 8 | 1 | 8 |
| 2021-05-14 | -16 | 33 | -2 | 20 | 4 | 7 |
| 2021-05-15 | -19 | 27 | 5 | 34 | 2 | 7 |
| 2021-05-16 | -22 | 29 | 12 | 17 | -5 | 9 |
| 2021-05-17 | -20 | 24 | -12 | 9 | 2 | 6 |
| 2021-05-18 | -22 | 27 | -18 | 4 | 2 | 7 |
| 2021-05-19 | -19 | 31 | -22 | 4 | 3 | 8 |
| 2021-05-20 | -18 | 31 | -6 | 10 | 4 | 7 |
| 2021-05-21 | -15 | 34 | 13 | 30 | 7 | 7 |
| 2021-05-22 | -18 | 30 | 16 | 36 | 3 | 7 |
| 2021-05-23 | -19 | 30 | 35 | 29 | -1 | 7 |
| 2021-05-24 | -24 | 22 | -6 | 7 | 2 | 7 |
| 2021-05-25 | -22 | 25 | -11 | 15 | 4 | 7 |
| 2021-05-26 | -19 | 31 | -14 | 6 | 4 | 7 |
| 2021-05-27 | -20 | 27 | -8 | 17 | 4 | 7 |
| 2021-05-28 | -14 | 36 | 17 | 29 | 8 | 7 |
| 2021-05-29 | -15 | 32 | 14 | 42 | 4 | 6 |
| 2021-05-30 | -21 | 37 | 15 | 22 | -8 | 7 |
| 2021-05-31 | -30 | 12 | -30 | 0 | 1 | 8 |
| 2021-06-01 | -18 | 40 | -16 | 13 | 3 | 6 |
| 2021-06-02 | -6 | 54 | -7 | 30 | 6 | 5 |
| 2021-06-03 | -31 | 17 | 29 | -1 | -43 | 15 |
| 2021-06-04 | -2 | 56 | 58 | 44 | -12 | 8 |
| 2021-06-05 | -10 | 42 | 36 | 63 | 1 | 6 |
| 2021-06-06 | -13 | 46 | 41 | 48 | -6 | 6 |

| | | | | | | |
|---|---|---|---|---|---|---|
| 2021-06-07 | -14 | 36 | -3 | 22 | 4 | 5 |
| 2021-06-08 | -15 | 37 | -14 | 19 | 4 | 6 |
| 2021-06-09 | -10 | 47 | -12 | 16 | 7 | 6 |
| 2021-06-10 | -13 | 41 | -6 | 22 | 6 | 6 |
| 2021-06-11 | -6 | 46 | 25 | 39 | 9 | 6 |
| 2021-06-12 | 1 | 44 | 43 | 65 | 5 | 5 |
| 2021-06-13 | -14 | 35 | 37 | 41 | -9 | 7 |
| 2021-06-14 | -17 | 27 | 1 | 14 | 2 | 6 |
| 2021-06-15 | -19 | 32 | -6 | 8 | 4 | 6 |
| 2021-06-16 | -13 | 41 | -5 | -2 | 6 | 6 |
| 2021-06-17 | -17 | 32 | -5 | 4 | 5 | 6 |
| 2021-06-18 | -12 | 36 | 19 | 12 | 8 | 6 |
| 2021-06-19 | -16 | 30 | 14 | 39 | 1 | 6 |
| 2021-06-20 | -14 | 37 | 44 | 30 | -6 | 7 |
| 2021-06-21 | -19 | 28 | -5 | 5 | 3 | 6 |
| 2021-06-22 | -22 | 24 | -16 | 1 | 4 | 7 |
| 2021-06-23 | -15 | 35 | -11 | 6 | 7 | 7 |
| 2021-06-24 | -13 | 31 | 3 | 11 | 6 | 6 |
| 2021-06-25 | -7 | 36 | 31 | 24 | 10 | 6 |
| 2021-06-26 | -10 | 30 | 35 | 48 | 2 | 5 |
| 2021-06-27 | -6 | 36 | 72 | 35 | -8 | 6 |
| 2021-06-28 | -17 | 23 | 0 | 16 | 5 | 5 |
| 2021-06-29 | -20 | 21 | -12 | 5 | 4 | 5 |
| 2021-06-30 | -15 | 34 | -23 | 1 | 6 | 7 |
| 2021-07-01 | -10 | 41 | 3 | 12 | 7 | 6 |
| 2021-07-02 | -4 | 45 | 30 | 32 | 10 | 6 |
| 2021-07-03 | -7 | 44 | 34 | 58 | 5 | 5 |
| 2021-07-04 | -6 | 45 | 70 | 46 | -5 | 6 |
| 2021-07-05 | -11 | 34 | -9 | 20 | 7 | 5 |
| 2021-07-06 | -12 | 39 | -1 | 19 | 8 | 6 |
| 2021-07-07 | -2 | 56 | 2 | 19 | 10 | 5 |
| 2021-07-08 | -4 | 47 | 9 | 22 | 9 | 5 |
| 2021-07-09 | 1 | 49 | 31 | 41 | 11 | 5 |
| 2021-07-10 | -1 | 48 | 48 | 61 | 7 | 4 |
| 2021-07-11 | -2 | 53 | 74 | 53 | -4 | 5 |
| 2021-07-12 | -9 | 39 | 1 | 24 | 6 | 5 |
| 2021-07-13 | -11 | 37 | 0 | 20 | 7 | 5 |
| 2021-07-14 | -5 | 48 | 5 | 14 | 9 | 6 |
| 2021-07-15 | -6 | 43 | 13 | 22 | 8 | 5 |
| 2021-07-16 | -4 | 43 | 45 | 41 | 11 | 5 |
| 2021-07-17 | -6 | 41 | 57 | 58 | 4 | 4 |
| 2021-07-18 | -2 | 46 | 93 | 57 | -4 | 4 |

| | | | | | | |
|---|---|---|---|---|---|---|
| 2021-07-19 | -12 | 31 | 6 | 24 | 3 | 5 |
| 2021-07-20 | -11 | 37 | 9 | 26 | 3 | 5 |
| 2021-07-21 | -7 | 47 | 9 | 16 | 5 | 6 |
| 2021-07-22 | -6 | 44 | 31 | 28 | 4 | 5 |
| 2021-07-23 | -5 | 43 | 51 | 39 | 8 | 5 |
| 2021-07-24 | -9 | 39 | 61 | 67 | 3 | 4 |
| 2021-07-25 | -4 | 43 | 106 | 55 | -7 | 5 |
| 2021-07-26 | -13 | 35 | 15 | 23 | 4 | 5 |
| 2021-07-27 | -13 | 35 | 11 | 11 | 4 | 6 |
| 2021-07-28 | -10 | 42 | 8 | 7 | 7 | 6 |
| 2021-07-29 | -13 | 37 | 14 | 15 | 6 | 6 |
| 2021-07-30 | -6 | 44 | 50 | 43 | 11 | 6 |
| 2021-07-31 | -10 | 40 | 47 | 50 | 5 | 6 |
| 2021-08-01 | -15 | 36 | 51 | 46 | -4 | 7 |
| 2021-08-02 | -13 | 40 | 4 | 20 | 10 | 5 |
| 2021-08-03 | -13 | 35 | -6 | 19 | 11 | 6 |
| 2021-08-04 | -5 | 54 | -1 | 14 | 13 | 6 |
| 2021-08-05 | -8 | 46 | 6 | 25 | 12 | 5 |
| 2021-08-06 | 4 | 55 | 36 | 62 | 15 | 4 |
| 2021-08-07 | -1 | 52 | 31 | 78 | 7 | 4 |
| 2021-08-08 | -9 | 50 | 32 | 73 | 2 | 3 |
| 2021-08-09 | -10 | 36 | 4 | 37 | 10 | 4 |
| 2021-08-10 | -6 | 42 | 6 | 29 | 11 | 4 |
| 2021-08-11 | -3 | 52 | 13 | 22 | 13 | 5 |
| 2021-08-12 | -9 | 41 | 8 | 20 | 11 | 5 |
| 2021-08-13 | -5 | 43 | 29 | 41 | 14 | 5 |
| 2021-08-14 | -8 | 39 | 26 | 48 | 5 | 5 |
| 2021-08-15 | -6 | 46 | 62 | 50 | -4 | 6 |
| 2021-08-16 | -24 | 15 | -2 | 23 | -7 | 8 |
| 2021-08-17 | -10 | 40 | -2 | 21 | 10 | 5 |
| 2021-08-18 | -8 | 47 | 1 | 20 | 14 | 6 |
| 2021-08-19 | -8 | 41 | 13 | 23 | 12 | 5 |
| 2021-08-20 | -3 | 46 | 40 | 44 | 15 | 5 |
| 2021-08-21 | -6 | 41 | 48 | 63 | 6 | 4 |
| 2021-08-22 | -6 | 52 | 84 | 58 | -4 | 5 |
| 2021-08-23 | -15 | 34 | 3 | 27 | 11 | 4 |
| 2021-08-24 | -15 | 35 | 4 | 21 | 12 | 5 |
| 2021-08-25 | -5 | 50 | 10 | 19 | 13 | 5 |
| 2021-08-26 | -11 | 40 | 9 | 27 | 13 | 5 |
| 2021-08-27 | -10 | 40 | 31 | 42 | 16 | 5 |
| 2021-08-28 | -14 | 35 | 27 | 52 | 7 | 5 |
| 2021-08-29 | -14 | 46 | 36 | 46 | -4 | 6 |

| | | | | | | |
|---|---|---|---|---|---|---|
| 2021-08-30 | -21 | 29 | -11 | 20 | 11 | 5 |
| 2021-08-31 | -19 | 35 | -20 | 12 | 13 | 5 |
| 2021-09-01 | -7 | 56 | -3 | 16 | 13 | 5 |
| 2021-09-02 | -8 | 52 | 17 | 18 | 14 | 4 |
| 2021-09-03 | -3 | 56 | 43 | 65 | 18 | 3 |
| 2021-09-04 | -9 | 49 | 44 | 65 | 5 | 2 |
| 2021-09-05 | 2 | 65 | 103 | 58 | -3 | 3 |
| 2021-09-06 | -1 | 58 | 64 | 47 | -15 | 5 |
| 2021-09-07 | -36 | -1 | 62 | 21 | -48 | 13 |
| 2021-09-08 | -5 | 61 | 2 | 31 | 17 | 4 |
| 2021-09-09 | -5 | 54 | 16 | 38 | 16 | 3 |
| 2021-09-10 | -2 | 54 | 29 | 44 | 17 | 4 |
| 2021-09-11 | -9 | 44 | 24 | 46 | 8 | 5 |
| 2021-09-12 | -5 | 56 | 55 | 61 | 0 | 4 |
| 2021-09-13 | -9 | 42 | 5 | 44 | 12 | 3 |
| 2021-09-14 | -9 | 44 | 6 | 26 | 13 | 3 |
| 2021-09-15 | -4 | 55 | 10 | 27 | 16 | 4 |
| 2021-09-16 | -8 | 46 | 9 | 22 | 14 | 4 |
| 2021-09-17 | -2 | 49 | 39 | 45 | 17 | 4 |
| 2021-09-18 | -7 | 41 | 44 | 66 | 7 | 2 |
| 2021-09-19 | -4 | 51 | 70 | 53 | -5 | 3 |
| 2021-09-20 | -10 | 38 | 13 | 32 | 12 | 3 |
| 2021-09-21 | -13 | 37 | 0 | 42 | 14 | 3 |
| 2021-09-22 | -9 | 47 | 4 | 6 | 17 | 4 |
| 2021-09-23 | -8 | 46 | 12 | 28 | 17 | 4 |
| 2021-09-24 | -2 | 48 | 38 | 44 | 18 | 4 |
| 2021-09-25 | -10 | 41 | 32 | 48 | 9 | 3 |
| 2021-09-26 | -9 | 53 | 58 | 51 | -1 | 3 |
| 2021-09-27 | -17 | 34 | 0 | 24 | 14 | 3 |
| 2021-09-28 | -14 | 40 | 0 | 8 | 16 | 3 |
| 2021-09-29 | -8 | 51 | 7 | 5 | 19 | 4 |
| 2021-09-30 | -7 | 50 | 12 | 9 | 17 | 4 |
| 2021-10-01 | -1 | 56 | 33 | 38 | 20 | 4 |
| 2021-10-02 | -6 | 45 | 18 | 43 | 12 | 3 |
| 2021-10-03 | -6 | 62 | 51 | 34 | -1 | 4 |
| 2021-10-04 | -14 | 35 | 4 | 23 | 16 | 3 |
| 2021-10-05 | -8 | 47 | 8 | 10 | 18 | 3 |
| 2021-10-06 | 1 | 63 | 17 | 9 | 21 | 3 |
| 2021-10-07 | 0 | 53 | 22 | 17 | 20 | 2 |
| 2021-10-08 | 3 | 55 | 26 | 57 | 22 | 2 |
| 2021-10-09 | -4 | 46 | 19 | 70 | 8 | 3 |
| 2021-10-10 | -15 | 38 | 11 | 33 | -9 | 7 |

| | | | | | | |
|---|---|---|---|---|---|---|
| 2021-10-11 | -10 | 40 | 4 | 25 | -16 | 9 |
| 2021-10-12 | -36 | -2 | -26 | 6 | -52 | 17 |
| 2021-10-13 | 0 | 60 | 4 | 25 | 19 | 3 |
| 2021-10-14 | -4 | 53 | 20 | 32 | 19 | 3 |
| 2021-10-15 | 2 | 53 | 47 | 38 | 13 | 3 |
| 2021-10-16 | -9 | 39 | 25 | 44 | 10 | 4 |
| 2021-10-17 | -17 | 43 | 6 | 41 | -4 | 7 |
| 2021-10-18 | -13 | 31 | -3 | 33 | 16 | 4 |
| 2021-10-19 | -13 | 36 | -9 | 17 | 16 | 4 |
| 2021-10-20 | -9 | 49 | -14 | 7 | 19 | 5 |
| 2021-10-21 | -9 | 44 | -2 | 21 | 20 | 4 |
| 2021-10-22 | -1 | 56 | 35 | 33 | 22 | 3 |
| 2021-10-23 | -6 | 44 | 30 | 44 | 11 | 4 |
| 2021-10-24 | -16 | 42 | 15 | 50 | -4 | 6 |
| 2021-10-25 | -13 | 37 | -3 | 19 | 18 | 3 |
| 2021-10-26 | -9 | 41 | -3 | 15 | 19 | 3 |
| 2021-10-27 | 0 | 55 | 2 | 6 | 23 | 3 |
| 2021-10-28 | -2 | 50 | 16 | 17 | 22 | 2 |
| 2021-10-29 | 1 | 54 | 23 | 38 | 24 | 2 |
| 2021-10-30 | -5 | 48 | 11 | 52 | 12 | 3 |
| 2021-10-31 | -8 | 57 | 24 | 36 | -2 | 6 |
| 2021-11-01 | -19 | 25 | -20 | 11 | -14 | 10 |
| 2021-11-02 | -39 | 4 | -14 | 6 | -51 | 16 |
| 2021-11-03 | 6 | 68 | 0 | 35 | 24 | 1 |
| 2021-11-04 | 4 | 58 | 14 | 24 | 23 | 2 |
| 2021-11-05 | 11 | 64 | 36 | 47 | 26 | 1 |
| 2021-11-06 | 4 | 54 | 25 | 55 | 16 | 1 |
| 2021-11-07 | 5 | 73 | 40 | 61 | 4 | 3 |
| 2021-11-08 | 0 | 51 | 3 | 31 | 22 | 1 |
| 2021-11-09 | -1 | 50 | 4 | 26 | 22 | 1 |
| 2021-11-10 | 3 | 58 | -7 | 20 | 25 | 2 |
| 2021-11-11 | -5 | 39 | -8 | 23 | 22 | 2 |
| 2021-11-12 | 3 | 56 | 17 | 53 | 25 | 2 |
| 2021-11-13 | 0 | 47 | 20 | 81 | 11 | 2 |
| 2021-11-14 | 5 | 60 | 85 | 34 | 0 | 4 |
| 2021-11-15 | -27 | 5 | 56 | 36 | -54 | 11 |
| 2021-11-16 | 0 | 55 | 12 | 39 | 22 | 0 |
| 2021-11-17 | 6 | 62 | 16 | 33 | 27 | 1 |
| 2021-11-18 | 2 | 51 | 24 | 21 | 25 | 1 |
| 2021-11-19 | 2 | 50 | 19 | 44 | 28 | 2 |
| 2021-11-20 | -8 | 41 | -2 | 39 | -1 | 5 |
| 2021-11-21 | -2 | 61 | 31 | 55 | 4 | 4 |

| | | | | | | |
|---|---|---|---|---|---|---|
| 2021-11-22 | 0 | 51 | 25 | 35 | 24 | 1 |
| 2021-11-23 | 1 | 51 | 15 | 21 | 26 | 1 |
| 2021-11-24 | 9 | 77 | 16 | 17 | 28 | 0 |
| 2021-11-25 | 6 | 63 | 19 | 24 | 27 | 0 |
| 2021-11-26 | 14 | 81 | 31 | 45 | 30 | 0 |
| 2021-11-27 | -2 | 46 | 37 | 40 | 17 | 2 |
| 2021-11-28 | -2 | 61 | 17 | 55 | 1 | 4 |
| 2021-11-29 | 0 | 50 | 19 | 31 | 25 | 0 |
| 2021-11-30 | -1 | 51 | -1 | 30 | 25 | 1 |
| 2021-12-01 | 5 | 57 | -2 | 22 | 26 | 1 |
| 2021-12-02 | 9 | 60 | 12 | 30 | 26 | 0 |
| 2021-12-03 | 7 | 55 | 30 | 49 | 30 | 1 |
| 2021-12-04 | 10 | 55 | 33 | 56 | 17 | 0 |
| 2021-12-05 | 21 | 82 | 55 | 64 | 12 | 1 |
| 2021-12-06 | -6 | 39 | -10 | 24 | 21 | 2 |
| 2021-12-07 | 4 | 58 | 5 | 29 | 25 | 1 |
| 2021-12-08 | 2 | 65 | -10 | 18 | 25 | 2 |
| 2021-12-09 | 6 | 70 | 12 | 31 | 28 | 0 |
| 2021-12-10 | 9 | 62 | 36 | 59 | 31 | 0 |
| 2021-12-11 | 7 | 59 | 29 | 85 | 21 | 0 |
| 2021-12-12 | 14 | 80 | 44 | 71 | 15 | 2 |
| 2021-12-13 | -3 | 44 | -1 | 37 | 23 | 1 |
| 2021-12-14 | 1 | 57 | 8 | 33 | 24 | 1 |
| 2021-12-15 | 8 | 62 | 0 | 32 | 25 | 2 |
| 2021-12-16 | 13 | 67 | 14 | 34 | 21 | 1 |
| 2021-12-17 | 4 | 50 | 12 | 49 | 24 | 1 |
| 2021-12-18 | 11 | 58 | -1 | 69 | 22 | 0 |
| 2021-12-19 | 34 | 111 | 27 | 78 | 33 | 1 |
| 2021-12-20 | 17 | 73 | 11 | 53 | 7 | 1 |
| 2021-12-21 | 18 | 81 | 17 | 48 | 4 | 2 |
| 2021-12-22 | 29 | 107 | 20 | 51 | 1 | 1 |
| 2021-12-23 | 29 | 114 | 19 | 87 | -10 | 2 |
| 2021-12-24 | -5 | 87 | 1 | 57 | -40 | 7 |
| 2021-12-25 | -64 | -59 | -35 | 0 | -62 | 5 |
| 2021-12-26 | -9 | 39 | 34 | 120 | -7 | 2 |
| 2021-12-27 | -15 | 31 | 4 | 63 | -29 | 8 |
| 2021-12-28 | -11 | 38 | 0 | 50 | -29 | 8 |
| 2021-12-29 | -3 | 59 | 12 | 55 | -28 | 7 |
| 2021-12-30 | -10 | 56 | 5 | 99 | -34 | 7 |
| 2021-12-31 | -35 | 39 | 13 | 56 | -57 | 12 |
| 2022-01-01 | -66 | -72 | -19 | -14 | -65 | 9 |
| 2022-01-02 | -16 | 22 | 24 | 115 | -15 | 2 |

| | | | | | | |
|---|---|---|---|---|---|---|
| 2022-01-03 | -18 | 36 | -9 | 53 | -22 | 7 |
| 2022-01-04 | -15 | 40 | -1 | 30 | -13 | 7 |
| 2022-01-05 | -6 | 54 | 5 | 21 | -7 | 7 |
| 2022-01-06 | -14 | 43 | -5 | 33 | -9 | 8 |
| 2022-01-07 | -16 | 31 | -9 | 45 | -3 | 9 |
| 2022-01-08 | -27 | 20 | -35 | 36 | 0 | 9 |
| 2022-01-09 | -28 | 28 | -34 | 34 | -4 | 10 |
| 2022-01-10 | -15 | 36 | -15 | 26 | 4 | 6 |
| 2022-01-11 | -11 | 46 | 0 | 23 | 6 | 5 |
| 2022-01-12 | -5 | 57 | 2 | 15 | 10 | 5 |
| 2022-01-13 | -9 | 44 | 10 | 18 | 7 | 4 |
| 2022-01-14 | -5 | 46 | 26 | 37 | 13 | 4 |
| 2022-01-15 | -14 | 39 | 17 | 35 | 8 | 4 |
| 2022-01-16 | -5 | 62 | 37 | 40 | 2 | 3 |
| 2022-01-17 | -8 | 47 | 12 | 24 | 7 | 3 |
| 2022-01-18 | -10 | 45 | 13 | 15 | 7 | 4 |
| 2022-01-19 | -4 | 48 | 14 | 21 | 10 | 5 |
| 2022-01-20 | -4 | 50 | 39 | 22 | 6 | 4 |
| 2022-01-21 | -2 | 48 | 48 | 42 | 10 | 5 |
| 2022-01-22 | -13 | 32 | 35 | 49 | 6 | 4 |
| 2022-01-23 | -6 | 53 | 74 | 54 | -3 | 3 |
| 2022-01-24 | -11 | 41 | 23 | 21 | 5 | 4 |
| 2022-01-25 | -11 | 42 | 28 | 15 | 6 | 5 |
| 2022-01-26 | -5 | 49 | 23 | 11 | 8 | 5 |
| 2022-01-27 | -7 | 41 | 33 | 14 | 7 | 5 |
| 2022-01-28 | -7 | 43 | 47 | 37 | 10 | 5 |
| 2022-01-29 | -19 | 26 | 6 | 37 | 3 | 6 |
| 2022-01-30 | -16 | 40 | 28 | 46 | -3 | 5 |
| 2022-01-31 | -10 | 44 | 16 | 23 | 9 | 4 |
| 2022-02-01 | -11 | 40 | 1 | 24 | 10 | 5 |
| 2022-02-02 | -9 | 43 | -7 | 14 | 17 | 5 |
| 2022-02-03 | -8 | 48 | 8 | 20 | 16 | 4 |
| 2022-02-04 | 4 | 60 | 72 | 49 | 21 | 3 |
| 2022-02-05 | -3 | 53 | 25 | 54 | 12 | 3 |
| 2022-02-06 | 2 | 69 | 56 | 58 | 5 | 3 |
| 2022-02-07 | -2 | 48 | 3 | 32 | 19 | 1 |
| 2022-02-08 | -1 | 45 | -3 | 21 | 21 | 1 |
| 2022-02-09 | -1 | 55 | -8 | 14 | 21 | 3 |
| 2022-02-10 | 1 | 46 | 2 | 23 | 21 | 2 |
| 2022-02-11 | 6 | 53 | 33 | 49 | 27 | 2 |
| 2022-02-12 | -10 | 33 | -13 | 48 | 12 | 5 |
| 2022-02-13 | 2 | 60 | 39 | 81 | 5 | 4 |

| | | | | | | |
|---|---|---|---|---|---|---|
| 2022-02-14 | 3 | 47 | 21 | 32 | 23 | 0 |
| 2022-02-15 | 3 | 55 | 15 | 24 | 24 | 0 |
| 2022-02-16 | 7 | 60 | 9 | 24 | 28 | 1 |
| 2022-02-17 | 5 | 56 | 20 | 27 | 27 | 0 |
| 2022-02-18 | 4 | 51 | 26 | 43 | 31 | 0 |
| 2022-02-19 | -10 | 34 | -13 | 50 | 14 | 3 |
| 2022-02-20 | -5 | 60 | 20 | 59 | 6 | 3 |
| 2022-02-21 | -6 | 37 | 1 | 31 | 26 | 0 |
| 2022-02-22 | -5 | 38 | 3 | 28 | 27 | 0 |
| 2022-02-23 | 3 | 61 | 22 | 36 | 30 | 0 |
| 2022-02-24 | 6 | 59 | 32 | 47 | 28 | -1 |
| 2022-02-25 | 13 | 64 | 56 | 94 | 30 | -2 |
| 2022-02-26 | -3 | 43 | 55 | 111 | 7 | -1 |
| 2022-02-27 | 17 | 68 | 151 | 69 | 4 | -1 |
| 2022-02-28 | -6 | 41 | 151 | 29 | -44 | 7 |
| 2022-03-01 | -27 | 1 | 110 | 29 | -58 | 9 |
| 2022-03-02 | -1 | 58 | 42 | 76 | -6 | 4 |
| 2022-03-03 | 3 | 60 | 40 | 64 | 16 | 1 |
| 2022-03-04 | 1 | 54 | 89 | 58 | 21 | 3 |
| 2022-03-05 | -4 | 46 | 38 | 79 | 15 | 1 |
| 2022-03-06 | 3 | 66 | 69 | 108 | 11 | 0 |
| 2022-03-07 | -1 | 54 | 24 | 58 | 29 | 0 |
| 2022-03-08 | 1 | 61 | 22 | 64 | 31 | -1 |
| 2022-03-09 | 5 | 69 | 23 | 46 | 35 | 0 |
| 2022-03-10 | 0 | 58 | 29 | 62 | 35 | 0 |
| 2022-03-11 | 1 | 55 | 58 | 77 | 39 | 1 |
| 2022-03-12 | -9 | 41 | 30 | 90 | 16 | 3 |
| 2022-03-13 | -3 | 59 | 68 | 114 | 9 | 2 |
| 2022-03-14 | -8 | 47 | 17 | 51 | 34 | 1 |
| 2022-03-15 | -9 | 49 | 14 | 52 | 34 | 1 |
| 2022-03-16 | 0 | 65 | 26 | 53 | 37 | 1 |
| 2022-03-17 | -3 | 53 | 31 | 53 | 35 | 1 |
| 2022-03-18 | 0 | 53 | 54 | 83 | 38 | 0 |
| 2022-03-19 | -7 | 43 | 31 | 94 | 16 | 2 |
| 2022-03-20 | -10 | 54 | 67 | 103 | 7 | 3 |
| 2022-03-21 | -14 | 39 | 2 | 55 | 31 | 3 |
| 2022-03-22 | -11 | 47 | 16 | 53 | 33 | 2 |
| 2022-03-23 | -5 | 58 | 18 | 67 | 38 | 2 |
| 2022-03-24 | -5 | 51 | 26 | 71 | 35 | 1 |
| 2022-03-25 | 1 | 53 | 52 | 88 | 40 | 1 |
| 2022-03-26 | -9 | 40 | 42 | 91 | 13 | 3 |
| 2022-03-27 | -11 | 53 | 60 | 105 | 8 | 3 |

| | | | | | | |
|---|---|---|---|---|---|---|
| 2022-03-28 | -15 | 34 | 12 | 70 | 34 | 2 |
| 2022-03-29 | -11 | 48 | 16 | 61 | 35 | 2 |
| 2022-03-30 | 0 | 65 | 23 | 64 | 40 | 1 |
| 2022-03-31 | -4 | 57 | 24 | 68 | 36 | 1 |
| 2022-04-01 | 1 | 56 | 46 | 97 | 42 | 0 |
| 2022-04-02 | -15 | 33 | 3 | 89 | 15 | 4 |
| 2022-04-03 | -6 | 62 | 51 | 94 | 9 | 4 |
| 2022-04-04 | -7 | 53 | 23 | 66 | 37 | 0 |
| 2022-04-05 | -6 | 55 | 15 | 35 | 37 | 0 |
| 2022-04-06 | 1 | 70 | 26 | 41 | 41 | 0 |
| 2022-04-07 | 1 | 66 | 31 | 50 | 38 | 0 |
| 2022-04-08 | 7 | 67 | 53 | 91 | 43 | -1 |

### 6 Description of the database sources in this manuscript

| Data | Source | Source Address | Accessed |
|---|---|---|---|
| Basic education dataset | Anísio Teixeira National Institute for Educational Studies and Research. "INEP" | https://inepdata.inep.gov.br/analytics/saw.dll?Dashboard&PortalPath=%2Fshared%2FCenso%20da%20Educa%C3%A7%C3%A3o%20B%C3%A1sica%2F_portal%2FCat%C3%A1logo%20de%20Escolas&Page=Lista%20das%20Escolas&P1=dashboard&Action=Navigate&ViewState=3hl5ae08n83u3c11a6ttaji3m2&P16=NavRuleDefault&NavFromViewID=d%3Adashboard~p%3Asf156n9k0qs70741 | April 04, 2022. |
| Colored flags dataset | Nova Friburgo's Official Social Media | https://www.instagram.com/prefeituranovafriburgo/ | May 14, 2021-April 07, 2022. |
| | Nova Friburgo's Official Website | https://www.pmnf.rj.gov.br/ | May 14, 2021-June 19, 2022. |
| | Rio de Janeiro State Government's Official Panel for Monitoring COVID-19 | https://painel.saude.rj.gov.br/monitoramento/covid19.html# | May 14, 2021-April 01, 2022. |
| Community mobility | Google COVID-19 Community Mobility Reports | https://www.google.com/covid19/mobility/ | April 13, 2022. |

| | | | |
|---|---|---|---|
| COVID-19 Cases and Deaths | Nova Friburgo's Official Social Media<br>Nova Friburgo's Transparency Portal | https://www.instagram.com/prefeituranovafriburgo/<br>http://novafriburgo-rj.portaltp.com.br/consultas/documentos.aspx?id=145 | May 14, 2021-April 07, 2022.<br>May 14, 2021-April 05, 2022. |
| COVID-19 Cases per groups | Nova Friburgo's Official Panel for Monitoring COVID-19 | https://covid19.novafriburgo.rj.gov.br/index/?data=24%2F06%2F2021 | May 14, 2021-April 05, 2022. |
| COVID-19 Variants dataset | Genomic Network Dashboard . "FIOCRUZ"<br>Phylodynamics of Pandemic Coronavirus in Brazil. "GISAID"<br>Rio de Janeiro State Government's Official Panel for Monitoring COVID-19 | http://www.genomahcov.fiocruz.br/dashboard-en/<br>https://www.gisaid.org/phylodynamics/brazil/)<br>https://painel.saude.rj.gov.br/monitoramento/covid19.html# | May 03, 2022.<br>April 28, 2022.<br>May 14, 2021-April 01, 2022. |
| Hospital systems and beds | National Registry of Health Establishments. "CNES"<br>Nova Friburgo's Official Social Media<br>Rio de Janeiro State Government's Official Panel for Monitoring COVID-19 | http://cnes.datasus.gov.br/pages/estabelecimentos/consulta.jsp<br>https://www.instagram.com/prefeituranovafriburgo/<br>https://painel.saude.rj.gov.br/monitoramento/covid19.html# | April 01, 2022.<br>May 14, 2021-April 07, 2022.<br>May 14, 2021-April 01, 2022. |
| Population dataset | Brazilian Institute of Geography and Statistics. "IBGE" | https://cidades.ibge.gov.br/brasil/rj/nova-friburgo/panorama<br>https://cidades.ibge.gov.br/brasil/rj/nova-friburgo/pesquisa/23/22714<br>https://cidades.ibge.gov.br/ | April 01, 2022.<br>April 01, 2022.<br>April 01, 2022. |
| City Profile | Brazilian Institute of Geography and Statistics. "IBGE"<br>Municipal Law nº4692 | https://cidades.ibge.gov.br/brasil/rj/nova-friburgo/panorama<br>https://www.pmnf.rj.gov.br/uploads/pagina/arquivos/Lei-Muni | June 23, 2021.<br>April 01, 2022. |

| | | | |
|---|---|---|---|
| | Nova Friburgo's Official Website | cipal-4692-2019-Bairros.pdf<br>https://www.pmnf.rj.gov.br/pagina/1_A-Cidade.html | April 03, 2022. |
| Regulamentation and health events | Nova Friburgo's -Official Social Media | https://www.instagram.com/prefeituranovafriburgo/ | May 14, 2021-April 07, 2022. |
| | Nova Friburgo's Official Website | https://www.pmnf.rj.gov.br/ | May 14, 2021-April 05, 2022. |
| | Nova Friburgo's Transparency Portal | http://novafriburgo-rj.portaltp.com.br/consultas/documentos.aspx?id=132 | May 14, 2021-April 05, 2022. |
| | Nova Friburgo's Official News Website | https://www.pmnf.rj.gov.br/noticias/ | May 14, 2021-April 05, 2022. |
| Vaccination progress | Nova Friburgo's -Official Social Media | https://www.instagram.com/prefeituranovafriburgo/ | May 14, 2021-April 07, 2022. |
| | Nova Friburgo's Transparency Portal | http://novafriburgo-rj.portaltp.com.br/consultas/documentos.aspx?id=222 | May 14, 2021-April 05, 2022. |

### 7 Description of the color flag system indicators

| **Color flag system** | | | | | | |
|---|---|---|---|---|---|---|
| **Indicator** | **Rio de Janeiro State Government's** | **Nova Friburgo municipality Government's** | | | | |
| | **Technical note 01/2020** | **Decree 591** | **Decree 625** | **Decree 645** | **Decree 678** | **Decree 819** |
| Variation in number of COVID-19 Intensive Care Unit (ICU) occupation | ✓ | ✓ | ✓ | ✓ | ✓ | ✓ |
| Variation in number of COVID-19 infirmary occupation | ✓ | - | - | ✓ | ✓ | ✓ |

| | | | | | | |
|---|---|---|---|---|---|---|
| Variation in number of COVID-19 deaths | ✓ | - | - | ✓ | - | - |
| Variation in number of COVID-19 cases | ✓ | - | - | ✓ | ✓ | ✓ |
| Positivity rates (%) | ✓ | - | - | ✓ | - | - |
| Lethality rate | - | - | - | - | ✓ | ✓ |
| Prediction of ICU bed depletion | ✓ | - | - | - | - | - |
